# The global prevalence of female genital mutilation/cutting: A systematic review and meta-analysis of national, regional, facility and school-based studies

**DOI:** 10.1101/2022.03.08.22272068

**Authors:** Leen Farouki, Zeinab El Dirani, Sawsan Abdulrahim, Christelle Akl, Chaza Akik, Stephen J McCall

## Abstract

**Background:** Female Genital Mutilation/Cutting (FGM/C) is a non-medical procedure entailing the modification of the external female genitalia. A description of the prevalence and distribution of FGM/C allows the tracking of progress towards ending FGM/C by 2030 (Sustainable Development Goal (SDG) Target 5.3). This systematic review aimed to examine FGM/C prevalence and types, by World Health Organization (WHO) region and country.

**Methods:** A systematic search using Medical Subject Headings (MeSH) and keywords from 2009 to March 24, 2022 was undertaken in MEDLINE, PubMED, PsycINFO, Web of Science, and Embase to identify studies presenting FGM/C prevalence. Abstract and full-text screening, quality assessment, and data extraction were undertaken by two reviewers. Only nationally representative studies were included in the meta-analysis. Pooled FGM/C prevalence was estimated by random-effects meta-analysis using generalised linear mixed models (GLMM). FGM/C prevalence with 95% confidence intervals (CI), prediction intervals (PI) and FGM/C type were presented separately by women aged 15-49 years and girls aged 0-14 years.

**Findings:** 161 studies met the inclusion criteria and 28 were included in the meta-analysis, of which 22 were from the WHO African region (AFR), 5 from the Eastern Mediterranean region (EMR), and 1 from the South-East Asia (SEAR) region. These studies included data from 397,683 women across 28 countries and 283,437 girls across 23 countries; the pooled prevalence estimate of FGM/C amongst women aged 15-49 years was 38.3% (95% CI: 20.8–59.5%; PI:0.48–98.8%), and 7.25% (95% CI: 3.1–16.0%; PI: 0.1-88.9%) amongst girls aged 0-14 years. Amongst included countries, this gave a total estimated prevalence of 86,080,915 women (95% CI: 46,736,701–133,693,929) and 11,982,031 girls with FGM/C (95% CI: 5,123,351–26,476,156). Somalia had the highest FGM/C prevalence amongst women (99.2%) and Mali had the highest amongst girls (72.7%). The most common type of FGM/C amongst women was “flesh removed” (Type I or II) in 19 countries, and “not sewn closed” (Type I, II, or IV) amongst girls in 9 countries. Among repeated nationally representative studies, FGM/C decreased for women and girls in 23 and 25 countries respectively, although in several countries there was a minor decrease (0-3%) or increase in prevalence. The main limitation of the study methodology is that estimates were based on the available published data, which may not reflect the actual global prevalence of FGM/C.

**Discussion:** In this study, we observed large variation in FGM/C prevalence between countries, and the prevalence appears to be declining in many countries, which is encouraging as it minimises physical and physiological harm for a future generation of women. This prevalence estimate is lower than the actual global prevalence of FGM/C due to data gaps, non-comparable denominators, and unavailable surveys. Yet, considerable policy and community-level interventions are required in many countries to meet the SDG target 5.3.

**Funding:** None

**Registration:** CRD42020186937

**Author Summary:** *Why was this study done?:*

- FGM/C is an extreme form of gender inequality that violates women’s and girls’ human rights, and the practice has lifelong health and economic consequences for women and girls.
- Previous studies on prevalence of FGM/C have used repeated nationally representative cross-sectional studies and found that FGM/C is decreasing in many countries.
- This study aimed to provide a baseline prevalence estimate and to understand the data gaps in prevalence required for tracking progress towards the Sustainable Development Goal (SDG) Target 5.3.

*What did the researchers do and find?:*

- This was a systematic review and meta-analysis of all available studies on FGM/C and it provided a thorough overview of studies published on FGM/C prevalence at a national, sub-regional, school, facility, and community level.
- Approximately 100 million girls and women of reproductive age have experienced FGM/C across 28 countries in three WHO regions, with a prevalence of 38% in women and 7% among girls.
- There were large differences between regions and countries; where some countries practiced FGM/C universally, and FGM/C appeared to be decreasing in 23 countries for women and 25 countries for girls.

*What do these findings mean?:*

- Current findings imply that progress towards SDG 5.3 is attainable in some countries, but much work is required in others, including Egypt, Somalia, Sudan, Indonesia, Guinea, and Mali.
- Evaluation of structural or community level policies and interventions in countries that had a decline in FGM/C will be beneficial for countries that have a high prevalence of FGM/C.
- The prevalence estimate of this study is accurate of the included countries but is an underestimate of the global prevalence due to gaps in available data across the world, which are important to resolve to understand actual progress towards SDG 5.3.

## Introduction

Female Genital Mutilation/Cutting (FGM/C), also referred to as female circumcision, is a non-medical procedure that entails the total or partial removal of external female genitalia and other injuries to the female genital organs [1]. The United Nations Sustainable Development Goal (SDG) target 5.3 on gender equality refers to FGM/C as a harmful traditional practice and calls for ending it by 2030.

While the exact global prevalence of FGM/C is unknown, estimates of FGM/C range from 100-140 million women and girls in African region and the Middle East [2, 3], while UNICEF estimates the global prevalence to be over 200 million women and girls living with FGM/C [1–4]. Nationally representative data show that there is a decline in the prevalence of FMG/C but this is not universal across countries [1, 5, 6]. FGM/C persists due to religious, social, and cultural factors [7]. It is commonly believed to create better marriage prospects because it associates with morality, hygiene, and aesthetics; FGM/C is also believed to curb sexual urges and maintain virginity [8]. However, the procedure has no health benefits; it has resulted in negative health outcomes, including menstrual difficulties, infertility, urinary problems, mental health problems, pregnancy and labour complications severe pain, risk of contracting infections, septicaemia, and even death [9–11]. FGM/C is also an economic burden throughout the life course for girls and women [12].

FGM/C is most often performed on girls between infancy and adolescence, and has been classified into four types [13]. Type I (clitoridectomy) involves the partial or total removal of the prepuce and/or the clitoral gland. Type II involves the partial or total removal of the labia minora and clitoral glans without the excision of the labia majora. Type III (infibulation) involves narrowing the vaginal canal by modifying the labia majora and minora and may also include the removal of the clitoral glans. Type IV involves any other non-medical, harmful procedure, such as cauterization, pricking, and scraping [14]. Risks defer by type; the most severe type, Type III, has the more serious obstetric risks of FGM/C including infant resuscitation, stillbirth, and neonatal death; while Types I and II carry risks of caesarean section or postpartum bleeding [15].

An important aspect of the SDGs is to track progress on ending harmful traditional practices, such as FGM/C. However, to our knowledge, there is no comprehensive review in the literature that provides estimates of FGM/C globally, by World Health Organization (WHO) region, or specific countries, which can be used to track improvements towards SDG 5.3. A review of the prevalence of FGM/C will support efforts to understand the global burden of FGM/C and inform adequate prevention and intervention efforts, and local and international policies. A review of the types of FGM/C will contribute similarly by tracking the prevalence of the severity of the procedure. This systematic review and meta-analysis aimed to examine (1) the prevalence of FGM/C and (2) the proportion of the different types of FGM/C, amongst girls aged 0-14 years and women aged 15-49 years old by country and WHO region.

## Methods

### Search strategy and study selection

In this systematic review and meta-analysis of FGM/C prevalence, separate searches were conducted using MEDLINE, PubMED, PsycINFO, Web of Science, and Embase. Hand searches of the grey literature were conducted through searches of reports from international non-governmental organizations, including UNFPA and UNICEF amongst others, and other Google searches. Hand searches of the bibliographies of relevant systematic reviews were also conducted. Together, these databases provide international and interdisciplinary publications. The search strategy (S1 Methods and Results, S1 Table) was adapted to the format of each database. To present up-to-date data that can be used as a baseline to monitor progress on SDG 5.3 over the last decade, the search was limited to include publications from 2009 until 2020. The search was updated to include publications from 2009 until 2022. The last search in all databases was conducted on March 24^th^, 2022. For nationally representative studies, the hand searches were conducted to include studies prior to 2009 in a post-hoc analysis to present FGM/C prevalence across time. The MeSH term for FGM/C was used when possible; otherwise, keywords were used, including “Female Genital Mutilation,” “Female Genital Alteration,” “Female Circumcision,” and “Female Genital Cutting”. No language restrictions were imposed. The references were imported from each database into EndNote then into systematic review software DistillerSR and duplicates were removed [16].

### Study protocol, registration, and reporting

The reporting of this study was based on the Preferred Reporting Items for Systematic Review (PRISMA) reporting guidelines (S2 PRISMA Checklist) [17, 18]. The prospectively written study protocol is (S3 Study Protocol) available at: https://osf.io/h54bu/ [19] and was registered with PROSPERO, number CRD42020186937.

### Inclusion and exclusion criteria

This systematic review and meta-analysis were part of a larger project on FGM/C prevalence and its determinants [7, 19]. Cohort or cross-sectional studies that reported on FGM/C prevalence at the national level, using representative samples or population-based methods, were included in the systematic review and meta-analysis. Sub-regional, facility, community and school-based studies and studies that used non population-based methods or non-probability sampling designs, including cross-sectional, cohort designs, were included in the systematic review but not in the meta-analysis. Furthermore, case-series in migrant populations outside of countries that practice FGM/C were included to understand the scope of the literature on FGM/C in these countries.

Studies were excluded if they (i) only reported on health outcomes of FGM/C, the attitudes and knowledge of healthcare providers, economic effects, or perceptions of FGM/C, (ii) only used qualitative methods, (iii) were systematic reviews (except for referencing), or (iv) were policy reports, conference proceedings or letters to the editor. If numerous journal articles used the same data source, e.g. secondary data analysis of international surveys, only the original report was included. Other than nationally representative studies, if the same data source completed multiple studies in a given country across time, then the most recent was included. The supplementary material contains further details on the included and excluded studies (S1 Methods and Results, S1 Text).

### Study Screening

Titles and abstracts were screened independently by two reviewers. Articles selected for full-text review were also screened by two reviewers, independently and in duplicates. The reasons for exclusion at both the abstract and full-text stages were recorded. Disagreements between the two reviewers were resolved by discussion and consulting a third reviewer who verified the eligibility of all included studies. The supplementary material contains further details on the screening process (S1 Methods and Results, S2 Table).

### Data extraction and quality assessment

Data were extracted from included articles using a structured data extraction form, uploaded into DistillerSR. Data were extracted by one reviewer and verified by a second reviewer; disagreements were resolved by a third reviewer. Data included in the final tables were verified against the original publication by a further reviewer. Items extracted from studies included study characteristics, sampling methods, design, host country and country of origin, ethnicity, age, age at FGM/C, location of procedure, performer of FGM/C, FGM/C prevalence, and proportion of the different FGM/C types. The FGM/C prevalence in each included study was extracted as a proportion or calculated from the numbers presented. All data items were extracted from the most recent nationally representative studies (e.g. MICS or DHS), while only prevalence estimates were extracted from the older nationally representative studies for the post-hoc analysis. Studies were assessed for risk of bias independently by two reviewers using an adapted tool by Hoy and colleagues, which is specific to prevalence studies [20]. This tool includes nine items that collectively assess the selection bias, representativeness of the sample, validity of the tool, and appropriateness of the estimate. Each item was scored as low or high risk of bias, and each paper was given an overall score rated as low, moderate, or high risk of bias.

### Data Analysis

Because the literature fell into certain categories, namely nationally representative, sub-regional, and non-probability samples, data in the present study were grouped similarly. Prevalence estimates from the different studies were grouped by country, WHO region and study design. Pooled estimates of FGM/C prevalence were only presented from studies with representative samples or population-based methods at a national level, and the most recent survey was used in the meta-analysis. Prevalence estimates were presented separately for women aged 15-49 years old and girls aged 0-14 years old as most studies collected data for women and girls separately as defined by these age groups; and it was considered inappropriate to pool these groups together due to a cohort effect [5, 21]. Studies that estimated FGM/C among girls using the number of women with at least 1 daughter with FGM/C were excluded from the meta-analysis because this does not provide an estimate of prevalence among all girls aged 0-14 years old. The denominator of FGM/C type was the total number of women and girls with FGM/C, respectively. In addition, a post-hoc summary of prevalence estimates of FGM/C for each country was presented across time for both women and girls.

For the meta-analysis, heterogeneity between studies is usually assessed using the *I**^2^*** statistic [22]. Although high values of *I**^2^*** are common in meta-analysis for prevalence studies, prediction intervals are recommended to be presented as a measure of heterogeneity [23]. The prediction interval is the range where a proportion from a future study would be expected to be located within if this study was randomly selected from the same group of studies included in the meta-analysis [24]. τ^2^ values were also presented as a measure of the variance of effect sizes amongst studies [25]. Using data extracted from survey reports, a random-effects meta-analysis was conducted to produce a pooled prevalence across all nationally representative studies and across each WHO region. The random-effects meta-analysis of the pooled prevalence, 95% confidence intervals (CI) and prediction intervals (PI) were estimated using Generalized Linear Mixed Models (GLMM) [26] through the ‘metaprop’ command within the Meta package, version 4.15-1 [27]. Funnel plots were constructed to inspect visual asymmetry using the funnelR package, version 0.1.0, which was developed for proportion data (S1 Figure and S2 Figure) [28]. To provide the total number of girls (0-14 years old) and women (15-49 years old) with FGM/C, the pooled prevalence estimate was extrapolated against the age-specific population total in 2020, which only included countries that were included in the meta-analysis, using the UN Population Division [29]. All statistical analyses were conducted using R version 4.1.2.

### Protocol amendments

The protocol was amended to include studies in any language and to specify the disaggregation by age group; available at: https://osf.io/h54bu/ (S3 Study Protocol). Other than studies involving migrants, case series and case-control studies were excluded as prevalence cannot be calculated. A data driven analysis was conducted to present prevalence of FMG/C across time from national surveys. A GLMM meta-analysis was used rather than a Freeman-Tukey transformation due to the limitations of the latter approach [26]. We also provided prediction intervals due to recent methodological recommendations and we present total number of women and girls with FGM/C to allow comparison with other global estimates [23].

### Ethical approval and role of the funding source

This was a systematic review of published studies, so no ethical approval was required. There was no funding source for this study.

## Results

Out of 2913 records retrieved from database and hand searches, 417 publications were assessed under full-text review. Of these, a total of 161 were included in the systematic review: 28 nationally representative studies were included in the meta-analysis of the prevalence of FGM/C and two were included in the systematic review but not in the meta-analysis; 33 sub-regional studies; and 98 non population-based studies including 44 on migrant populations (Figure 1). The Indonesia RISKESDAS [30] was not included in the meta-analysis because it did not provide the sample size, and the Pew Research Center survey [31], Eritrea Population and Health Survey [32] and Yemen DHS survey [33] were not included in the meta-analysis of FGM/C prevalence of girls as these surveys had non-comparable denominators.

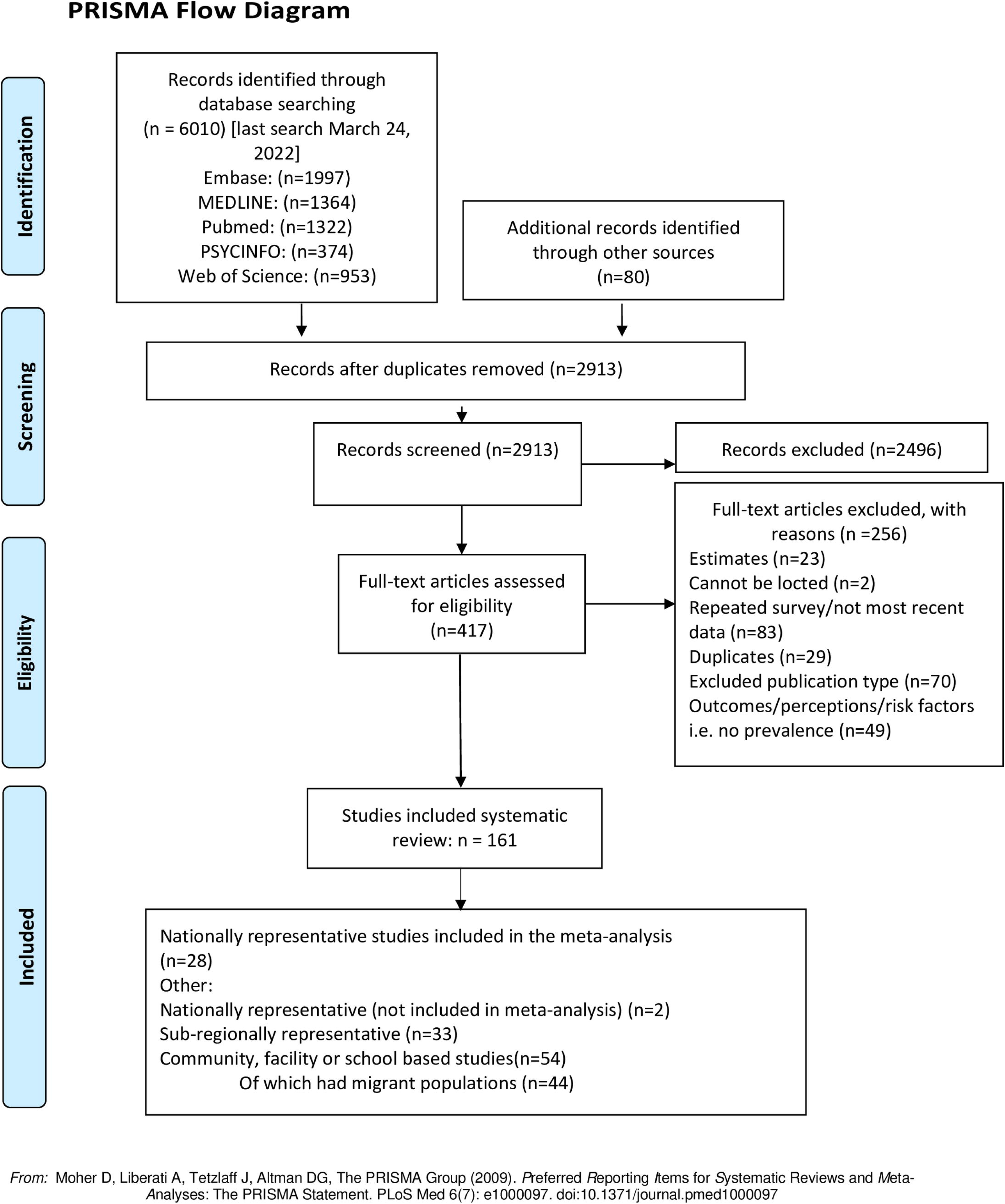

### Nationally representative studies

Of the 30 nationally representative studies, 17 used data from Demographic and Health Surveys (DHS), 10 use data from Multiple Indicator Cluster Surveys (MICS), and three used other population-based surveys (S4 Results, S4 Table). Furthermore, 22 represent the African Region (AFR) [32, 34–54], five represent the Eastern Mediterranean Region (EMR) [33, 55–58], two represent the South-East Asian Region (SEAR) [30, 59], and one represented both EMR and AFR [31]. All national studies reported FGM/C prevalence among the total number of women and girls in surveyed households, except surveys from Liberia (reported on women who have heard of FGM/C) [45], Niger [49], and Uganda [54] that reported only on women, and surveys from Yemen [33], Eritrea [32] and Pew Research Center survey [31] which asked women whether at least one of their daughters had FGM/C. Apart from that of the Pew Research Center, all studies had a low risk of bias and used a cross-sectional design with multi-stage cluster sampling. The Pew Research Center survey had a moderate risk of bias, a cross-sectional design, and used stratified random sampling [31].

The 28 nationally representative studies included in the meta-analysis provided data on women in 28 countries and data on girls in 23 countries. Out of a total of 397,683 women aged 15-49 years in 28 countries, 163,415 women had FGM/C representing a pooled prevalence of 38.3% (CI: 20.8-59.5%; PI: 0.5%-98.8%; τ^2^=5.4) (Table 1 & Figure 2). Prevalence estimates varied considerably by country and ranged from 99.2% in Somalia [58] to 0.3% in Uganda [54]. Out of a total of 283,437 girls aged 0-14 years in 23 countries, 46,713 girls had FGM/C, and this gave a pooled prevalence of 7.3% (95% CI: 3.1-16.0%; PI: 0.1-88.9%; τ^2^=4.8). The country level prevalence ranged between 72.7% in Mali [46] and 0.1% in Ghana [41] (Table 1 & Figure 3). Amongst included countries, the total estimated prevalence was 86,080,915 women (95%CI: 46,736,701–133,693,929) and 11,982,031 girls with FGM/C (95% CI: 5,123,351–26,476,156) (Table 1).

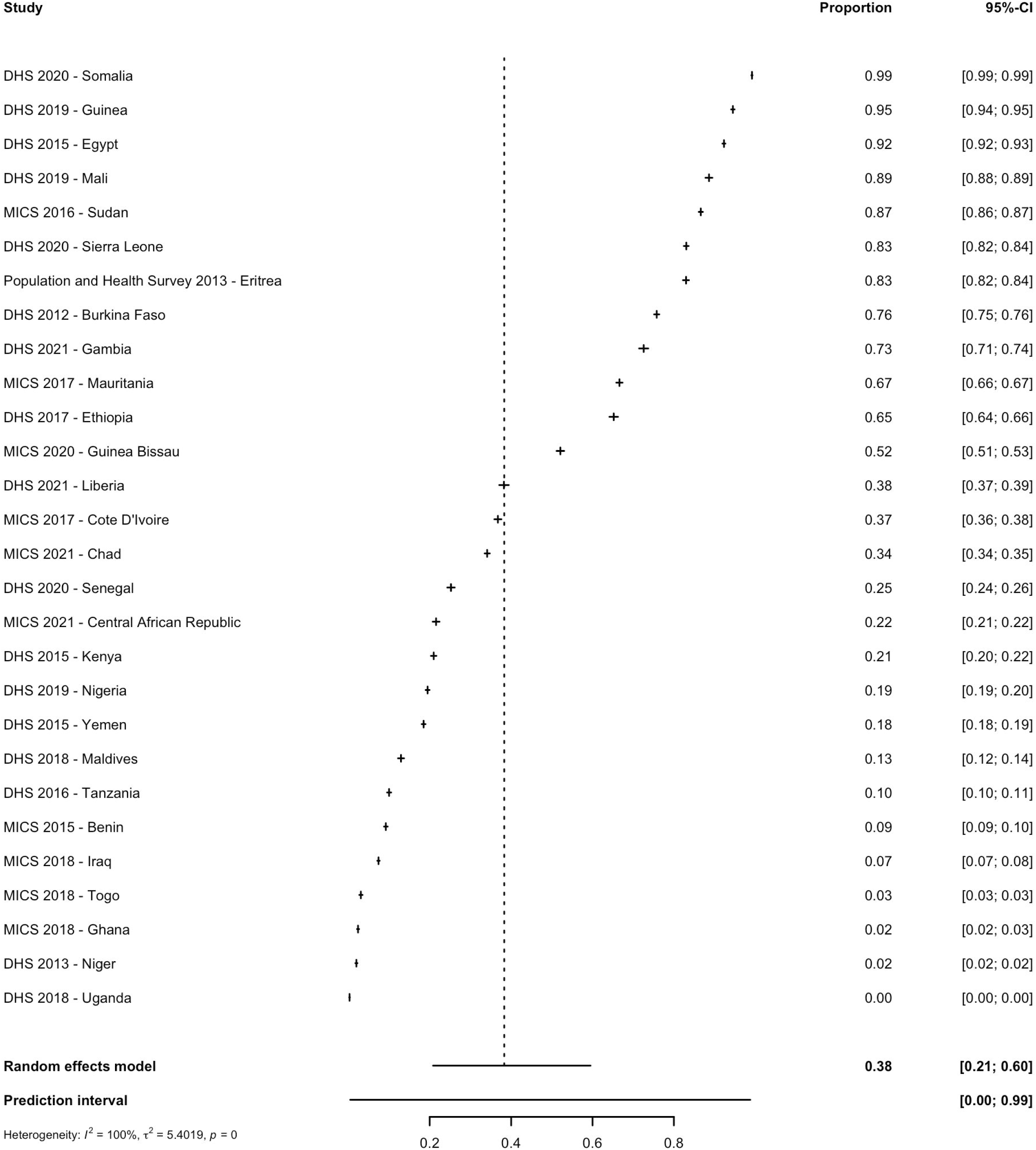

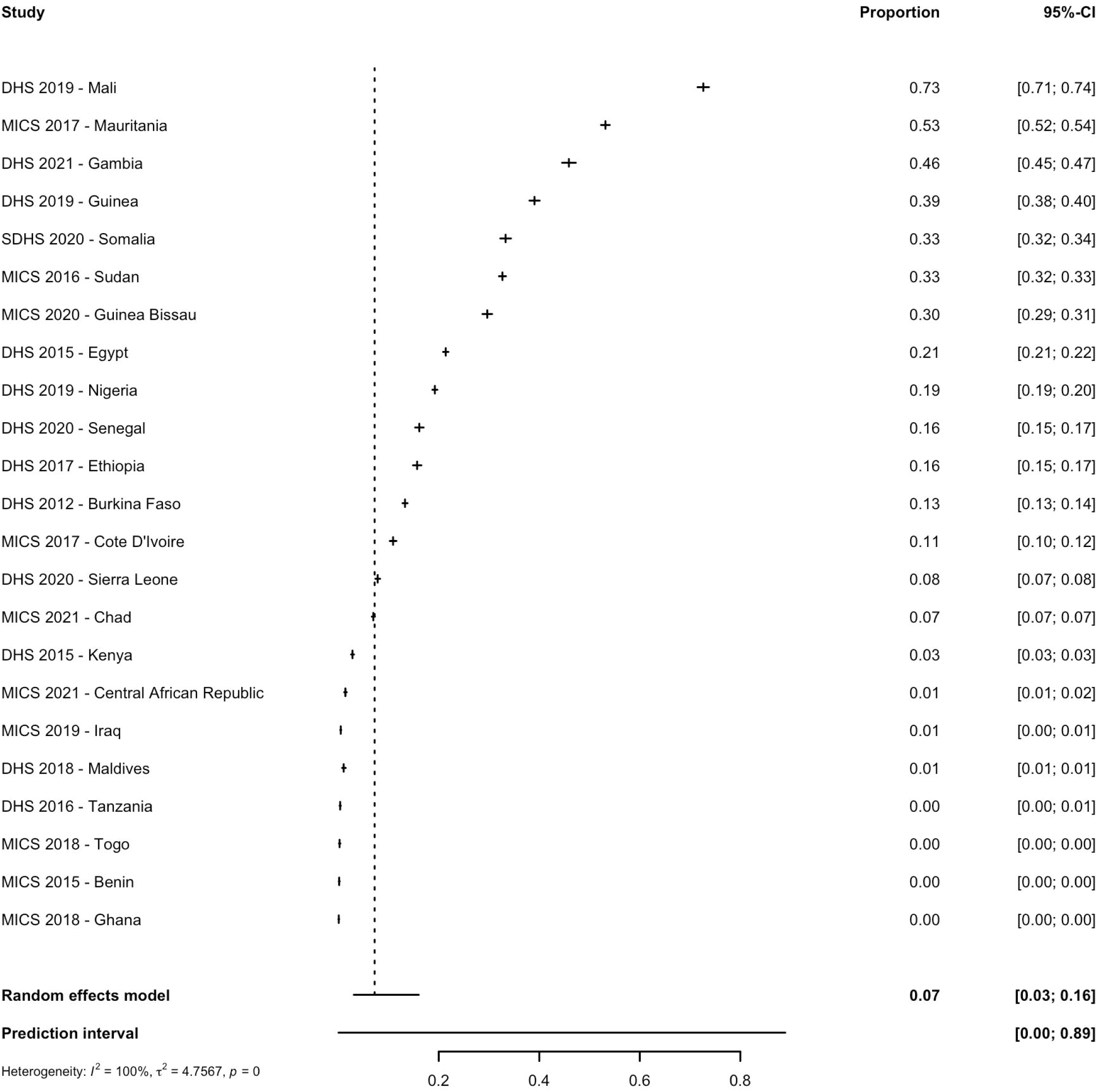

**Table 1.**
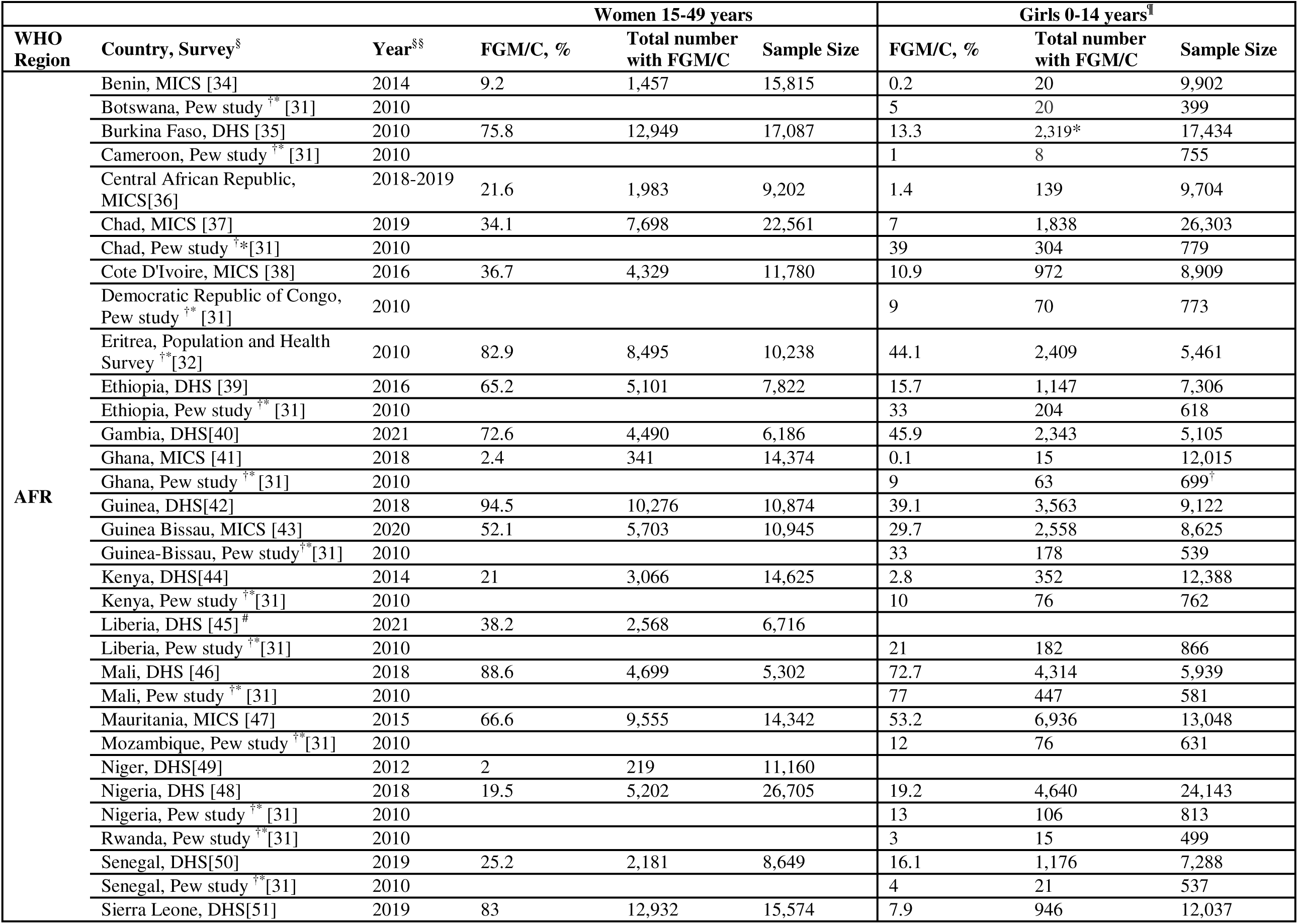
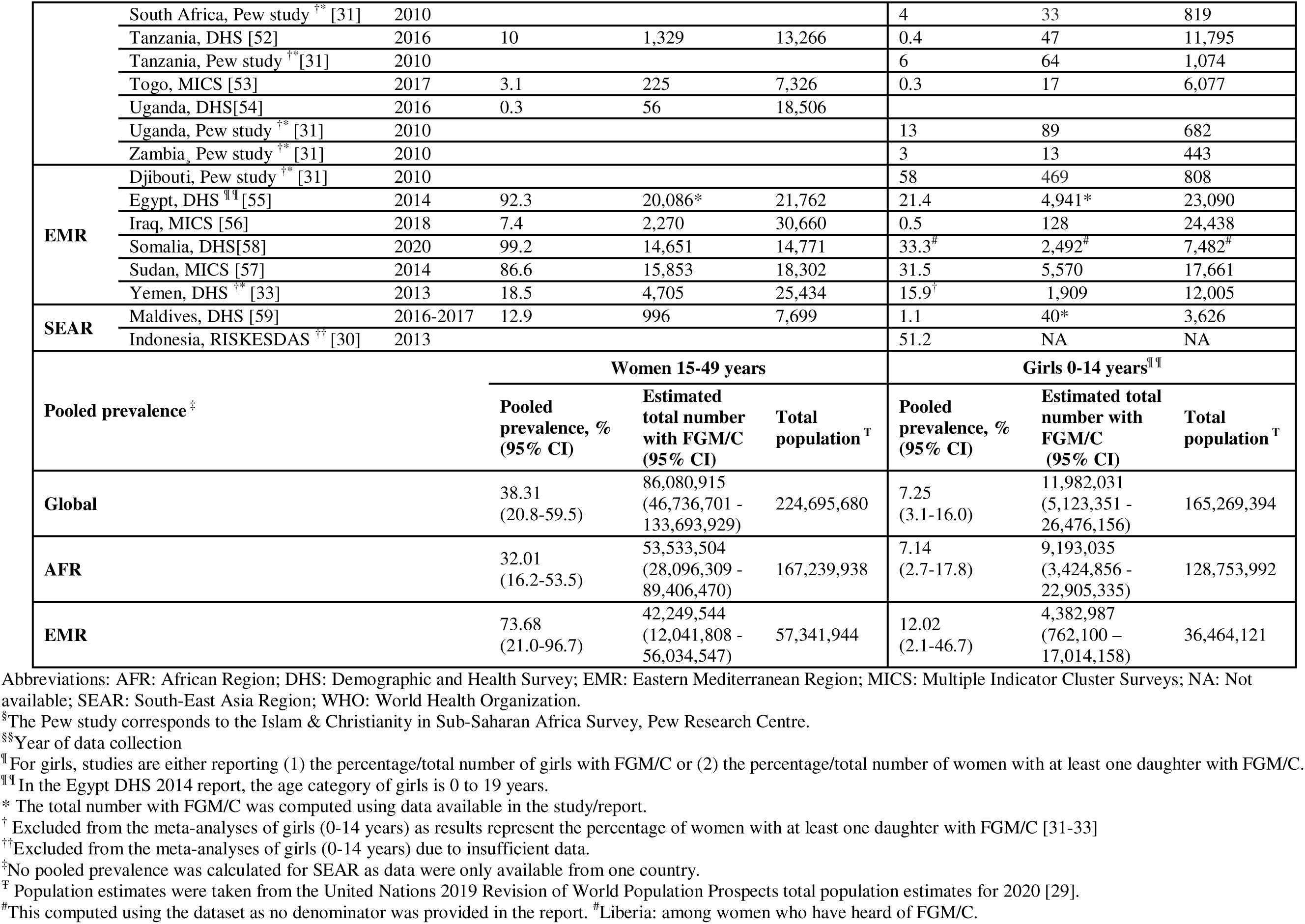
Prevalence of Female Genital Mutilation/Cutting (FGM/C) in Women and Girls in Nationally Representative Studies

| WHO Region | Country, Survey<sup>§</sup> | Year<sup>§§</sup> | Women 15-49 years |  |  | Girls 0-14 years<sup>¶</sup> |  |  |
| --- | --- | --- | --- | --- | --- | --- | --- | --- |
|  |  |  | FGM/C, % | Total number with FGM/C | Sample Size | FGM/C, % | Total number with FGM/C | Sample Size |
| AFR | Benin, MICS [34] | 2014 | 9.2 | 1,457 | 15,815 | 0.2 | 20 | 9,902 |
|  | Botswana, Pew study <sup>†*</sup> [31] | 2010 |  |  |  | 5 | 20 | 399 |
|  | Burkina Faso, DHS [35] | 2010 | 75.8 | 12,949 | 17,087 | 13.3 | 2,319* | 17,434 |
|  | Cameroon, Pew study <sup>†*</sup> [31] | 2010 |  |  |  | 1 | 8 | 755 |
|  | Central African Republic, MICS[36] | 2018-2019 | 21.6 | 1,983 | 9,202 | 1.4 | 139 | 9,704 |
|  | Chad, MICS [37] | 2019 | 34.1 | 7,698 | 22,561 | 7 | 1,838 | 26,303 |
|  | Chad, Pew study <sup>†*</sup>[31] | 2010 |  |  |  | 39 | 304 | 779 |
|  | Cote D'Ivoire, MICS [38] | 2016 | 36.7 | 4,329 | 11,780 | 10.9 | 972 | 8,909 |
|  | Democratic Republic of Congo, Pew study <sup>†*</sup> [31] | 2010 |  |  |  | 9 | 70 | 773 |
|  | Eritrea, Population and Health Survey <sup>†*</sup>[32] | 2010 | 82.9 | 8,495 | 10,238 | 44.1 | 2,409 | 5,461 |
|  | Ethiopia, DHS [39] | 2016 | 65.2 | 5,101 | 7,822 | 15.7 | 1,147 | 7,306 |
|  | Ethiopia, Pew study <sup>†*</sup> [31] | 2010 |  |  |  | 33 | 204 | 618 |
|  | Gambia, DHS[40] | 2021 | 72.6 | 4,490 | 6,186 | 45.9 | 2,343 | 5,105 |
|  | Ghana, MICS [41] | 2018 | 2.4 | 341 | 14,374 | 0.1 | 15 | 12,015 |
|  | Ghana, Pew study <sup>†*</sup> [31] | 2010 |  |  |  | 9 | 63 | 699<sup>†</sup> |
|  | Guinea, DHS[42] | 2018 | 94.5 | 10,276 | 10,874 | 39.1 | 3,563 | 9,122 |
|  | Guinea Bissau, MICS [43] | 2020 | 52.1 | 5,703 | 10,945 | 29.7 | 2,558 | 8,625 |
|  | Guinea-Bissau, Pew study<sup>†*</sup> [31] | 2010 |  |  |  | 33 | 178 | 539 |
|  | Kenya, DHS[44] | 2014 | 21 | 3,066 | 14,625 | 2.8 | 352 | 12,388 |
|  | Kenya, Pew study <sup>†*</sup>[31] | 2010 |  |  |  | 10 | 76 | 762 |
|  | Liberia, DHS [45] <sup>#</sup> | 2021 | 38.2 | 2,568 | 6,716 |  |  |  |
|  | Liberia, Pew study <sup>†*</sup>[31] | 2010 |  |  |  | 21 | 182 | 866 |
|  | Mali, DHS [46] | 2018 | 88.6 | 4,699 | 5,302 | 72.7 | 4,314 | 5,939 |
|  | Mali, Pew study <sup>†*</sup> [31] | 2010 |  |  |  | 77 | 447 | 581 |
|  | Mauritania, MICS [47] | 2015 | 66.6 | 9,555 | 14,342 | 53.2 | 6,936 | 13,048 |
|  | Mozambique, Pew study <sup>†*</sup>[31] | 2010 |  |  |  | 12 | 76 | 631 |
|  | Niger, DHS[49] | 2012 | 2 | 219 | 11,160 |  |  |  |
|  | Nigeria, DHS [48] | 2018 | 19.5 | 5,202 | 26,705 | 19.2 | 4,640 | 24,143 |
|  | Nigeria, Pew study <sup>†*</sup> [31] | 2010 |  |  |  | 13 | 106 | 813 |
|  | Rwanda, Pew study <sup>†*</sup>[31] | 2010 |  |  |  | 3 | 15 | 499 |
|  | Senegal, DHS[50] | 2019 | 25.2 | 2,181 | 8,649 | 16.1 | 1,176 | 7,288 |
|  | Senegal, Pew study <sup>†*</sup>[31] | 2010 |  |  |  | 4 | 21 | 537 |
|  | Sierra Leone, DHS[51] | 2019 | 83 | 12,932 | 15,574 | 7.9 | 946 | 12,037 |

|  | South Africa, Pew study <sup>††</sup> [31] | 2010 |  |  |  | 4 | 33 | 819 |
| --- | --- | --- | --- | --- | --- | --- | --- | --- |
|  | Tanzania, DHS [52] | 2016 | 10 | 1,329 | 13,266 | 0.4 | 47 | 11,795 |
|  | Tanzania, Pew study <sup>††</sup> [31] | 2010 |  |  |  | 6 | 64 | 1,074 |
|  | Togo, MICS [53] | 2017 | 3.1 | 225 | 7,326 | 0.3 | 17 | 6,077 |
|  | Uganda, DHS[54] | 2016 | 0.3 | 56 | 18,506 |  |  |  |
|  | Uganda, Pew study <sup>††</sup> [31] | 2010 |  |  |  | 13 | 89 | 682 |
|  | Zambia, Pew study <sup>††</sup> [31] | 2010 |  |  |  | 3 | 13 | 443 |
| **EMR** | Djibouti, Pew study <sup>††</sup> [31] | 2010 |  |  |  | 58 | 469 | 808 |
|  | Egypt, DHS <sup>††</sup> [55] | 2014 | 92.3 | 20,086* | 21,762 | 21.4 | 4,941* | 23,090 |
|  | Iraq, MICS [56] | 2018 | 7.4 | 2,270 | 30,660 | 0.5 | 128 | 24,438 |
|  | Somalia, DHS[58] | 2020 | 99.2 | 14,651 | 14,771 | 33.3<sup>#</sup> | 2,492<sup>#</sup> | 7,482<sup>#</sup> |
|  | Sudan, MICS [57] | 2014 | 86.6 | 15,853 | 18,302 | 31.5 | 5,570 | 17,661 |
|  | Yemen, DHS <sup>††</sup> [33] | 2013 | 18.5 | 4,705 | 25,434 | 15.9<sup>†</sup> | 1,909 | 12,005 |
| **SEAR** | Maldives, DHS [59] | 2016-2017 | 12.9 | 996 | 7,699 | 1.1 | 40* | 3,626 |
|  | Indonesia, RISKESDAS <sup>††</sup> [30] | 2013 |  |  |  | 51.2 | NA | NA |
|  |  |  | **Women 15-49 years** |  |  | **Girls 0-14 years<sup>††</sup>** |  |  |
| **Pooled prevalence <sup>‡</sup>** |  |  | **Pooled prevalence, % (95% CI)** | **Estimated total number with FGM/C (95% CI)** | **Total population <sup>†</sup>** | **Pooled prevalence, % (95% CI)** | **Estimated total number with FGM/C (95% CI)** | **Total population <sup>†</sup>** |
| **Global** |  |  | 38.31<br>(20.8-59.5) | 86,080,915<br>(46,736,701 - 133,693,929) | 224,695,680 | 7.25<br>(3.1-16.0) | 11,982,031<br>(5,123,351 - 26,476,156) | 165,269,394 |
| **AFR** |  |  | 32.01<br>(16.2-53.5) | 53,533,504<br>(28,096,309 - 89,406,470) | 167,239,938 | 7.14<br>(2.7-17.8) | 9,193,035<br>(3,424,856 - 22,905,335) | 128,753,992 |
| **EMR** |  |  | 73.68<br>(21.0-96.7) | 42,249,544<br>(12,041,808 - 56,034,547) | 57,341,944 | 12.02<br>(2.1-46.7) | 4,382,987<br>(762,100 - 17,014,158) | 36,464,121 |

Abbreviations: AFR: African Region; DHS: Demographic and Health Survey; EMR: Eastern Mediterranean Region; MICS: Multiple Indicator Cluster Surveys; NA: Not available; SEAR: South-East Asia Region; WHO: World Health Organization.
<sup>§</sup>The Pew study corresponds to the Islam & Christianity in Sub-Saharan Africa Survey, Pew Research Centre.
<sup>§§</sup>Year of data collection
<sup>¶</sup>For girls, studies are either reporting (1) the percentage/total number of girls with FGM/C or (2) the percentage/total number of women with at least one daughter with FGM/C.
<sup>¶¶</sup>In the Egypt DHS 2014 report, the age category of girls is 0 to 19 years.
\* The total number with FGM/C was computed using data available in the study/report.
<sup>†</sup> Excluded from the meta-analyses of girls (0-14 years) as results represent the percentage of women with at least one daughter with FGM/C [31-33]
<sup>††</sup> Excluded from the meta-analyses of girls (0-14 years) due to insufficient data.
<sup>‡</sup> No pooled prevalence was calculated for SEAR as data were only available from one country.
<sup>†</sup> Population estimates were taken from the United Nations 2019 Revision of World Population Prospects total population estimates for 2020 [29].
<sup>#</sup>This computed using the dataset as no denominator was provided in the report. <sup>#</sup>Liberia: among women who have heard of FGM/C.

Within AFR, the prevalence amongst women was 32.0% (95% CI: 16.2-53.5%; PI: 0.5-97.8%; τ^2^=4.6) while amongst girls, it was 7.1% (95% CI: 2.7-17.8%; PI: 0.1-90.9%; τ^2^=5.0). This provides a regional estimate of 53,533,504 (95% CI: 28,096,309 – 89,406,470) women with FGM/C and 9,193,035 (95% CI: 3,424,856 – 22,905,335) girls with FGM/C. Within EMR, the prevalence amongst women was 73.7% (95% CI: 21.0-96.7%; PI: 0.02-1%; τ^2^=7.2), while amongst girls it was 12.0% (95% CI: 2.1-46.7%; PI: 0-99.9%; τ^2^=3.6). This provides a EMR regional estimate of 42,249,544 (95% CI: 12,041,808 – 56,034,547) women with FGM/C and 4,382,987 (95% CI: 762,100 – 17,014,158) girls with FGM/C.

Among available nationally representative surveys that ranged between 1994 and 2020, most countries showed a decline in the prevalence of FGM/C across repeated cross-sections of women (23 countries) and girls (25 countries) (Table 2). In addition, among repeated cross-sections of women, 7 countries showed a minor decrease in prevalence (0-3%) and three countries showed an increase in the prevalence of FGM/C. In particular, there was an increase from 97.9% to 99.2% in Somalia (2006 to 2020), from 71.6% to 75.8% in Burkina Faso (1998-99 to 2010), and from 44.5% to 52.1% in Guinea-Bissau (2006 to 2018-19). For repeated cross-sections of girls, 5 countries had a minor decrease in prevalence (0-3%) and two countries had an increase (Djibouti: 48.5% in 2006, to 58% in 2010; and Cameroon: 0.7% in 2004, to 1% in 2010). The largest decline was in Central African Republic (43.4% in 1994-95, to 21.6% in 2018-19) among repeated cross-sections of women; and in Ethiopia from 51.9% in 2000 to 15.7% in 2016, which was among women who reported having at least one daughter who had FGM/C.

**Table 2.**
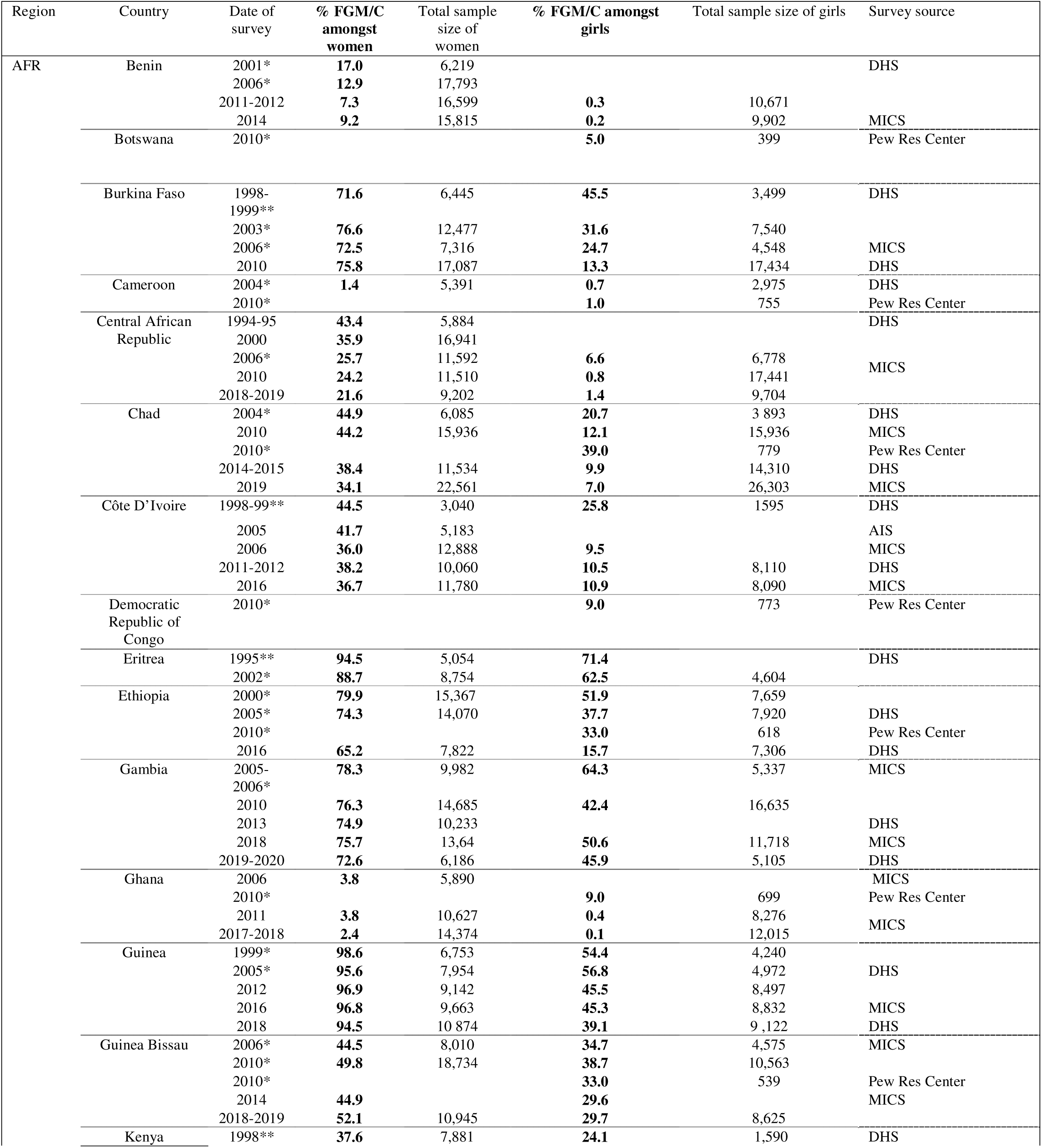
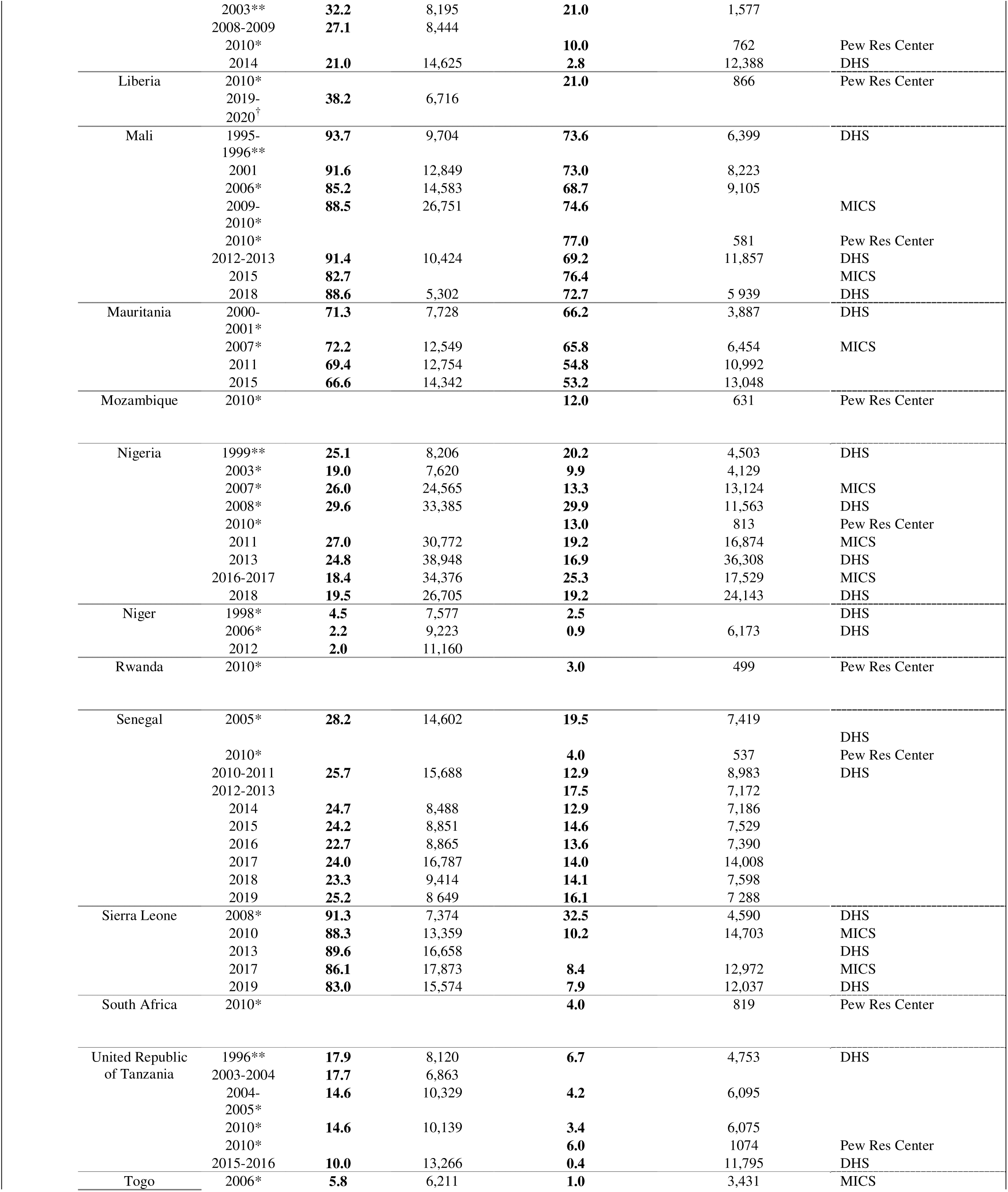
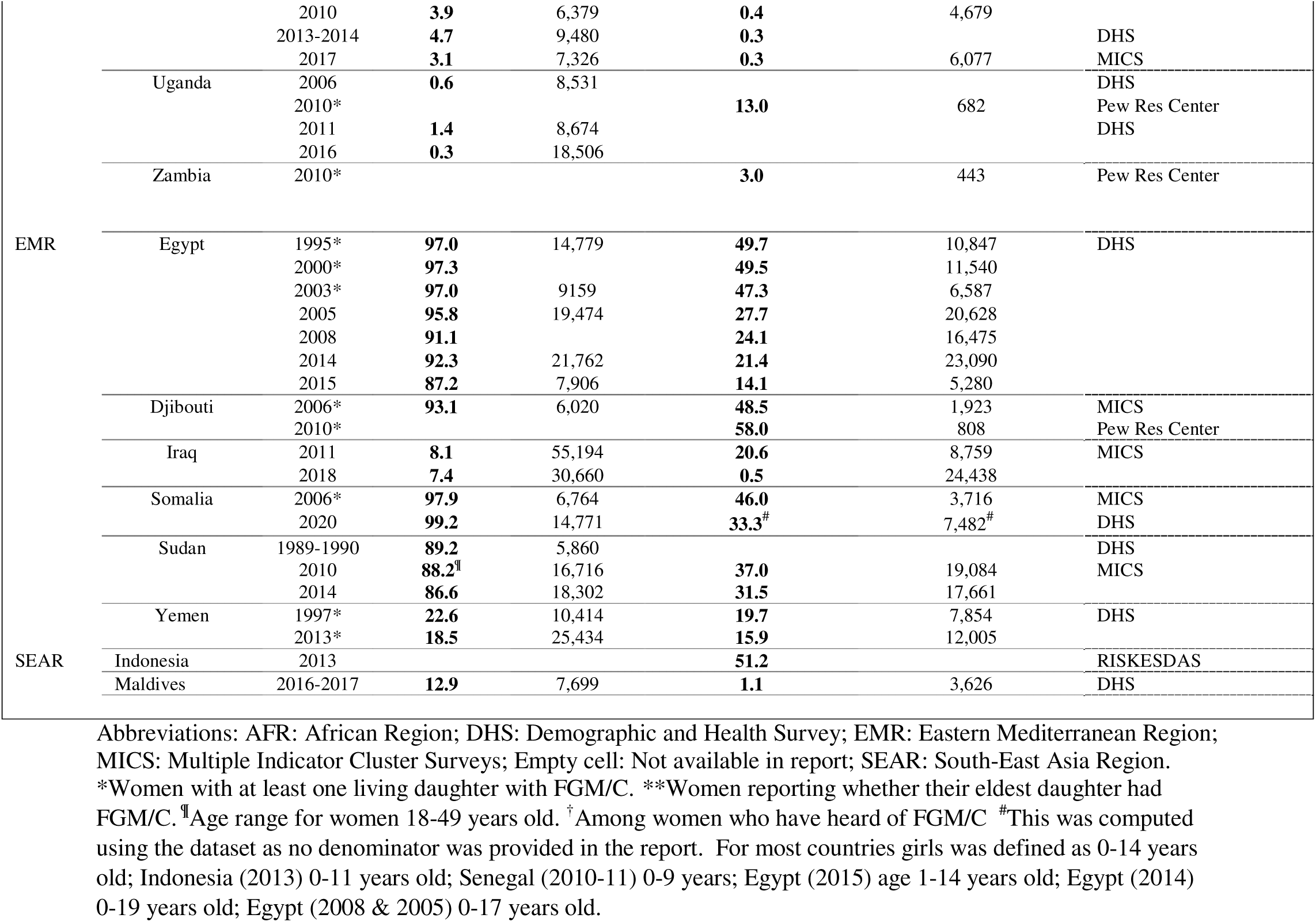
Repeated nationally representative cross-sectional studies reporting the prevalence of Female Genital Mutilation/Cutting (FGM/C) by country

Twenty-three of the 28 national reports recorded FGM/C type for women (Table 3). In MICS and DHS Type I and II were described as “cut with flesh removed”, Type III was described as “sewn closed” and Type IV was described as “nicked” or “cut”. Amongst women, the type “flesh removed” was the most common type in 19 countries, “nicked” was the least common type in 13 countries and “sewn closed” was most common amongst women in two countries (Sudan (77.0%) and Central African Republic (49.6%)). The pooled proportion of women with FGM/C that were “nicked” was 4.8% (95% CI: 2.9-8.1%) (Figure 4a), had “flesh removed” was 65.7% (95% CI: 56.7-73.8%) (Figure 4b), or had their genital area “sewn closed” was 12.1% (CI: 7.4% -19.4%) (Figure 4c). No pooled proportion of types was conducted amongst girls due to inconsistent reporting of types and because the type of FMG/C was only collected in 14 out of 23 countries. Amongst girls with FGM/C, “not sewn closed” and “flesh removed” were the most common type in 6 countries each and “sewn closed” was the least common type in 7 countries although it was the most common type in Sierra Leone (83.3%). Surveys using the terms “not sewn closed” may refer to Types, I, II, and IV (Table 3).

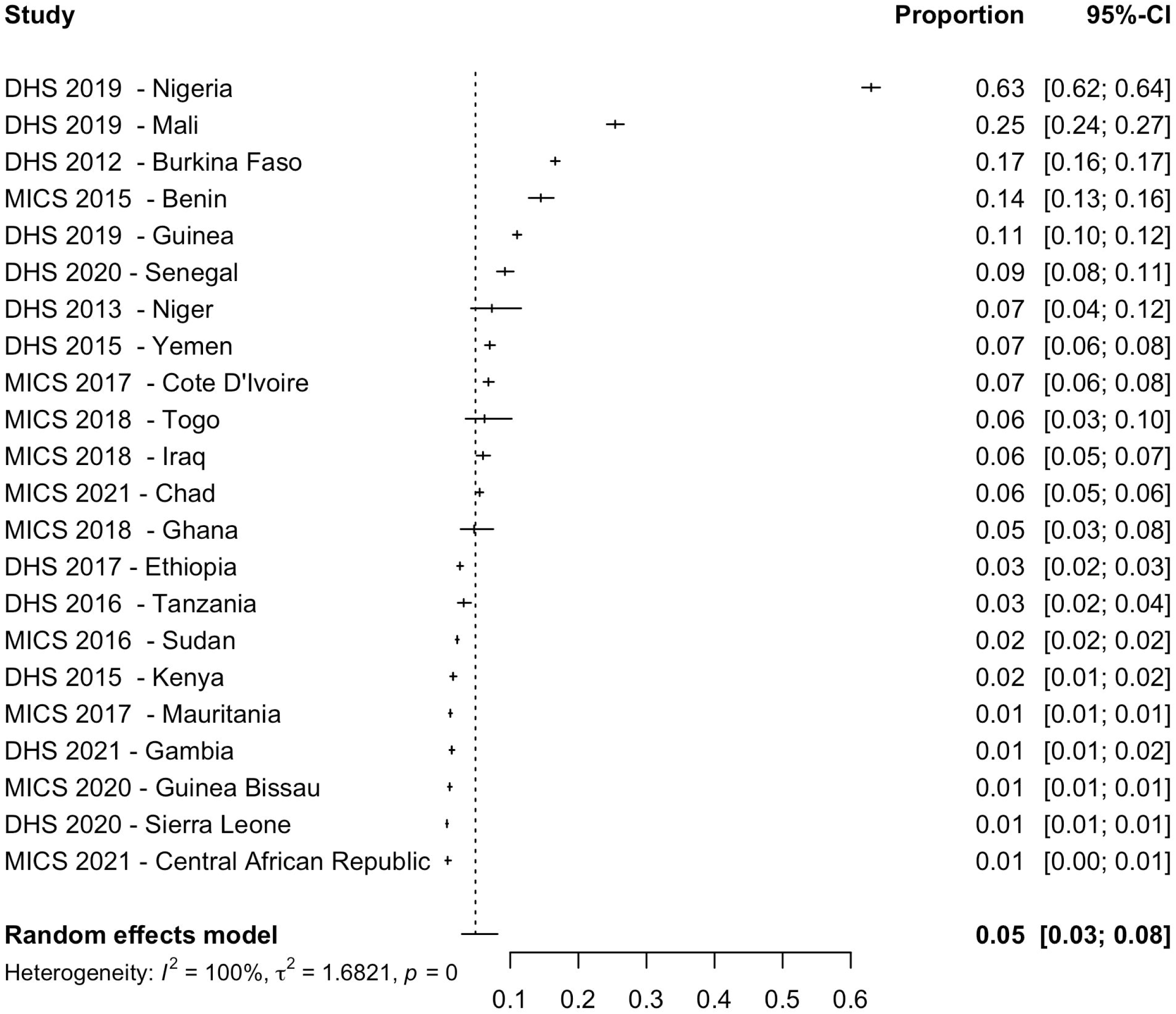

**Table 3.**
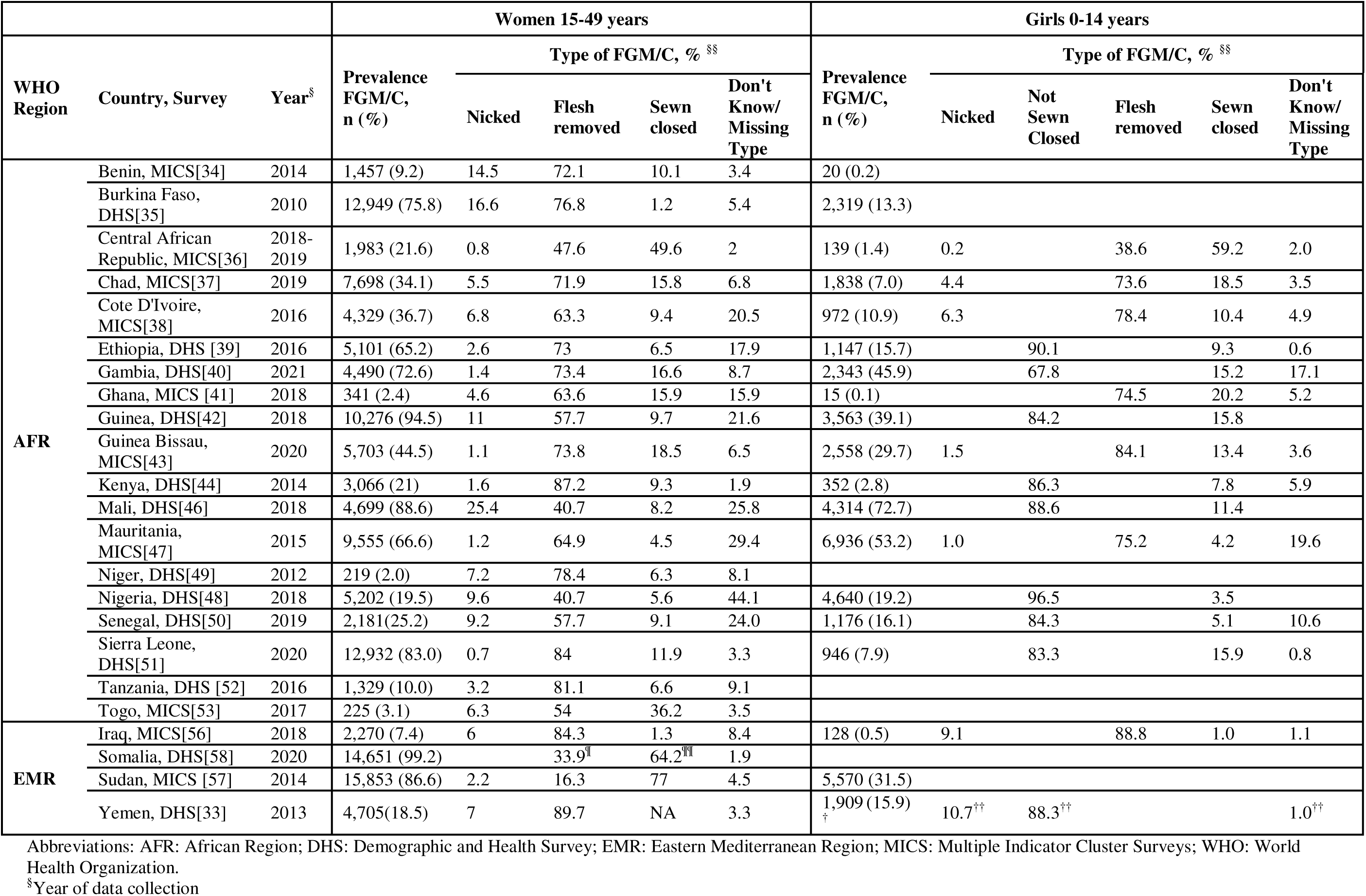
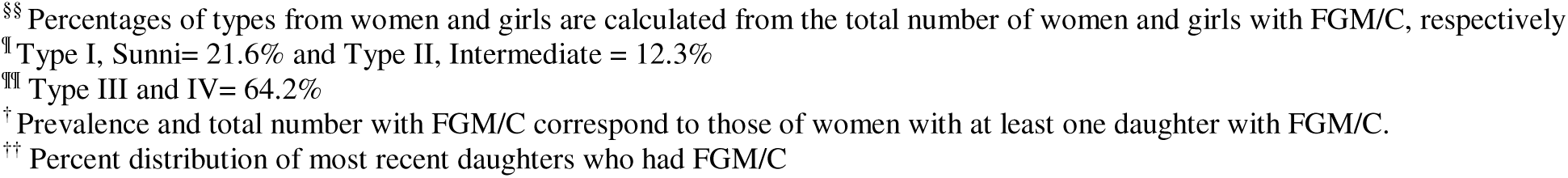
Types of Female Genital Mutilation/Cutting (FGM/C) in Nationally Representative Studies

|  |  |  | Women 15-49 years |  |  |  |  | Girls 0-14 years |  |  |  |  |  |
| --- | --- | --- | --- | --- | --- | --- | --- | --- | --- | --- | --- | --- | --- |
| WHO Region | Country, Survey | Year<sup>§</sup> | Prevalence FGM/C, n (%) | Type of FGM/C, % <sup>§§</sup> |  |  |  | Prevalence FGM/C, n (%) | Type of FGM/C, % <sup>§§</sup> |  |  |  |  |
|  |  |  |  | Nicked | Flesh removed | Sewn closed | Don't Know/ Missing Type |  | Nicked | Not Sewn Closed | Flesh removed | Sewn closed | Don't Know/ Missing Type |
| AFR | Benin, MICS[34] | 2014 | 1,457 (9.2) | 14.5 | 72.1 | 10.1 | 3.4 | 20 (0.2) |  |  |  |  |  |
|  | Burkina Faso, DHS[35] | 2010 | 12,949 (75.8) | 16.6 | 76.8 | 1.2 | 5.4 | 2,319 (13.3) |  |  |  |  |  |
|  | Central African Republic, MICS[36] | 2018-2019 | 1,983 (21.6) | 0.8 | 47.6 | 49.6 | 2 | 139 (1.4) | 0.2 |  | 38.6 | 59.2 | 2.0 |
|  | Chad, MICS[37] | 2019 | 7,698 (34.1) | 5.5 | 71.9 | 15.8 | 6.8 | 1,838 (7.0) | 4.4 |  | 73.6 | 18.5 | 3.5 |
|  | Cote D'Ivoire, MICS[38] | 2016 | 4,329 (36.7) | 6.8 | 63.3 | 9.4 | 20.5 | 972 (10.9) | 6.3 |  | 78.4 | 10.4 | 4.9 |
|  | Ethiopia, DHS [39] | 2016 | 5,101 (65.2) | 2.6 | 73 | 6.5 | 17.9 | 1,147 (15.7) |  | 90.1 |  | 9.3 | 0.6 |
|  | Gambia, DHS[40] | 2021 | 4,490 (72.6) | 1.4 | 73.4 | 16.6 | 8.7 | 2,343 (45.9) |  | 67.8 |  | 15.2 | 17.1 |
|  | Ghana, MICS [41] | 2018 | 341 (2.4) | 4.6 | 63.6 | 15.9 | 15.9 | 15 (0.1) |  |  | 74.5 | 20.2 | 5.2 |
|  | Guinea, DHS[42] | 2018 | 10,276 (94.5) | 11 | 57.7 | 9.7 | 21.6 | 3,563 (39.1) |  | 84.2 |  | 15.8 |  |
|  | Guinea Bissau, MICS[43] | 2020 | 5,703 (44.5) | 1.1 | 73.8 | 18.5 | 6.5 | 2,558 (29.7) | 1.5 |  | 84.1 | 13.4 | 3.6 |
|  | Kenya, DHS[44] | 2014 | 3,066 (21) | 1.6 | 87.2 | 9.3 | 1.9 | 352 (2.8) |  | 86.3 |  | 7.8 | 5.9 |
|  | Mali, DHS[46] | 2018 | 4,699 (88.6) | 25.4 | 40.7 | 8.2 | 25.8 | 4,314 (72.7) |  | 88.6 |  | 11.4 |  |
|  | Mauritania, MICS[47] | 2015 | 9,555 (66.6) | 1.2 | 64.9 | 4.5 | 29.4 | 6,936 (53.2) | 1.0 |  | 75.2 | 4.2 | 19.6 |
|  | Niger, DHS[49] | 2012 | 219 (2.0) | 7.2 | 78.4 | 6.3 | 8.1 |  |  |  |  |  |  |
|  | Nigeria, DHS[48] | 2018 | 5,202 (19.5) | 9.6 | 40.7 | 5.6 | 44.1 | 4,640 (19.2) |  | 96.5 |  | 3.5 |  |
|  | Senegal, DHS[50] | 2019 | 2,181(25.2) | 9.2 | 57.7 | 9.1 | 24.0 | 1,176 (16.1) |  | 84.3 |  | 5.1 | 10.6 |
|  | Sierra Leone, DHS[51] | 2020 | 12,932 (83.0) | 0.7 | 84 | 11.9 | 3.3 | 946 (7.9) |  | 83.3 |  | 15.9 | 0.8 |
|  | Tanzania, DHS [52] | 2016 | 1,329 (10.0) | 3.2 | 81.1 | 6.6 | 9.1 |  |  |  |  |  |  |
|  | Togo, MICS[53] | 2017 | 225 (3.1) | 6.3 | 54 | 36.2 | 3.5 |  |  |  |  |  |  |
| EMR | Iraq, MICS[56] | 2018 | 2,270 (7.4) | 6 | 84.3 | 1.3 | 8.4 | 128 (0.5) | 9.1 |  | 88.8 | 1.0 | 1.1 |
|  | Somalia, DHS[58] | 2020 | 14,651 (99.2) |  | 33.9<sup>¶</sup> | 64.2<sup>¶¶</sup> | 1.9 |  |  |  |  |  |  |
|  | Sudan, MICS [57] | 2014 | 15,853 (86.6) | 2.2 | 16.3 | 77 | 4.5 | 5,570 (31.5) |  |  |  |  |  |
|  | Yemen, DHS[33] | 2013 | 4,705(18.5) | 7 | 89.7 | NA | 3.3 | 1,909 (15.9)<sup>†</sup> | 10.7<sup>††</sup> | 88.3<sup>††</sup> |  |  | 1.0<sup>††</sup> |

Abbreviations: AFR: African Region; DHS: Demographic and Health Survey; EMR: Eastern Mediterranean Region; MICS: Multiple Indicator Cluster Surveys; WHO: World Health Organization.
<sup>§</sup>Year of data collection
§§ Percentages of types from women and girls are calculated from the total number of women and girls with FGM/C, respectively
¶ Type I, Sunni= 21.6% and Type II, Intermediate = 12.3%
¶¶ Type III and IV= 64.2%
† Prevalence and total number with FGM/C correspond to those of women with at least one daughter with FGM/C.
†† Percent distribution of most recent daughters who had FGM/C

In all countries, for the majority of women and girls, FGM/C was performed by traditional circumcisers, whilst a lower proportion was performed by medical professionals. The exception was girls in Egypt, where the proportion of FGM/C performed by medical professionals was 81.9% (Table 4) [55]. For women, in all countries where age of FGM/C was reported, FGM/C was most commonly performed at early ages (0-5 years) except for Kenya, Egypt, Sierra Leone, Guinea, and Tanzania where the procedure was most commonly done at 9-14 years, and Somalia where it was most commonly done at 5-9 years. For girls, the highest proportion of FGM/C was performed at the lowest age category: under 1 year of age (seven countries). Exceptions include Burkina Faso, Gambia, and Tanzania where the category 1-4 years had higher proportions, Sierra Leone, Kenya, and Guinea (most commonly done at 5-9 years), Egypt (most commonly done at 11-12 years) and Somalia (most commonly done at 10-14 years).

**Table 4.** Characteristics of Female Genital Mutilation/Cutting (FGM/C) Procedure in Nationally Representative Studies

| WHO Region | Percent distribution of women 15-49 years by: |  |  |  | Percent distribution of girls 0-14 years by: |  |
| --- | --- | --- | --- | --- | --- | --- |
|  | Country, Survey | Year<sup>s</sup> | Age at FGM/C (%) | Performer of FGM (%) | Age at FGM/C (%) | Performer of FGM (%) |
| AFR | Burkina Faso, DHS[35] | 2010 | <5y (60.4%), 5-9y (28.2%), 10-14y (8.9%), 15+y (2.0%), Don't know/missing (0.5%) | Traditional (97.2%)<br>Medical (0.2%)<br>Don't know/missing (2.6%) | <1y (2.8%), 1-4y (7.3%), 5-9y (3.1%), 10-14y (0.1%), Don't know/missing (0.1%) | Traditional (98.3%)<br>Medical (0.2%)<br>Don't know/missing (1.5%) |
|  | Eritrea, Population and Health Survey [32] | 2010 | <1y (47.4%), 1-3y (4.7%) | Traditional (84.4%)<br>Medical (0.3%)<br>Other (15.4%) | <1y (65.7%), 1-4y (20.9%), 5-6y (8.1%), 7-8y (4.3%), 9-10y (0.4%), 11-12y (0.1%)<br>13+y (0.1%), Don't know/missing (0.4%) | Traditional (98.3%)<br>Medical (0.09%)<br>Don't know/missing (1.6%) |
|  | Ethiopia, DHS [39] | 2016 | <5y (48.6%), 5-9y (21.7%), 10-14y (18.0%), 15+y (5.9%), Don't know/missing (5.8%) | Traditional (90.1%)<br>Medical (1%)<br>Don't know/missing (8.9%) | <1y (7.2%), 1-4y (3.4%), 5-9y (3.7%), 10-14y (1.0%), Don't know/missing (0.3%) | Traditional (97.6%)<br>Medical (1.9%)<br>Don't know/missing (0.5%) |
|  | Kenya, DHS [44] | 2014 | <5y (2.3%), 5-9y (26.6%), 10-14y (42.6%), 15+y (26.9%), Don't know /missing (1.7%) | Traditional (83.3%)<br>Medical (14.8%)<br>Don't know/missing (1.9%) | <1y (0.0%), 1-4y (0.2%)<br>5-9y (2.1%), 10-14y (0.5%) | Traditional (74.9%),<br>Medical (19.7%)<br>Don't know/missing (5.4%). |
|  | Mali, DHS [46] | 2018 | <5y (75.5%), 5-9y (16.1%), 10-14y (4.4%). 15+y (0.3%), Don't know missing (3.6%) | Traditional (91.5%)<br>Medical (0.3%)<br>Don't know/missing (8.2%) | <1y (34.2%), 1-4y (31.9%), 5-9 (5.2%), 10-14y (0.4%), Don't know/missing (0.9%) | Traditional (98.6%)<br>Medical (1.4%) |
|  | Nigeria, DHS [48] | 2018 | <5y (85.6%), 5-9y (4.2%), 10-14y (3.9%), 15+y (4.5%), Don't know/missing (1.8%) | Traditional (85.4%)<br>Medical (8.6%)<br>Don't know/missing (6%) | <1y (17.2%), 1-4y (1.1%), 5-9 (0.7%), 10-14y (0.0%), Don't know/missing (0.1%) | Traditional (92.8%)<br>Medical (7%)<br>Don't know/missing (0.1%) |
|  | Niger, DHS[49] | 2012 | <5y (75.7%), 5-9y (7.3%), 10-14y (7.9%), 15+y (1.4%), Don't know/missing (7.8%) | Traditional (95.8%)<br>Other (0.2%)<br>Don't know/missing (4%) |  |  |
|  | Senegal, DHS [50] | 2019 | <5y (84.9%), 5-9y (10.4%), 10-14y (2.7%), 15+y (0.4%), Don't know /missing (1.7%) | Traditional (100%) | <1y (9.8%), 1-4y (5.4%), 5-9y (0.8%), 10-14y (0.0%), Don't know/missing (0.1) | Traditional (100%) |
|  | Sierra Leone, DHS [51] | 2019 | <5y (12.3%), 5-9y (14.1%), 10-14y (44.9%), 15+y (26.1%), Don't know /missing (2.5%) | Traditional (98.4%)<br>Medical (0.4%)<br>Don't know/missing (1.2%) | < 1y (0.0%), 1-4y (0.6%), 5-9y (4.1%), 10-14y (3.1%), Don't know/missing (0.1%) | Traditional (99.4%)<br>Medical (0.6%) |
|  | Guinea, DHS[42] | 2018 | <5y (22.4%), 5-9y (36.7%), 10-14y (28.4%), 15+y (3.9%), Don't know/missing (8.6%) | Traditional (77.6%)<br>Medical (17.3%)<br>Don't know/missing (5.1%) | <1y (1.5%), 1-4y (11.9%), 5-9y (22.7%), 10-14y (2.3%), Don't know/missing (0.8%) | Traditional (64.8%)<br>Medical (34.9%)<br>Don't know/missing (0.3%) |
|  | Liberia, DHS[45] | 2020 | <5y (24.6%), 5-9y (16.7%), 10-14y (33%), 15+y (21.6%), Don't know (4.1%) |  |  |  |
|  | Gambia, DHS[40] | 2021 | <5y (64.9%), 5-9y (17.7%), 10-14y (6%), 15+y (0.7%), Don't | Traditional (95.1%)<br>Medical (0.4%) | <1y (21.9%), 1-4y (19.4%), 5-9y (3.9%), 10-14y (0.2%), Don't know | Traditional (98.8%)<br>Medical (0.1%), |

|  |  |  | know /missing (10.6%) | Don't know/missing (4.5%) | (4.5%) | Don't know/missing (1.1%) |
| --- | --- | --- | --- | --- | --- | --- |
|  | Tanzania, DHS[52] | 2016 | <1y (35.4%), 1-4y (2.3%), 5-6y (5.2%), 7-8y (7.5%), 9-10y (10.9%), 11-12y (9.3%), 13+y (27.6%), Don't know/missing (1.8%) | Traditional (86%)<br>Medical (14.8%)<br>Don't know/missing (1.9%) | <1y (0.1%), 1-4y (0.2%), 5-9y (0.1%), 10-14y (0.1%) | Traditional (74.9%)<br>Medical (19.7%)<br>Don't know/missing (5.4%) |
| EMR | Egypt, DHS[55] | 2014 | <3y (0.6%), 3-4y (1%), 5-6y (7.4%), 7-8y (13.4%), 9-10y (40.9%) 11-12y (24.6%), 13-14y (5.3%), 15-17y (2.6%), 18-19y (0.1%), Don't know/missing (4.2%) | Traditional (60.5%)<br>Medical (37.9%)<br>Other (0.1%),<br>Don't know/missing (1.5%) | <3 y (3.5%); 3-4y (3.4%), 5-6y (10.1%), 7-8y (14.1%), 9-10y (32.8%) 11-12y (28.6%), 13-14y (5.4%), 15-17y (1.3%), Don't know /missing (0.7%) | Medical (81.9%)<br>Traditional (18.1%)<br>Don't know/missing (0.3%) |
|  | Somalia, DHS [58] | 2020 | <5y (0.2%), 5-9y (70.9%), 10-14y (27.7%), 15+y (0.7%), Don't know /missing (0.5%) |  | 0-4y (3.3%), 5-9y (29.7%), 10-14y (75.9%) |  |
|  | Yemen, DHS [33] | 2013 | First week after birth (83.8%), after first week but before first year (10.5%), >=1y (1.2%), Don't know /missing (4.5%) | Traditional (92.8%)<br>Medical (2.9%)<br>Don't know/missing (4.3%) | First week after birth (84.9%), after 1st week but before 1 year (14.3%), >=1y (0.6%), Don't know/missing (0.2%) | Traditional (84.7%)<br>Medical (12.8%)<br>Don't know/missing (2.5%) |
| SEAR | Indonesia, RISKESDAS [30] | 2013 |  |  | 1-5 months (72.4%), 1-4y (13.9%), 5-11 y (3.3%) |  |
|  | Maldives, DHS[149] | 2016-2017 | <5y (83.1%), 5-9y (1.6%), 10-14y (0.4%), 15+y (0.4%), Don't know /missing (14.5%) |  | <1y (0.7%), 1-4y (0.2%), Don't know/missing (0.1%) |  |

Abbreviations: AFR: African Region; DHS: Demographic and Health Survey; EMR: Eastern Mediterranean Region; MICS: Multiple Indicator Cluster Surveys; WHO: World Health Organization; y: years.
<sup>s</sup>Year of data collection

### Sub-regional studies

Thirty-three sub-regional studies were from 13 countries, with ten from EMR and 23 from AFR. Among studies including women, the highest FGM/C prevalence was in Somaliland, Somalia (99.1%) [60] and the lowest was in Axum Town, North Ethiopia (0.7%) [61]. Regarding the 17 sub-regional studies including girls, the highest FGM/C prevalence was in Kersa, Ethiopia (88.1%) [62] and the lowest was in Axum Town, Ethiopia (0%) [61] (S5 Results, S6 Table). Eight out of the 33 sub-regional studies reported on FGM/C type. Type IV was most common in one study [63], Type II was the most common in four studies [64–67] and “sewn closed” was the most common in two sub-regional DHS reports on Somaliland [60] and the Northeast Zone of Somalia [68] (S5 Results, S7 Table). In 12 studies, the most common performers of FGM/C were traditional circumcisers [62, 63, 66, 69–77]. In three studies, in Egypt [78, 79] and Saudi Arabia [80] medical professionals were more common (S5 Results, S8 Table).

### School, Community or Facility based studies excluding studies on migrant populations

Within 98 non population-based studies, 54 studies (excluding studies on migrant populations) were from 15 countries, with 30 studies from countries in AFR, three studies from Malaysia in SEAR, and 21 studies from countries in EMR (S6 Results, S9 Table). Thirty-one were hospital/clinic-based, 14 school-based, and nine community-based studies. School and university-based studies reported a prevalence ranging from 9.4% [81] to 83.3% [82]; hospital or clinic-based studies reported a prevalence from 13% [83] to 100% [84], and community-based studies reported a prevalence from 0.4% [85] to 99.3% [86] (S6 Results, S10 Table). Two had prospective designs, two were retrospective, one was a cohort study, and 49 were cross-sectional. Twenty-five studies reported on FGM/C types. In ten studies Type I was most common [83, 87–95], Type II was most common in four studies [96–99], Type III in three studies [100–102], and Type IV in two studies [86, 103] (S6 Results, S11 Table).

### Studies on migrant populations

Within the 98 non population-based studies, 44 studies on migrant populations with FGM/C were identified. The included studies were from the Region of the Americas (AMR) (9 studies), European Region (EUR) (25 studies), Western Pacific Region (WPR) (5 studies), and EMR (5 studies) (S7 Results, S13 Table). Most studies had a moderate risk of bias and four had a high risk of bias. Participants in these studies were categorized as migrants, refugees, or asylum seekers. Study designs were case control (n=1), and randomised controlled trial (n=1), population based (n=5), retrospective or database studies (n=5), and cross-sectional studies (n=15), case series (n=17). Prevalence within these migrant populations ranged from 0.32% (of a sample of 145,492) [104] to 99% (of a sample of 191) [105] (S8 Results, S14 Table). Type III [104–113] was the most common type in 10 studies, followed by Type II in 9 studies [114–122], Type I (8 studies) [123–130], and Type IV (three studies) [131–133] (S7 Results, S15 Table).

## Discussion

This systematic review and meta-analysis estimated that nearly 100 million girls and women of reproductive age had FGM/C, which was among countries included in the analysis. Results indicated that the practice remains widespread in countries where it is reported. In particular, across 28 countries there was a pooled prevalence of 38% among women aged 15-49 years old, and across 23 countries, there was a pooled prevalence of 7% among girls aged 0-14 years old. Over repeated cross-sectional surveys, the prevalence of FGM/C appears to have decreased in 23 countries for women and 25 countries for girls. It appears to have increased in three countries for women (Guinea-Bissau, Burkina Faso, and Somalia) and two countries for girls (Djibouti and Cameroon). For both women and girls who had FGM, most had the type “flesh removed” (Types I and II), and “sewn closed” (Type III), the most severe type of FMG/C, was practised over three-quarters of women and girls in Sudan and Sierra Leone. In most countries, FGM/C commonly occurred in early childhood and was performed by traditional circumcisers. FGM/C appears to continue in those who migrate from countries where FGM/C is prevalent.

The total prevalence of FGM/C specified in this study is consistent with previous estimates of FGM/C among girls and women of reproductive age where estimates of FGM/C range from 100-140 million women and girls [2, 3]. Our study findings differ to the most recent UNICEF report, which states the global prevalence of FGM/C to be over 200 million among living women and girls; although the upper end of the combined confidence interval was close to this estimate [1, 4]. UNICEF extrapolated their prevalence to women of all ages and this study was unable to locate reports to provide an estimate for women from Djibouti, women and girls from Indonesia and this study excluded estimates from surveys that used a household level prevalence of FGM/C among girls.

The decline of FGM/C across repeated cross-sectional studies in many countries is encouraging and corresponds with previous research, which showed an absolute decline in the prevalence of FGM/C amongst girls aged 0-14 years by 51.8%; from 67.6% in 1990-1996 to 15.8% in 2015-2017 [21]. Results were consistent with previous research regarding large variations in prevalence between countries and regions [5, 21, 134].

Structural level changes including legislative bans and policy changes are likely to play a role in the possible decline. Globally, there are 84 countries that either have specific legislation that bans FGM/C or other legislation that enables the persecution of FGM/C [135, 136]. In Egypt, the lower prevalence for girls may relate to the legal ban implemented in 2008 [55]. However, the efficacy of laws against FGM/C depend on enforcement and the specificities of the law. For example, in Liberia and Mauritania laws only protect girls below the age of 18 [136, 137] and in Indonesia, FGM/C was legalized in a medical setting in 2010, however, the repeal of that law in 2014 left no explicit ban or consequences [136, 138]. In Somalia, there is no national legislation that enforces the Somalia constitution which states that “circumcision is prohibited” [135, 136]. Furthermore, there is no legislative ban in Mali and the prevalence remains high at 88.6% of women and 72.7% of girls [46].

In addition to legislation and judicial enforcement, other mechanisms may have contributed to a reduction in FGM/C, such as education, literacy and change in social norms [139, 140]. To end the propagation of FGM/C future research should undertake process evaluations of structural, community and family level interventions and policies in countries where FGM/C has declined. Understanding the underlying mechanisms for change in FGM/C, in countries where there has been success, will be instrumental for the adoption of effective policies and interventions to meet the SDG target 5.3.

Consistent with other studies, the most common FGM/C type amongst women and girls was ‘cut with flesh removed’, equivalent to Type I or II [5, 141]. Koski and colleagues reported that there were no significant differences regarding the types and severity of FGM/C across cohorts [5].

Similar to other findings, this review found that FGM/C most often occurs in early childhood [141]. Similar to the findings of this study, UNICEF reported that traditional circumcisers perform most procedures. Yet, the opposite occurs in Egypt where medicalization of FGM/C was high despite its ban [55]. WHO and UNICEF have called for the end of medicalization of FGM/C [142, 143]. Discussions around the medicalisation of FGM/C are beyond the scope of this study but this has been discussed elsewhere [142, 144, 145].

Studies of different regions or facilities in the same countries had different prevalence reports, a phenomenon also reported by UNICEF [141], likely owing to regional or community risk factors. For example, the national prevalence in Ethiopian women was 65.2% [39], while in one region, the East Gojjam Zone, it was 96% [72]. Studies based on migrant populations have widely varying prevalence estimates. They demonstrate that FGM/C is present in countries where it is not traditionally practiced; however, high quality studies are needed to understand FGM/C in these countries, and to inform policies, interventions, and relevant healthcare services.

The strengths of the study ensure a thorough and accurate examination of the research question. The review had broad inclusion criteria to provide a comprehensive review of all FGM/C studies. The study used robust methods to identify studies, extract data, and present findings. The broadest possible scope of research was scanned with no restrictions on language. A hand search of grey literature was conducted to be as comprehensive as possible. Moreover, DHS and MICS data, which are collected via representative sampling methodology with high response rates and a low risk of bias, ensured the quality of the meta-analyses.

This study had several limitations. Estimates were based on the available published data, which may not reflect the actual global prevalence of FGM/C. There were two missing country reports unavailable for analysis (S1 Methods and Results, S1 Text). The actual global total number of girls and women with FGM/C will be higher than that reported in this study due to missing data from key countries. For example, Indonesia was not included in the meta-analysis due to lack of a denominator. FGM/C was self-reported, thus the prevalence estimates may be underreported due to legal ramifications or social desirability. Furthermore, the translation of terms within surveys may impact recall and comprehension, which emphasizes the need for survey tools to be validated within each context. In addition, women and girls may not be able to accurately recall the type of procedure performed on them, or there may be confusion due to multiple ways of describing each type [146]. Furthermore, recollection of who performed the procedure may be inaccurate [147].

The prevalence in the 0-14 age group may be underreported as these girls are still at risk of FGM/C at the time of survey. Future research should adjust prevalence by age at FGM/C procedure or conduct analyses based on age cohorts to be inclusive of those still at risk of FGM/C. A future study examining FGM/C prevalence among five-year age cohorts will be useful to understand if trends exist across age groups [141]. This study also shows the need for consistency in future research regarding the denominator of FGM/C among girls and terminology used to describe each type of FGM/C.

This study highlights the need to expand data collection and surveillance using robust methodologies particularly in high resource countries with migrant populations from countries that practice FGM/C. There are numerous data gaps on the national prevalence of FGM/C in multiple countries, including: Colombia, Georgia, Russia, Iran, Oman, Kuwait, Singapore, Thailand, the Philippines, India, Pakistan, Ecuador, Peru, Saudi Arabia, the State of Palestine, Sri Lanka and United Arab Emirates [148]. In Indonesia approximately 50% of girls aged 0-14 had FGM/C; however, we know relatively little about FGM/C in Indonesia, which warrants further investigation given its large population size.

In conclusion, approximately 100 million women and girls have had FGM/C among countries included in the analysis, and there is large variation between countries in progress to ending FGM/C by 2030. Current findings may be used as a baseline in future attempts to track progress to meeting SDG 5.3. A decline to end FGM/C across future generation of girls may be possible in the near future in low-prevalence countries such as Niger, Uganda, and Ghana. However, the decline in FGM/C must be greater in countries where the current prevalence of FGM/C is higher such as Egypt, Sudan, Indonesia, Somalia, Djibouti, Guinea, and Mali, which emphasizes the need for immediate interventions and policies to end this harmful practice.

## Data sharing statement

All data generated or analysed for this study are included in this article and its supplementary files.

## Contributors

SM conceived the study and SA, C Akl, and C Akik contributed to the study design. LF and SM wrote the protocol with contributions from SA, C Akl, C Akik. LF completed the literature search. LF and ZD selected the studies and extracted relevant data. LF analysed the data and wrote the first draft of the paper. SM, C Akl, C Akik, SA, and ZD revised drafts and approved the final manuscript. SM supervised LF, and SM is the guarantor of the study.

## Supporting information

S1 Methods and Results

S2 Checklist

S3 Protocol

S4 Results

S5 Results

S6 Results

S7 Results

## Data Availability

All data produced in the present work are contained in the manuscript

## Acknowledgements

We thank Dr Marie-Claire Rebeiz, Mrs Tanya Khoury, and Ms Sara Mansour who verified the data in the tables.

## Supporting information

### Captions for Figures

**Figure 2.** Footnote: There were 30 studies included in the systematic review as nationally-representative studies, however, The Pew Research Study [31] did not include women and the Indonesia RISKESDAS [30] did not report sample sizes, thus they were not included in this analysis.

**Figure 3.** Footnote: There were 30 studies included in the systematic review as nationally-representative studies, however, surveys from Liberia [45], Niger [49], and Uganda [54] did not include girls, and The Pew Research Study [31], Yemen [33], and Eritrea [32] only included women who reported on at least one daughter in their household who has had FGM/C, and the Indonesia RISKESDAS [30] did not report sample sizes, thus they were not included in this analysis.

**S1 Methods and Results**

S1 Table. Search Strategy

S2 Table. Inter-rater reliability rate at different stages of the screening process.

Footnote: A third reviewer confirmed the inclusion of all studies. The Cohen’s kappa provided a global score across all three inclusion criteria; after the full text screening it was decided that the risk factors of FGM/C would be presented in separate paper. At stage 1, reviewers had the option to indicate if they were unsure, which may also partially explain the low score before resolution. Agreement was higher on the first two points of the inclusion criteria: (i) prevalence studies and (ii) non population-based studies examining FGM/C.

S1 Text. Inclusion and Exclusion Criteria.

S2 Text. Supplementary results.

**S2 Preferred Reporting Items for Systematic Reviews and Meta-Analyses (PRISMA) Checklist**

**S3 Study Protocol**

S1 Fig. Funnel plot of FGM/C prevalence in Women of Reproductive Age (15-49 years old) in Nationally Representative Studies.

S2 Fig. Funnel plot of FGM/C prevalence in Girls (0-14 years old) in Nationally Representative Studies.

**S4 Results: Nationally representative studies**

S4 Table. Characteristics of nationally representative studies.

Footnote: Abbreviations: AFR: African Region; DHS: Demographic and Health Survey; EMR: Eastern Mediterranean Region; MICS: Multiple Indicator Cluster Surveys; SEAR: South-East Asia Region; WHO: World Health Organization.

Legend*Not included in meta-analysis.

**S5 Results: Sub-Regional Population-Based Studies**

S5 Table. Characteristics of Sub-Regional Population-Based Studies.

Legend: *Patient report and examination, all others: Patient Report † women reported that at least 1 daughter had FGM/C in the household.

Footnote: All studies used cross-sectional methods.

S6 Table. Prevalence of FGM/C in Women and Girls in Sub-Regional Population-Based Studies. Footnote: Abbreviations: AFR: African Region EMR: Eastern Mediterranean Region, FGM/C: Female Genital Mutilation/Cutting.

Legend: * Women reported that at least 1 daughter had FGM/C in the household. † Youngest daughter had FGM/C. ‡ Due to inconsistent data reported in the study, this number was calculated by the authors of this review.

S7 Table. Types of FGM/C in Sub-Regional Population-Based Studies.

Legend * % of Women † % of youngest daughter ‡ % of girls

Footnote: Somaliland and Northeast Zone MICS calculate the prevalence of types out of the total number of participants, and report type II as “flesh removed” and type II as “sewn closed”.

Abberviations FGM/C: Female Genital Mutilation/Cutting

S8 Table. Characteristics of FGM/C Procedure in Sub-Regional Population-Based Studies.

Footnote Abbreviations: AFR: African Region EMR: Eastern Mediterranean Region Female Genital Mutilation/Cutting

**S6 Results: School, Community or Facility based studies excluding studies on migrant populations**

S9 Table. Characteristics of School, Community or Facility based studies excluding studies on migrant populations.

Legend: **\***Types of FGM/C mentioned were: Clitoral tip excision, Complete clitoridectomy, Clitoridectomy/labia minora Excision, Clitoridectomy/labia minora/Inner majora excision.

Footnote: Abbreviations: AFR: African Region EMR: Eastern Mediterranean Region SEAR: South East Asian Region

S10 Table. Proportion of FGM/C in Women and Girls in School, Community or Facility based studies excluding studies on migrant populations.

Legend: *Out of the female school teachers **Without excluding those who were unsure if they had been mutilated.*** Prevalence according to clinical examination.

Footnote: Abbreviations: AFR: African Region EMR: Eastern Mediterranean Region, SEAR: South East Asian Region, FGM/C: Female Genital Mutilation/Cutting

S11 Table. Types of FGM/C in School, Community or Facility based studies excluding studies on migrant populations.

Footnote: Abbreviations: AFR: African Region EMR: Eastern Mediterranean Region SEAR: South East Asian Region, FGM/C: Female Genital Mutilation/Cutting

S12 Table. Characteristics of FGM/C Procedure in School, Community or Facility based studies excluding studies on migrant populations.

Footnote: Abbreviations: AFR: African Region EMR: Eastern Mediterranean Region SEAR: South East Asian Region FGM/C: Female Genital Mutilation/Cutting

**S7 Results: Studies on Migrant Populations.**

S13 Table. Characteristics of Studies on Migrant Populations.

Legend: * Tissue removed and sewn closed, tissue removed and some stitching, some tissue removed, pricking. † Flesh removed, Genital area just nicked, Genital area sewn closed

Footnote: Abbreviations: EMR: Eastern Mediterranean Region. SEAR: South East Asian Region. EUR: European Region. WPR: Western Pacific Region AMR: American Region

S14 Table. Prevalence of FGM/C in Migrant Populations.

Footnote: Abbreviations: EMR: Eastern Mediterranean Region. SEAR: South East Asian Region. EUR: European Region. WPR: Western Pacific Region AMR: American Region FGM/C: Female Genital Mutilation/Cutting

S15 Table. Types of FGM/C in Migrant Populations.

Legend: *Flesh removed and some stitching † Flesh removed and sown closed. Footnote: Abbreviations: EMR: Eastern Mediterranean Region. SEAR: South East Asian Region. EUR: European Region. WPR: Western Pacific Region AMR: American Region

FGM/C: Female Genital Mutilation/Cutting

S16 Table. Characteristics of FGM/C Procedure for Migrant Populations.

Footnote: Abbreviations: EMR: Eastern Mediterranean Region. SEAR: South East Asian Region. EUR: European Region. WPR: Western Pacific Region AMR: American Region FGM/C: Female Genital Mutilation/Cutting

---

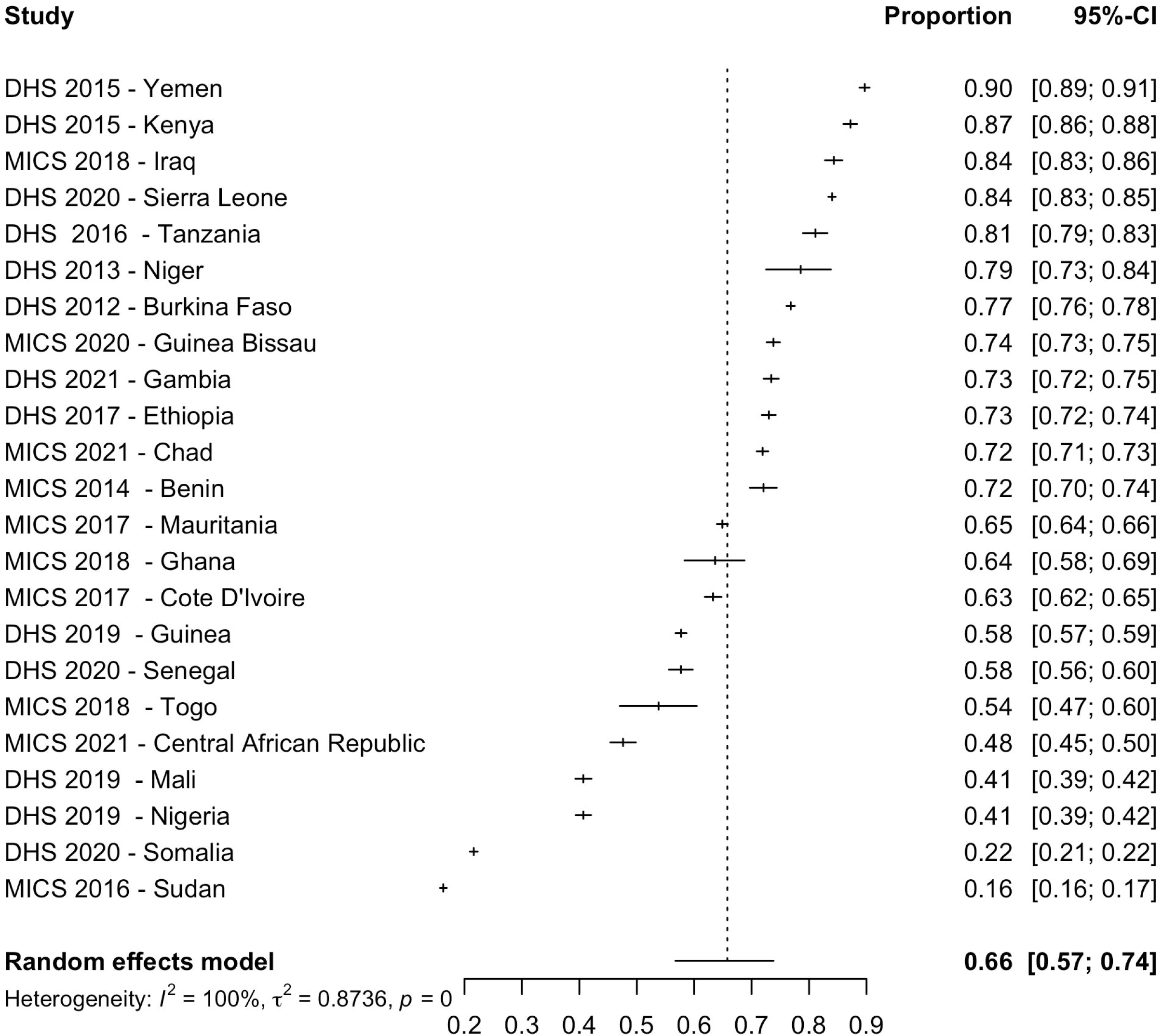

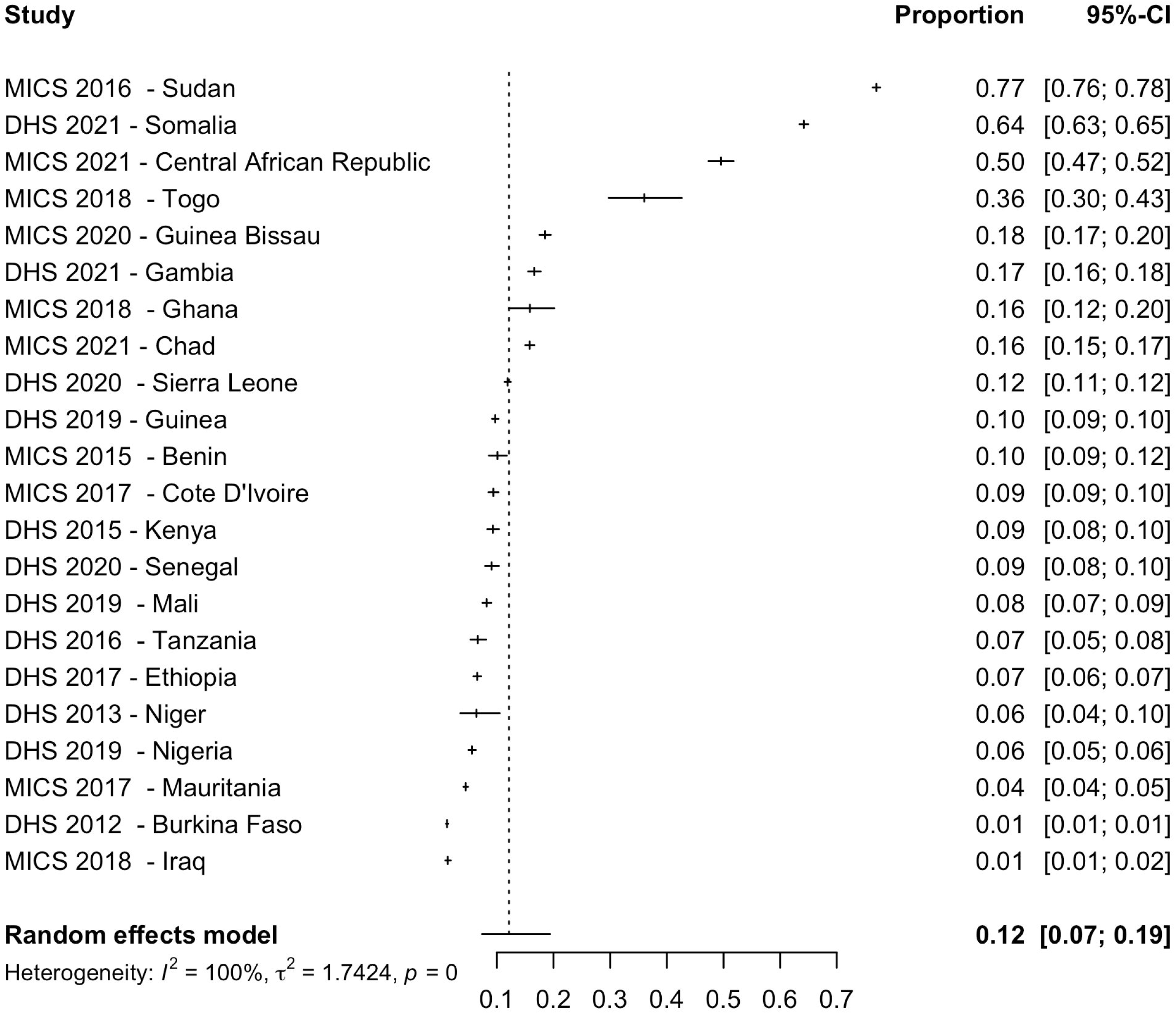

