## Supplementary material for "The global prevalence of female genital mutilation/cutting: A systematic review and meta-analysis of national, regional, facility and school-based studies": S1 Methods and Results

**S1 Table. Search strategy**

| **Database** | **Search Terms** |
| --- | --- |
| PsycINFO | 1 Female Genital Mutilation;2 Female Circumcision;3 Female Genital Cutting; 4 Female Genital Alteration; 5 or/1-4; 6 Limit 5 by 2009-2022 |
| PubMed | (("Female Genital Alteration") OR ("Female Genital Cutting") OR ("Female Circumcision") OR ("Female Genital Mutilation")) 2009:2022 [dp] |
| Embase | ('female genital mutilation'/exp OR 'female genital mutilation' OR (('female' OR 'female'/exp OR female) AND genital AND ('mutilation' OR 'mutilation'/exp OR mutilation))) AND [2009-2022]/py |
| Ovid MEDLINE(R) and In-Process, In-Data-Review & Other Non-Indexed Citations  <1946 to March 23, 2022> | 1 Female Genital Mutilation.mp.  2 Female Circumcision.mp.  3 Female Genital Cutting.mp.  4 Female Genital Alteration.mp.  5 1 or 2 or 3 or 4 (1831)  6 Circumcision, Female/ (1569)  7 5 or 6 (2208)  8 ((female adj3 circumcis*) or (girl adj3 circumcis*) or (wom?n adj3 circumcis*)).mp.  9 ((female adj3 genital* adj3 cut*) or (girl adj3 genital* adj3 cut*) or (wom?n adj3 genital* adj3 cut*)).mp.  10 ((female adj3 genital* adj3 alteration*) or (girl adj3 genital* adj3 alteration*) or (wom?n adj3 genital* adj3 alteration*)).mp.  11 ((female adj3 genital* adj3 mutilation) or (girl adj3 genital* adj3 mutilation) or (wom?n adj3 genital* adj3 mutilation)).mp.  12 ((female adj3 genital* adj3 alter) or (girl adj3 genital* adj3 alter) or (wom?n adj3 genital* adj3 alter)).mp.  13 8 or 9 or 10 or 11 or 12 (2358)  14 5 or 7 or 13 (2358)  15 limit 14 to yr="2009 -Current" (1364) |
| Web of Science | 1 Female Genital Mutilation; 2 Female Circumcision;3 Female Genital Cutting; 4 Female Genital Alteration, 5 or/1-4; 6 Limit 5 by 2009-2022 & Document Type: Articles |

**S2 Table. Detailed inter-rate reliability at different stages of the screening process. [1]**

|  | **Stage of systematic review** | Cohen’s Kappa |
| --- | --- | --- |
| Stage 1 | Agreement after initial screening of titles and abstracts  *Coding decisions and conflicts were discussed and a third reviewer (experienced systematic reviewer) assisted in decision making* | 0.34 |
| Stage 1 | Agreement after completing conflict resolution and consensus | 0.95 |
| Stage 2 | Agreement after initial screening of full text  *Coding decisions and conflicts were discussed to reach consensus and a third reviewer (experienced systematic reviewer) assisted in decision making.* | 0.59 |
| Stage 2 | Agreement after completing conflict resolution and consensus | 1.0 |

A third reviewer confirmed the inclusion of all studies. The Cohen’s kappa provided a global score across all three inclusion criteria; after the full text screening it was decided that the risk factors of FGM/C would be presented in separate paper. At stage 1, reviewers had the option to indicate if they were unsure, which may also partially explain the low score before resolution. Agreement was higher on the first two points of the inclusion criteria: (i) prevalence studies and (ii) non-population based studies examining FGM/C.

**S1 Text. Inclusion and Exclusion Criteria**

Inclusion criteria:

(i) Reported on FGM/C prevalence using population-based methods (cross-sectional or cohort studies) at the national or subnational level or examined FGM/C in facility
(ii) **or** non-population based studies examining FGM/C which may include case series, regional studies, facility-based studies or school-based studies.
(iii) **or** examined the risk factors of FGM/C within a population (cross-sectional, case-control or cohort studies). To meet the last criterion the comparison had to occur between women or girls with FGM/C to those without FGM.

Exclusion criteria:

(i) only reported health impacts of FGM/C, healthcare providers, policy, economic effects, or perceptions,
(ii) only used qualitative methods
(iii) were systematic reviews (except for the purpose of referencing)
(iv) conference proceedings and letters to the editor.

**Excluded Studies**

These exclusions are relevant to the accuracy of the pooled prevalence estimate. In Cameroon, the 2014 MICS does not report FGM/C prevalence, therefore it was excluded from this analysis. It is important to note that UNICEF Data reports the prevalence of FGM/C in Cameroon as 1% which is very low and indicates near eradication of the procedure. Djibouti 2012 EDSF/PAPFAM reported high prevalence of FGM, 43% amongst girls alone and 94% amongst women and girls [2]. However, it was also excluded from this analysis because the original report could not be located. The original report of the Zambia Sexual Behaviors Survey 2009 could not be located as well, and was therefore excluded. It reports a 0.7% national prevalence (women and girls together) [3]. Finally, the Indonesian 2013 RISKESDAS did not provide sample size and was excluded from the meta-analysis [4].

**S2 Text. Supplementary results.**

**Nationally Representative Studies.**

Egger’s [151] test results showed P=0.93, and this did not indicate funnel plot asymmetry or bias among nationally representative surveys on the prevalence of FGM/C amongst women (Supplementary Figure 1). Egger’s test results showed P=0.013, indicating funnel plot asymmetry and implying bias among nationally representative surveys on the prevalence of FGM/C amongst girls (Supplementary Figure 2). In addition, the funnel plots visually show high heterogeneity, although it is likely that this is due to different prevalence rates in different countries rather than publication bias.

**Studies with Non-population-based Methods**

In sub-region-based studies that did not discriminate between women and girls, the highest prevalence rate was recorded at 99.7% in clinics in Somalia [5]. Nigeria (16 studies), followed by Egypt (9 studies) accounted for the largest contribution of studies. In SEAR studies ranged from 88% [6] to 99.3% [7]. In EMR, the range was between 14.7% [8] and 99.7% [5]. In AFR, the range was 0.4% [9] to 100% [10]. The most common types were Type I in EMR and AFR and Type IV in SEAR (Malaysia only). In 24 studies where FGM/C performer was reported, FGM/C was more commonly performed by traditional circumcisers than medical performers (18 studies reporting majority performed by traditional circumcisers, 6 studies reporting majority performed by medically trained circumcisers all of which were based in Egypt and one in Nigeria). In addition, the procedure was more commonly occurred more commonly at home than at a clinic.

**Studies on Migrant Populations.**

Most studies on migrant populations used convenience or purposive samples with small sample sizes. In AMR, the prevalence ranged from 2.1% [11] to 100% [12] almost only in African migrants; EUR studies reported 0.54% [13] to 99% [14] among African, Central Asian, and Balkan migrants; EMR studies reported 9.4%[15] to 67.3%[16] among Sudanese and other unspecified migrants in Saudi Arabia. One non-case series study in WPR reported a prevalence of 0.71% [17] and 1.64% [18]. The most common type varied between studies. Where it was reported, there were more traditional than medical FGM/C performers except in three studies in England [19], Turkey [20], and Saudi Arabia [21]. Generally, published articles reporting FGM/C prevalence in immigrants and refugees had fallible designs and higher bias risks. Nonetheless, they do report actual numbers in host locations, and despite their sample sizes, this may be more accurate than estimates based on prevalence in countries of origin.
