## Supplementary material for "The global prevalence of female genital mutilation/cutting: A systematic review and meta-analysis of national, regional, facility and school-based studies": S2 Checklist

| **Section and Topic** | **Item #** | **Checklist item** | **Location where item is reported** |
| --- | --- | --- | --- |
| **TITLE** | | |  |
| Title | 1 | Identify the report as a systematic review. | Title, Cover page |
| **ABSTRACT** | | |  |
| Abstract | 2 | See the PRISMA 2020 for Abstracts checklist. | Abstract, Page 2 |
| **INTRODUCTION** | | |  |
| Rationale | 3 | Describe the rationale for the review in the context of existing knowledge. | Introduction, Paragraph 4 |
| Objectives | 4 | Provide an explicit statement of the objective(s) or question(s) the review addresses. | Introduction, Paragraph 4 |
| **METHODS** | | |  |
| Eligibility criteria | 5 | Specify the inclusion and exclusion criteria for the review and how studies were grouped for the syntheses. | Methods – Inclusion and exclusion criteria, Paragraph 1  Methods – Inclusion and exclusion criteria, Paragraph 2  Data analysis, Paragraph 1 |
| Information sources | 6 | Specify all databases, registers, websites, organisations, reference lists and other sources searched or consulted to identify studies. Specify the date when each source was last searched or consulted. | Methods - Search strategy & study selection, Paragraph 1 |
| Search strategy | 7 | Present the full search strategies for all databases, registers and websites, including any filters and limits used. | S1 Methods and Results – S1 Table. Search Strategy |
| Selection process | 8 | Specify the methods used to decide whether a study met the inclusion criteria of the review, including how many reviewers screened each record and each report retrieved, whether they worked independently, and if applicable, details of automation tools used in the process. | Methods - Study screening |
| Data collection process | 9 | Specify the methods used to collect data from reports, including how many reviewers collected data from each report, whether they worked independently, any processes for obtaining or confirming data from study investigators, and if applicable, details of automation tools used in the process. | Methods – Data extraction and quality assessment |
| Data items | 10a | List and define all outcomes for which data were sought. Specify whether all results that were compatible with each outcome domain in each study were sought (e.g. for all measures, time points, analyses), and if not, the methods used to decide which results to collect. | Methods – Data extraction and quality assessment |
|  | 10b | List and define all other variables for which data were sought (e.g. participant and intervention characteristics, funding sources). Describe any assumptions made about any missing or unclear information. | Methods – Data extraction and quality assessment |
| Study risk of bias assessment | 11 | Specify the methods used to assess risk of bias in the included studies, including details of the tool(s) used, how many reviewers assessed each study and whether they worked independently, and if applicable, details of automation tools used in the process. | Methods – Data extraction and quality assessment |
| Effect measures | 12 | Specify for each outcome the effect measure(s) (e.g. risk ratio, mean difference) used in the synthesis or presentation of results. | Methods – Data analysis, Paragraph 1 |
| Synthesis methods | 13a | Describe the processes used to decide which studies were eligible for each synthesis (e.g. tabulating the study intervention characteristics and comparing against the planned groups for each synthesis (item #5). | Methods- Data analysis, Paragraph 1 |
|  | 13b | Describe any methods required to prepare the data for presentation or synthesis, such as handling of missing summary statistics, or data conversions. | Methods- Data analysis |
|  | 13c | Describe any methods used to tabulate or visually display results of individual studies and syntheses. | Methods- Data analysis, Paragraph 1 |
|  | 13d | Describe any methods used to synthesize results and provide a rationale for the choice(s). If meta-analysis was performed, describe the model(s), method(s) to identify the presence and extent of statistical heterogeneity, and software package(s) used. | Methods- data analysis  “A random-effects meta-analysis was conducted due to high heterogeneity between studies. The pooled prevalence estimates were transformed using the Freeman-Tukey double arcsine method and presented as a pooled prevalence with 95% CIs^18^ using the ‘metaprop’ command within the Meta package, version 4.15-1.^19^ |
|  | 13e | Describe any methods used to explore possible causes of heterogeneity among study results (e.g. subgroup analysis, meta-regression). | Methods- Data analysis, Paragraph 2 |
|  | 13f | Describe any sensitivity analyses conducted to assess robustness of the synthesized results. | None undertaken |
| Reporting bias assessment | 14 | Describe any methods used to assess risk of bias due to missing results in a synthesis (arising from reporting biases). | Methods- Data analysis, Paragraph 2 |
| Certainty assessment | 15 | Describe any methods used to assess certainty (or confidence) in the body of evidence for an outcome. | Methods- Data analysis, Paragraph 2 |
| **RESULTS** | | |  |
| Study selection | 16a | Describe the results of the search and selection process, from the number of records identified in the search to the number of studies included in the review, ideally using a flow diagram. | Results - Paragraph 1, Figure 1 |
|  | 16b | Cite studies that might appear to meet the inclusion criteria, but which were excluded, and explain why they were excluded. | Results - Paragraph 1, Figure 1 |
| Study characteristics | 17 | Cite each included study and present its characteristics. | S4 Results Nationally representative studies – S4 Table  S5 Results Subregional population based studies – S5 Table  S6 Results School, Community or Facility based studies excluding studies on migrant populations - S9 Table  S7 Results Studies on migrant populations – S13 Table |
| Risk of bias in studies | 18 | Present assessments of risk of bias for each included study. | S4 Results Nationally representative studies – S4 Table  S5 Results Subregional population based studies – S5 Table  S6 Results School, Community or Facility based studies excluding studies on migrant populations - S9 Table  S7 Results Studies on migrant populations – S13 Table |
| Results of individual studies | 19 | For all outcomes, present, for each study: (a) summary statistics for each group (where appropriate) and (b) an effect estimate and its precision (e.g. confidence/credible interval), ideally using structured tables or plots. | Results Page 9-15, Tables 1-4, S6 Appendix Tables S4-15  Results Paragraph 1 to 10.  Tables 1-4.  S4 Results: Nationally representative studies  S5 Results: Sub-Regional Population-Based Studies  S6 Results: School, Community or Facility based studies excluding studies on migrant populations  S7 Results: Studies on Migrant Populations. |
| Results of syntheses | 20a | For each synthesis, briefly summarise the characteristics and risk of bias among contributing studies. | Results – Nationally Representative Studies, Paragraph 1 |
|  | 20b | Present results of all statistical syntheses conducted. If meta-analysis was done, present for each the summary estimate and its precision (e.g. confidence/credible interval) and measures of statistical heterogeneity. If comparing groups, describe the direction of the effect. | Abstract  Results – Nationally Representative Studies, Paragraph 2  Results – Nationally Representative Studies, Paragraph 3  Results – Nationally Representative Studies, Paragraph 5 |
|  | 20c | Present results of all investigations of possible causes of heterogeneity among study results. | Figure 1  Figure 2  Figure 3  Figure 4a  Figure 4b  Figure 4c  Results – Nationally Representative Studies, Paragraph 2  Results – Nationally Representative Studies, Paragraph 3 |
|  | 20d | Present results of all sensitivity analyses conducted to assess the robustness of the synthesized results. | None undertaken |
| Reporting biases | 21 | Present assessments of risk of bias due to missing results (arising from reporting biases) for each synthesis assessed. | S1 Figure  S2 Figure |
| Certainty of evidence | 22 | Present assessments of certainty (or confidence) in the body of evidence for each outcome assessed. | Abstract  Results – Nationally Representative Studies, Paragraph 2  Results – Nationally Representative Studies, Paragraph 3  Results – Nationally Representative Studies, Paragraph 5 |
| **DISCUSSION** | | |  |
| Discussion | 23a | Provide a general interpretation of the results in the context of other evidence. | Discussion, Paragraph 2  Discussion, Paragraph 3  Discussion, Paragraph 4  Discussion, Paragraph 5  Discussion, Paragraph 6  Discussion, Paragraph 7  Discussion, Paragraph 8 |
|  | 23b | Discuss any limitations of the evidence included in the review. | Discussion, Paragraph 8  Discussion, Paragraph 9  Discussion, Paragraph 10  Discussion, Paragraph 11 |
|  | 23c | Discuss any limitations of the review processes used. | Discussion, Paragraph 10  Discussion, Paragraph 11 |
|  | 23d | Discuss implications of the results for practice, policy, and future research. | Discussion, Paragraph 4  Discussion, Paragraph 5  Discussion, Paragraph 8  Discussion, Paragraph 11  Discussion, Paragraph 12 |
| **OTHER INFORMATION** | | |  |
| Registration and protocol | 24a | Provide registration information for the review, including register name and registration number, or state that the review was not registered. | Abstract  Methods – Study protocol, registration and reporting |
|  | 24b | Indicate where the review protocol can be accessed, or state that a protocol was not prepared. | Abstract  Methods – Study protocol, registration and reporting, Protocol amendments |
|  | 24c | Describe and explain any amendments to information provided at registration or in the protocol. | Protocol amendments |
| Support | 25 | Describe sources of financial or non-financial support for the review, and the role of the funders or sponsors in the review. | Abstract  Ethical approval and role of the funding source |
| Competing interests | 26 | Declare any competing interests of review authors. | Declaration of interests |
| Availability of data, code and other materials | 27 | Report which of the following are publicly available and where they can be found: template data collection forms; data extracted from included studies; data used for all analyses; analytic code; any other materials used in the review. | Data sharing statement |

*From:*  Page MJ, McKenzie JE, Bossuyt PM, Boutron I, Hoffmann TC, Mulrow CD, et al. The PRISMA 2020 statement: an updated guideline for reporting systematic reviews. BMJ 2021;372:n71. doi: 10.1136/bmj.n71

For more information, visit: <http://www.prisma-statement.org/>
