## Supplementary material for "The global prevalence of female genital mutilation/cutting: A systematic review and meta-analysis of national, regional, facility and school-based studies": S3 Protocol

**Protocol amendments:**

1. No language restrictions were imposed on included articles. Modification on *Page 7.*

After an initial screen of the titles and abstracts, it was decided by the review team to improve the reach and the quality of the systematic review by placing no language restrictions on the search results.

2. All pooled meta-analyses will be presented by age group, in particular, by girls aged 0-14 and women aged 15-49. Modification on *Page 11.*

After piloting the data extraction form, it became clear that several studies presented prevalence estimates for both women of reproductive age (15-49 years old) and girls (0-14 years old). To prevent analyses from being subject to a ‘cohort effect’, the review team decided that pooled prevalence estimates will be presented by age group, where possible.

3. Clarification of included risk factors/determinants. As stated in objective 2 and the unmodified data extraction section, any reported risk factor/determinant of FGM will be included in the systematic review. Minor clarification on *Page 8.*
4. The prevalence of FGM objective will be presented and published separately from the determinants of FGM objective.
5. Clarification of inclusion of studies using the same data source. The prevalence FGM objective will only include the original reports; while the determinants of FGM objective will include the most relevant and comprehensive study. Modification on *Page 6 & 7.*

As per the original protocol all studies using the same data sources will be included in the wider systematic review project. However, the inclusion of the most relevant study will differ between the two objectives.

*All changes from the initial protocol have been highlighted within the text.*

The initial protocol was registered on 22nd May 2020**.** Amendments to the protocol were uploaded onto the OSF project page (<https://osf.io/h54bu/>).

**Title:** The global prevalence, distribution and determinants of female genital mutilation: a protocol for a systematic review and meta-analysis

**Authors:** Leen Farouki^1^, Sawsan Abdulrahim^2^, Christelle Akl^1^, Chaza Akik^1^, Stephen McCall^1*^

*Corresponding author

**Affiliations:**

1. Center for Research on Population and Health, Faculty of Health Sciences, American University of Beirut, Lebanon

2. Department of Health Promotion and Community Health, Faculty of Health Sciences, American University of Beirut, Lebanon

**Acknowledgements:** Zeinab El Dirani and Sandra Kahwaji joined the review team.

### Introduction

Female Genital Mutilation (FGM), also referred to as female circumcision or female genital cutting (FGC), is a non-medical procedure that entails the total or partial removal of external female genitalia and other injuries to the female genital organs (1). The UN Sustainable Development Goal (SDG) target 5.3 on gender equality calls for ending all harmful practices, including FGM (2). FGM is most often performed on girls between infancy and adolescence. The procedure has traditionally been performed in non-medical settings under unsanitary conditions, posing large risks of harm (3). In some settings, FGM has undergone a process of medicalization, whereby it is increasingly conducted in a medical setting (4).

As FGM is illegal in 44 countries, including in 25 African countries (1), it is likely that current estimates of the prevalence of FGM are under-represented. While the true global prevalence of FGM is unknown, an estimate by UNICEF sets the number at 200 million women and girls across thirty countries in Africa, Asia, and the Middle East, as well as migrants from those countries who have settled elsewhere in Europe and North and South America (5). In African countries where it is practised, over three million girls are at risk of FGM every year (6). A better grasp of the prevalence of FGM is necessary to inform adequate medical interventions for women and girls who have complications due to the procedure.

Reasons for the persistence of FGM are multifactorial and fall under religious, social, and cultural categories. In many settings, there are cultural beliefs that the procedure creates better marriage prospects because it associates with morality, hygiene, and aesthetics (7). The practice is associated with controlling women’s sexuality, as it is thought to curb sexual urges, which would maintain virginity until marriage and decrease a woman’s demands within marriage (8). In addition, lower socioeconomic status is commonly associated with FGM (9); however, this varies by country (10).

FGM may result in both immediate and long term negative health outcomes. During the procedure, a woman or girl may endure severe pain or shock and may contract HIV, Hepatitis B or C; the procedure may also lead to septicemia and death (1, 11). Medium to long term complications includes psychological, gynaecological, and obstetric consequences (12). Gynaecological problems include scarring, menstrual difficulties, infertility, and urinary problems. Psychological concerns include depression, anxiety, post-traumatic stress disorder, and memory problems. Obstetric consequences involve complications during pregnancy, haemorrhage and stillbirth (13). Moreover, FGM poses an economic burden for low- and middle-income countries as the global health care cost of addressing its complications amounts to 1.4 billion USD per year (14).

Currently, to our knowledge, there is no comprehensive review in the literature that provides estimates of FGM globally by countries/geographic regions. This review will help track improvements in reaching SDG 5.3. The availability of such information is essential for understanding the burden of FGM globally and for designing effective interventions and preventative measures. An understanding of FGM that considers local social norms contributes to the development of culturally adapted approaches to tackling the issue. Examining variations between countries and regions will allow the development of policies and interventions to be contextualized to the country.

### Objectives:

1. To describe the prevalence of FGM globally, by geographic region and country.
2. To describe the socio-demographic and other determinants of FGM by geographic region and country.

### Methods:

#### Inclusion and Exclusion Criteria

Studies will be included if they meet the following criteria: (i) report on FGM prevalence at the national, subnational, facility or school level (ii), or examine factors associated with FGM within a population. Studies that have a case-series, cross-sectional, case-control or cohort design will be included. To meet the second criterion, the comparison will have to occur between women or girls with FGM to those without FGM. Characteristics to be examined in this review include: socioeconomic status, religion, region, ethnicity, immigrant status ~~and~~, level of education and any other reported risk factor.

Studies will be excluded if they (i) only report on health outcomes of FGM, the attitudes and knowledge of healthcare providers, policy reports, economic effects, or perceptions of FGM; (ii) if they only use qualitative methods, or (iii) are systematic reviews (except for the purpose of referencing), (iv) conference proceedings or letters to the editor. If numerous studies use the same data source, e.g. secondary data analysis of international surveys, ~~both studies will be included in the review to present the total number of eligible studies; however, results will be presented by dataset, and~~ the study with the most complete and comparable data will be included. In particular, objective 1 (prevalence of FGM) will only include the original reports; while objective 2 (determinants of FGM) will include the most relevant and comprehensive study. If the same data source completed multiple studies in a given country, then the most recent study will be included.

#### Definition of FGM

The outcome in this systematic review is FGM, also referred to as female genital cutting or female circumcision. It is a non-medical procedure that entails the total or partial removal of external female genitalia and other injuries to the female genital organs (1). The types of FGM are Type I (clitoridectomy), Type II, Type III (infibulation), and Type IV. Type I, or clitoridectomy, involves the partial or total removal of the prepuce and/or the clitoral gland. Type II involves the partial or total removal of the labia minora and clitoral glans without the excision of the labia majora. Type III, or infibulation, involves narrowing the vaginal canal by modifying the labia majora and minora, and may also include the removal of the clitoral glands. Type IV involves any other non-medical, harmful procedure, such as cauterization, pricking, and scraping (15).

#### Search strategy

Separate searches were conducted using Ovid MEDLINE(R) and EPub Ahead of Print, In-Process & Other Non-Indexed Citations and Daily (1946-present), PsycINFO (1887-present), Web of Science (1900-present), and Embase (1947-present). The last search was conducted on 2nd March, 2020. Together, these databases provide international and interdisciplinary publications. The search strategy is shown in appendix I and was adapted to the format of each database. For the purpose of monitoring progress on SDG 5.3, the search was limited to include publications from 2009 until 2020 in order to provide the most up to date prevalence of FGM. ~~Results were restricted to publications in English, French, and Arabic~~. There were no language restrictions. The MeSH term for FGM was used when possible; otherwise, keywords were used, including “Female Genital Mutilation,” “Female Genital Alteration,” “Female Circumcision,” and “Female Genital Cutting”. The references were imported from each database into EndNote and duplicates were removed. The references were then imported into the systematic review software DistillerSR. Hand searches of the grey literature were conducted through searches of reports from international NGOs, including UNFPA, UNICEF amongst others, and other google searches. In addition, hand searches of the bibliographies of relevant systematic reviews were conducted.

**Study selection procedure**

In the first phase of the review, titles and abstracts will be screened by two separate reviewers working independently of one another to determine studies that qualify for a full-text review based on the abovementioned inclusion and exclusion criteria. Screening will be piloted between the two reviewers to ensure a common understanding of inclusion and exclusion criteria. In the second phase of the review, full texts will be screened by two reviewers, independently and in duplicates, to determine whether a study should be included according to the inclusion and exclusion criteria. Disagreements between the two reviewers will be resolved by discussion or through consulting a third reviewer. The reasons for exclusion at both the screening stage and full-text stage will be recorded. Agreement rate between the two reviewers at the full-text stage will be calculated using Cohen’s κ; a kappa of 0.8 will be judged as very good (16). The methodology was based on the PRISMA guidelines (17).

#### Data extraction

Data will be extracted from included articles using a structured data extraction form, which will be uploaded onto the DistillerSR interface. The data will be extracted by a primary reviewer, and a secondary reviewer will validate the information collected. Disagreements between the two reviewers at the data extraction phase will also be resolved by discussion or through consulting a third reviewer. Items extracted from each article will include: author, publication year, time of data collection, age range of population/ sample, study design, sample size, sampling method, country of origin, host country/region, ethnicity of sample, FGM elicitation method, national/regional crude prevalence and confidence intervals or standard error. Moreover, when available, information about the characteristics of FGM will be abstracted; this will include type, (Type I, Type II, Type III, which is also known as infibulation, and Type IV), who conducted the procedure (traditional circumciser/birth attendant or nurse/midwife/doctor), and the site of where the procedure was done (hospital, home or other). Finally, data on determinants that will be extracted include mother’s education, ethnicity, religion, region, and wealth and whether the study population were a migrant population. In order to determine the impact of these factors, the odds ratio/risk ratio for each variable will be extracted. If a study has not presented an odds ratio or risk ratio, the percentage in each category will be extracted.

#### Risk of bias

Studies will be assessed for risk of bias independently by two reviewers using study specific risk of bias tools.

Studies that present a prevalence estimate, including studies that present both determinants and prevalence of FGM, will be assessed using an adapted tool by Hoy et al., which is specific to prevalence studies (18). This tool **includes nine items that collectively assess the selection bias, representativeness of the sample, validity of the tool and appropriateness of the estimate. Each item will be scored as low or high risk of bias, and each paper will be given an overall score rated as low, moderate or high risk of bias. Studies that are not representative of the study population will be excluded from the pooled prevalence estimate.**

**For studies that only present determinants of FGM, the risk of bias will be assessed using the Evidence Project’s Clarity risk of bias tool for cross-sectional (19), case-control (20), and cohort studies (21). There are eight key domains assessed using this tool. Each study will be assessed on the eight domains and scored as** “definite risk of bias,” “probable risk of bias,” “probable risk of low bias,” or “definite risk of low bias”. **Studies will be judged as having either a “low risk of bias for all key domains”, “unclear risk of bias for one or more key domains” and “high risk of bias for one or more key domains”.** Disagreements between the two reviewers will be resolved by discussion or through consulting a third reviewer. **Studies will only be assessed for risk of bias using one tool. No studies will be excluded as a result of the risk of bias assessment.**

#### Data synthesis

Prevalence estimates from the different studies will be grouped together by country, region sub-population and study type. Studies that used population-based methods or representative sampling methods will be presented separately from studies that used non-representative sampling methods. A pooled national or regional estimate of FGM will only be presented from studies that used representative samples/population-based methods. Studies that examined FGM prevalence using non-representative sampling methods, e.g. single facility or school-based studies, will be excluded from pooled national/region prevalence estimates. Studies that present no standard error or 95% confidence interval will be excluded from the prevalence estimate. Heterogeneity will be assessed using the *I*^2^ statistic and low, moderate and high heterogeneity will be assessed using *I*^2^ results of 25, 50 and 75%, respectively (22). A meta-analysis of prevalence will be estimated using random effect models. A random-effects meta-analysis will be conducted due to the expectation of high heterogeneity between studies. The prevalence estimates will be transformed using the double arcsine method and presented using a pooled prevalence with a 95% CI (23). After piloting the data extraction form it became clear that many studies presented prevalence estimates for both women of reproductive age (15-49 years old) and girls (0-14 years old). All pooled meta-analyses will be presented by age group, in particular, by girls aged 0-14 and women aged 15-49. Primarily, prevalence estimates will be presented in forest plots by geographical regions, data source, age group and study type. If applicable in a subgroup meta-analysis, the prevalence estimates will be presented by migrant status. Meta-bias will also be evaluated using funnel plots for a visual inspection of asymmetry and tested using the Egger’s test to detect for publication bias (24). Funnel plot asymmetry test will be performed only if the meta-analysis includes at least ten studies.

All potential determinants in each study will be presented with a measure of effect. Initially, the broad themes of the determinants will be presented in a narrative synthesis, and these will be summarized by geographical region, sub-population and study type. If there are homogeneous determinants and similar study types, then a meta-analysis may be considered, and results will be presented using forest plots. Statistical analyses and meta-analyses will be conducted using Stata version 15.

**Funder/Sponsor:** This study had no formal source of funding.

| **Review progress** | **Completion date** |
| --- | --- |
| Search completed | March 2020 |
| Titles and abstracts screening | May 2020 |
| Full text screening | June 2020 |
| Data extraction form developed | July 2020 |
| Data extraction | October 2020 |
| Analysis and meta-analysis | February 2021 |

**Appendix I**

**Search strategy**

OVID MEDLINE:

1 female.mp.
2 Girl*.mp.
3 wom?n.mp.
4 or/1-3
5 adj3 genital*.mp.
6 adj3 mutilation.mp.
7 adj3 circumcis*.mp.
8 adj3 cut*.mp.
9 adj3 alter.mp.
10 adj3 alteration.mp.
11 or/5-10 (1448)
12 4 and 11
13 Female Genital Mutilation.mp.
14 Female Circumcision.mp.
15 Female Genital Cutting.mp.
16 Female Genital Alteration.mp. (1629)
17 or/13-16
18 Circumcision, Female.sh. / (1322)
19 12 or 17 or 18 (2024)
20 limit 4 to yr="2009 -Current" (1066)

PsycINFO and WEB OF SCIENCE:

1 Female Genital Mutilation

2 Female Circumcision

3 Female Genital Cutting

4 Female Genital Alteration

5 or/1-4

6 Limit 5 by 2009-2020

EMBASE:

1 'female genital mutilation'/exp

2 'female genital mutilation'

3 or/1-2

4 'female'/exp

5 female

6 or/4-5

7 genital

8 'mutilation'/exp

9 mutilation

10- or/8-9

11 7 and 10

12 11 and 6

13 Limit 12 by 2009-2020
