## Supplementary material for "The global prevalence of female genital mutilation/cutting: A systematic review and meta-analysis of national, regional, facility and school-based studies": S4 Results

**S4 Results: Nationally representative studies**

**S4 Table.** Characteristics of nationally representative studies

| **WHO Region** | **Country, Survey** | **Author** | **Publication date** | **Year of data collection** | **Population description** | **Age category (years)** | **Type** |
| --- | --- | --- | --- | --- | --- | --- | --- |
| **AFR** | **Benin, MICS[1]** | Institut National de la Statistique et de l’Analyse Économique | 2015 | 2014 | Women of reproductive age and girls. | 0-14; 15-49 | Flesh removed, Nicked, Sewn closed. |
|  | **Burkina Faso, DHS[2]** | National Institute of Statistics and Demography (INSD) and ICF International | 2012 | 2010 | Women of reproductive age and girls. | 0-14; 15-49 | Flesh removed, Nicked, Sewn closed. |
|  | **Central African Republic, MICS [3]** | Institut Centrafricain des Statistiques et des Etudes Economiques et Sociales | 2021 | 2018-2019 | Women of reproductive age and girls. | 0-14; 15-49 | Flesh removed, Nicked, Sewn closed. |
|  | **Chad, MICS [4]** | INSEED & UNICEF | 2021 | 2019 | Women of reproductive age and girls. | 0-14; 15-49 | Flesh removed, Nicked, Sewn closed. |
|  | **Cote D'Ivoire, MICS[5]** | Institut National de la Statistiques (INS) | 2017 | 2016 | Women of reproductive age and girls. | 0-14; 15-49 | Flesh removed, Nicked, Sewn closed. |
|  | **Ethiopia, DHS[6]** | Central Statistical Agency & ICF | 2017 | 2016 | Women of reproductive age and girls. | 0-14; 15-49 | Cut with flesh removed, Cut with no flesh removed, Sewn closed, Not sewn closed |
|  | **Eritrea, Population and Health Survey [7]*** | National Statistics Office | 2013 | 2010 | Women of reproductive age and their daughters. | 0-14; 15-49 | NA |
|  | **Gambia, DHS[8]** | Gambia Bureau of Statistics | 2021 | 2019-2020 | Women of reproductive age and girls. | 0-14; 15-49 | Flesh Removed, Nicked, Sewn Closed, Not sewn closed |
|  | **Ghana, MICS[9]** | Ghana Statistical Service | 2018 | 2017- 2018 | Women of reproductive age and girls. | 0-14; 15-49 | Flesh removed, Nicked, Sewn closed |
|  | **Guinea, DHS[10]** | Institut National de la Statistique Ministère du Plan et du Développement Economique & ICF | 2019 | 2018 | Women of reproductive age and girls. | 0-14; 15-49 | Flesh Removed, Nicked, Sewn closed, Not sewn closed |
|  | **Guinea Bissau, MICS**[11] | Ministério da Economia, do Plano e Integração Regional | 2020 | 2018, 2019 | Women of reproductive age and girls. | 0-14; 15-49 | Flesh Removed, Nicked, Sewn closed, Not determined |
|  | **Kenya, DHS[12]** | Kenya National Bureau of Statistics & ICF | 2015 | 2014 | Women of reproductive age and girls. | 0-14; 15-49 | Cut with flesh removed, Cut with no flesh removed, Sewn closed, Not sewn closed |
|  | **Liberia, DHS[13]** | Liberia Institute of Statistics and Geo-Information Services (LISGIS) | 2021 | 2019-2020 | Women of reproductive age | 15-49 | NA |
|  | **Mali, DHS[14]** | Institut National de la Statistique (INSTAT), Cellule de Planification et de Statistique Secteur Santé-Développement Social et Promotion de la Famille (CPS/SS-DS-PF) & ICF. | 2019 | 2018 | Women of reproductive age and girls. | 0-14; 15-49 | Cut with flesh removed, Cut with no flesh removed, Sewn closed, Not sewn closed |
|  | **Mauritania, MICS[15]** | Mauritania National Statistics Office | 2017 | 2015 | Women of reproductive age and girls. | 0-14; 15-49 | Cut and no flesh removed, Cut and flesh removed, Sewn closed, |
|  | **Nigeria, DHS[16]** | National Population Commission (NPC) [Nigeria] & ICF. | 2019 | 2018 | Women of reproductive age and girls. | 0-14; 15-49 | Cut and no flesh removed, Cut and flesh removed, Sewn closed, Not sewn closed |
|  | **Niger, DHS[17]** | Institut National de la Statistique (INS) & ICF | 2013 | 2012 | Women of reproductive age. | 15-49 | Flesh removed, Nicked, Sewn closed |
|  | **Senegal, DHS [18]** | Agence Nationale de la Statistique et de la Démographie (ANSD) | 2020 | 2019 | Women of reproductive age and girls | 0-14; 15-49 | Flesh removed, Nicked, Sewn closed, Not Sewn closed |
|  | **Sierra Leone DHS[19]** | Statistics Sierra Leone & ICF | 2020 | 2019 | Women of reproductive age and girls. | 0-14; 15-49 | Flesh Removed, Nicked, Sewn closed, Not Sewn Closed. |
|  | **Tanzania, DHS[20]** | Ministry of Health, Community Development, Gender, Elderly and Children (MoHCDGEC) [Tanzania  Mainland], Ministry of Health (MoH) [Zanzibar], National Bureau of Statistics (NBS), Office of the Chief Government Statistician (OCGS) & ICF. | 2016 | 2015- 2016 | Women of reproductive age and their daughters. | 0-14; 15-49 | Cut, with flesh removed; Cut with no flesh removed, Sewn closed |
|  | **Togo, MICS[21]** | Institut National de la Statistique et des Etudes Economiques et Démographiques (INSEED) | 2018 | 2017 | Women of reproductive age and girls. | 0-14; 15-49 | Cut with flesh removed, Cut with no flesh removed, Sewn closed |
|  | **Uganda, DHS[22]** | Uganda Bureau of Statistics (UBOS) & ICF | 2018 | 2016 | Women of reproductive age. | 15-49 | NA |
|  | **Multi-country, Tolerance and Tension: Islam and Christianity in Sub-Saharan Africa* [23]** | Pew Research Center | 2010 | 2008, 2009 | Daughters. | NA | NA |
| **EMR** | **Egypt, DHS[24]** | Ministry of Health and Population [Egypt], El-Zanaty and Associates [Egypt] & ICF | 2015 | 2014 | Women of reproductive age and girls. | 0-19; 15-49 | NA |
|  | **Iraq, MICS[25]** | Central Statistical Organization | 2019 | 2018 | Women of reproductive age and girls. | 0-14; 15-49 | Flesh removed, Nicked, Sewn closed |
|  | **Somalia, DHS[26]** | Directorate of National Statistics | 2020 | 2018-2019 | Women of reproductive age and girls. | 0-14; 15-49 | Sunni (Type I), Intermediate (Type II), Pharaonic (Type III or IV) |
|  | **Sudan, MICS[27]** | Central Bureau of Statistics (CBS) & UNICEF Sudan | 2016 | 2014 | Women of reproductive age and girls | 0-14; 15-49 | Flesh removed, Nicked, Sewn closed |
|  | **Yemen, DHS[28]*** | Ministry of Public Health and Population (MOPHP), Central Statistical Organization (CSO) [Yemen], Pan Arab  Program for Family Health (PAPFAM), and ICF International. | 2015 | 2013 | Women of reproductive age and their daughters | 0-14; 15-49 | Cut with flesh removed, Cut with no flesh removed, |
| **SEAR** | **Maldives, DHS[29]** | Ministry of Health (MOH) [Maldives] and ICF. | 2018 | 2016- 2017 | Women of reproductive age and girls. | 0-14; 15-49 | NA |
|  | **Indonesia, RISKESDAS[30]*** | Health Research and Development Agency | 2013 | 2013 | Girls. | 0-11 | NA |

Abbreviations: AFR: African Region; DHS: Demographic and Health Survey; EMR: Eastern Mediterranean Region; MICS: Multiple Indicator Cluster Surveys; SEAR: South-East Asia Region; WHO: World Health Organization.

*Not included in meta-analysis.

**References**

1.	Institut national de la statistique et de l’analyse économique (INSAE). Enquête par grappes à indicateurs multiples 2014, Rapport final. Cotonou, Bénin: UNICEF, 2015.

2.	Institut National de la Statistique et de la DÈmographie IBF, International ICF. Burkina Faso EnquÍte DÈmographique et de SantÈ et ‡ Indicateurs Multiples (EDSBF-MICS IV) 2010. Calverton, Maryland, USA: Institut National de la Statistique et de la DÈmographie - INSD/Burkina Faso and ICF International, 2012.

3.	Institut Centrafricain des Statistiques et des Etudes Economiques et Sociales. Central African Republic Multiple Indicator Cluster Survey 2018-2019 Bangui, République Centrafricaine: UNICEF, 2021.

4.	INSEED and UNICEF. Enquête par grappes à indicateurs multiples Tchad 2019. N’Djamena, Tchad: UNICEF, 2021.

5.	Institute National de la Statistique. Enquête par grappes à indicateurs multiples - Côte d’Ivoire 2016. Cote D’Ivoire: UNICEF, 2017.

6.	Central Statistical Agency & ICF. Ethiopia Demographic and Health Survey 2016. Addis Ababa, Ethiopia: CSA and ICF, 2017.

7.	National Statistics Office Fafo Institute For Applied International Studies. Eritrea Population and Health Survey 2010. Asmara, Eritrea: World Health Organisation, 2013.

8.	Gambia Bureau of Statistics. The Gambia Demographic and Health Survey 2019-20. Banjul, The Gambia: The DHS Program ICF Rockville, Maryland, USA, 2021.

9.	Ghana Statistical Service. Multiple Indicator Cluster Survey (MICS2017/18), Survey Findings Report. Accra, Ghana: UNICEF, 2018.

10.	Institut National de la Statistique Ministère du Plan et du Développement Economique & ICF. République de Guinée Enquête Démographique et de Santé 2018. Conakry, Guinée: The DHS Program, ICF Rockville, Maryland, USA, 2019.

11.	Ministério da Economia e Finanças D-GdP, Instituto Nacional de Estatistica,. Inquérito aos Indicadores Múltiplos (MICS6) 2018-2019, Relatório Final. Bissau, Guiné-Bissau: UNICEF, 2020.

12.	Kenya National Bureau of Statistics & ICF. Kenya Demographic and Health Survey 2014. Rockville, MD, USA: Kenya National Bureau of Statistics and ICF International, 2015.

13.	(LISGIS) LIoSaG-IS. Liberia 2019-2020 DHS. Freetown, Liberia: 2021.

14.	Institut National de la Statistique. Mali Demographic and Health Survey 2018. Bamako, Mali: 2019.

15.	Mauritania National Statistics Office. Enquête par Grappes à Indicateurs Multiples, 2015, Résultats clés. Nouakchott, Mauritanie: UNICEF, 2016.

16.	National Population Commission (NPC) [Nigeria] & ICF. Nigeria Demographic and Health Survey 2018 - Final Report. Abuja, Nigeria: NPC and ICF, 2019.

17.	Institut National de la Statistique INSN, International ICF. Niger Enquéte DÈmographique et de Santè et ‡ Indicateurs Multiples (EDSN-MICS IV) 2012. Calverton, Maryland, USA: INS/Niger and ICF International, 2013.

18.	Agence Nationale de la Statistique et de la Démographie (ANSD). Enquête Démographique et de Santé Continue - EDS-Continue 2019. Dakar, Sénégal: The DHS Program ICF Rockville, Maryland, USA 2020.

19.	Statistics Sierra Leone & ICF. Sierra Leone Demographic and Health Survey 2019. Freetown, Sierra Leone: Ministry of Health and Sanitation and ICF International, 2020.

20.	Ministry of Health Tanzania and Zanzibar National Bureau of Statistics Office of Chief Government Statistician Zanzibar ICF. Tanzania Demographic and Health Survey and Malaria Indicator Survey 2015-2016. Dar es Salaam, Tanzania: MoHCDGEC, MoH, NBS, OCGS, and ICF, 2016.

21.	Institut National de la Statistique et des Etudes Economiques et Démographiques. Togo - Enquête à Indicateurs Multiples 2017, Rapport final. Lomé, Togo: UNICEF, 2018.

22.	Uganda Bureau of Statistics and ICF. Uganda Demographic and Health Survey 2016. Kampala, Uganda: UBOS and ICF, 2018.

23.	Pew Research Center. Tolerance and Tension: Islam and Christianity in Sub-Saharan Africa. Pew Research Center, 2010.

24.	Ministry of Health Population El Zanaty and associates and ICF International. Egypt Demographic and Health Survey 2014. Cairo, Egypt: Ministry of Health and Population and ICF International, 2015.

25.	Central Statistical Organization. Iraq Multiple Indicator Cluster Survey 2018 Survey Findings Report. Iraq: UNICEF, 2019.

26.	Directorate of National Statistics. Somalia Health and Demographic Survey 2020. Federal Government of Somalia, United Nations Population Fund, 2020.

27.	Central Bureau of Statistics & UNICEF Sudan. Multiple Indicator Cluster Survey 2014 of Sudan, Final Report. Khartoum, Sudan: UNICEF, 2016.

28.	Ministry of Public Health and Population. Yemen National Health and Demographic Survey 2013. Rockville, Maryland, USA: MOPHP, CSO, PAPFAM, and ICF International, 2015.

29.	Ministry of Health and ICF. Maldives Demographic and Health Survey 2016-17. Malé, Maldives: MOH and ICF, 2018.

30.	Health Research and Development Agency. Riset Kesehatan Dasar (RISKESDAS) 2013. Indonesia: Government of Indonesia, 2013.
