## Supplementary material for "The global prevalence of female genital mutilation/cutting: A systematic review and meta-analysis of national, regional, facility and school-based studies": S5 Results

**S5 Results: Sub-Regional Studies**

**S5 Table.** Characteristics of Sub-Regional Representative Studies

|  | **Author** | **Year** | **Date of Data collection** | **Sampling Method** | **Sub region** | **Population Description** | **Ethnicity** | **Age** | **Data Collection Site** | **Types** | **Risk of Bias** |
| --- | --- | --- | --- | --- | --- | --- | --- | --- | --- | --- | --- |
| **South Africa** | Scorgie [1] | 2010 | 2007 | Multi-stage cluster | KwaZulu-Natal | Women | NA | 18-60 | Household | NA | Mod. |
| **Burkina Faso** | Greis[2] | 2020 | 2017 | Two-stage stratified (Villages)  Random (Participants) | 10 villages and one sector of Nouna town. | Young women and girls | NA | 12-20 | Village | Nicked, Sewn closed | Low |
|  | Komboigo[3] | 2019 | 2016 | Multi-staged cluster sampling, Cross-Sectional | Ouagadougou | Women living with a partner | NA | 16-65 | Community site | NA | Mod. |
| **Ethiopia** | Mitike [4] | 2009 | 2004 | Systematic random sampling | Somali Regional State in Eastern Ethiopia. Aysha refugee camp (45.1%), Kebribeyah (42.7%), Hartishek (12.2%). | Refugees from Somalia | Somali (100%) | 1-12 | Household | Clitoral cutting, Vaginal sewing | Mod. |
|  | Yirga  [5] | 2012 | 2008 | Systematic random sampling | Kersa district, East Hararge, Oromia region, Ethiopia | Women of reproductive age reporting on daughters. | Oromo (95.7%), Amhara (4.2%), Gurage (0.1%) | 15-49 | Household* | Clitoris only; Clitoris, labia; Clitoris, labia, and surrounding part; Clitoris, labia, surrounding part and stitching; | Mod. |
|  | Gajaa[6] | 2016 | 2014 | Two stage systematic random sampling | Hababo Guduru District | Women of reproductive age and girls | NA | 0-14, 15-49 | District | NA | Low |
|  | Oljira[7] | 2016 | 2013 | Two stage systematic random sampling | Harar | Women and daughters | Oromo (39.2%), Harari (5.0%), Amhara (37.8%), Tigre (4.6%), Gurage (8.9%), Other (4.5%) | 0-12 (girls) | Household | Removal of flesh, Cut without removal of flesh, Stitched with or without removing flesh | Mod. |
|  | Gebrekirstos[8] | 2014 | 2013 | Multi-stage random sampling | Axum town, North Ethiopia | Mothers and children under 5 | NA | <5 | Community | NA | Mod. |
|  | Andualem[9] | 2016 | 2014 | Systematic random sampling | East Gojjam Zone, Western Amhara | Mothers and infants | Amhara (97%), Others, (3%) | 15-49, <5 | Household | 1,2 | Low |
|  | Bogale[10] | 2014 | 2014 | Stratified random sampling | Bale zone | Women of reproductive age and daughters | Oromo (87.2%), Amhara (12.8%) | 15-49 (women) | Community | NA | Low |
|  | Sood[11] | 2022 | 2021 | Multi-stage sampling; random stratified sample of clusters | Afar and Southern Nations, Nationalities and Peoples (SNNP), Addis Ababa City Administration. | Households with adolescent girl and a primary caregiver in the household |  | 10-19 (girls), and older women | Household | NA | Mod. |
|  | Abebe[12] | 2020 | 2020 | Multi-stage sampling; Systematic random sampling | Afar and Amhara | Women with daughters under 15 years old |  | 15-49 (women), 0-14 (girls) | Household | NA | Low |
|  | Melese[13] | 2020 | 2018 | Multi-stage sampling; Systematic random sampling | Degadamot district, Amhara regional state, Northwest Ethiopia | Women with daughters under 5 years old |  | 0-5 Girls | Household | NA | Low |
|  | Gudeta[14] | 2022 | 2019 | Multi-stage cluster sampling | Keffa Zone, Southwest Ethiopia | Mothers with daughters younger than 15 years |  | Mothers 15-49, daughters under 15 years | Community | Nicked, Flesh removed | Low |
| **Kenya** | Mudege[15] | 2012 | 2007 -2008 | Population Based (Every household was visited) | Korogocho and Viwandani , 2 informal settlements in Nairobi, Kenya | Girls and women in informal settlements | Kikuyu (36.5%), Kisii (13.8%), Somali (12.9%), Garre (14.2%), Borana (11.9%), Other (17.3%) | 12-24 | Household | NA | Low |
| **Nigeria** | Ifeanyichukwu[16] | 2015 | 2014 | Cluster Sampling | Okada Community, Edo State | Women of reproductive age | Benin (36.3%), Yoruba (15.7%), Esan (15.4%), Igbo (12.0%), Urhobo (10.5%), others: Ijaw, Efik, Igala and Ogoja (10.2%) | 15-49 | Community | NA | Low |
|  | Alo[17] | 2011 | 2010 | Stratified random sampling | Southwest Nigeria | Mothers with at least one circumcised daughter | NA | 15-49 | Household | NA | Mod. |
|  | Adeyinka[18] | 2009 | 2007 | Multi-stage sampling | Igbo-Ora town, Oyo state, Southwestern Nigeria | Adult women | Yoruba (94.4%), Ibo (2.5%), Hausa (1.8%), Others (0.7%) | 18+ (18 to 80 years) | Household | NA | Low |
|  | Johnson[19] | 2012 | NA | Multi-stage (location) and random (participants) | 3 villages in Itu, Akwa Ibom state, southern Nigeria | Women | Ayadehe 100% | <20 – 40+  18+ | Household | 1, 2, unclear | Low |
|  | Jimoh[20] | 2018 | 2011 | All possible participants  (community) | Tsibiri village, Giwa Local Government area of Kaduna state, Nigeria. | Women and girls who were married, answering about their children | Hausa Fulani | 10-49 (mothers) | Household | NA | Mod. |
| **Tanzania** | Galukande[21] | 2015 | 2013-2014 | Multi-stage sampling; Systematic random sampling | Arusha | Women | Masai (predominant) | 18+ Mean 34 years | Household |  | Low |
| **Sierra Leone** | Bjälkander[22] | 2012 | 2009-2010 | Simple random sampling (streets), systematic random sampling (households),  purposive (sampling participants) | Bombali and Port Loko Districts in Northern Sierra Leone | Young women and girls | Fulah (3.5%), Kono (2.0%), Korankoh (0.6%), Limba (15.5%), Loko (23.2%), Madingo (2.0%), Mende (0.6%), Temne (52.6%) | 10-20 | Household | NA | Mod. |
| **Mali** | Diabate[23] | 2019 | 2009 | Cross sectional Household Survey -Enquête Nationale sur l’Excision au Mali | Kayes, Koulikoro, Sikasso, Segou, Mopti, and Bamako |  | Girls living in localities | 0-8 | Household | NA | High |
| **Somalia** | Gele[24] | 2013 | 2011 | Systematic random sampling | Hargeisa district | Women | NA | 18+  31 (Mean) | Household | 1,2,3 | Mod. |
|  | UNICEF Somalia [25] | 2014 | 2011 | Multi-stage cluster sampling | Somaliland | Women and girls | NA | 0-14, 15-49 | Household | Flesh removed, nicked, sewn closed | Low |
|  | UNICEF Somalia [26] | 2014 | 2011 | Multi-stage cluster sampling | Northeast Zone | Women and girls | NA | 0-14, 15-49 | Household | Flesh removed, nicked, sewn closed | Low |
| **Yemen** | Alosaimi[27] | 2019 | 2008-2009 | Multi-stage sampling | Alazareq (Dhale Governorate), Asabrah and Fara’a Aludain (Ibb Governorate), Mokaa and Maoza (Taiz Governorate), and Azidiah (Hodeidah Governorate). | Women of reproductive age. | NA | 15-49 | Household | NA | Low |
| **Iraq** | Abdulah[28] | 2019 | 2017 | Two stage random sampling | Iraqi Kurdistan region (Duhok, Erbil, and Sulaiymaniya) | Women of all ages | NA | All age groups | Household | NA | Mod. |
| **Egypt** | Ali [29] | 2018 | 2017 | Random cluster sampling (areas), random sampling (households) | Beni-Suef | Young women and girls. | NA | 12-25 | Household | NA | Mod. |
|  | Mohammed[30] | 2018 | 2016 | Multi-stage systematic random sampling | Rural area in Minia | Rural area inhabitants | NA | 18+ | Household | NA | Low |
|  | Salama[31] | 2021 | NA | Multi-stage cluster sampling | Six governorates representing Egypt | Children |  | 9 months – 16 years | Community | NA | Mod. |
|  | Zayed[32] | 2012 | NA | Random | Cairo & Giza | Women and girls | NA | 5-30 | Community | NA | Mod. |
| **Saudi Arabia** | Milaat[33] | 2018 | 2017 | Random cluster sampling (location), multi-stage random sampling (household) | Hali semi-urban region. | Girls | NA | ≤18 years | Household | NA | Mod. |

*Patient report and examination, all others: Patient Report † women reported that at least 1 daughter had FGM/C in the household.

All studies used cross-sectional methods.

**S6 Table.** Prevalence of FGM/C in Women and Girls in Sub-Regional Representative Studies

|  | | | | | | | **Women** | | | **Girls** | | |
| --- | --- | --- | --- | --- | --- | --- | --- | --- | --- | --- | --- | --- |
|  | **Author** | **Year** | **Sub region** | **Sampling method** | **Population description** | **Age** | **Prevalence %** | **Total FGM/C** | **Sample Size** | **Prevalence %** | **Total FGM/C** | **Sample Size** |
| **EMR** | | | | | | | | | | | | |
| **Somalia** | Gele[24] | 2013 | Hargeisa district, Somalia | Systematic random sampling | Adult women | Mean: 31, Range: <25-41+ (includes males) | 97% | 104 | 107 |  |  |  |
|  | UNICEF Somalia [25] | 2014 | Somaliland | Multistage cluster sampling | Women and girls | 0-14, 15-49 | 99.1% | 5812 | 5,865 | 27.7% | 1587 | 5,729 |
|  | UNICEF Somalia [26] | 2014 | Northeast Zone | Multistage cluster sampling | Women and girls | 0-14, 15-49 | 98% | 5382 | 5492 | 30.6% | 1779 | 5,813 |
| **Yemen** | Alosaimi [27] | 2019 | Alazareq (Dhale Governorate), Asabrah and Fara’a Aludain (Ibb Governorate), Mokaa and Maoza (Taiz Governorate), and Azidiah (Hodeidah Governorate). | Multi-stage sampling | Women of reproductive age | 15-49 (mothers) | 47.8% | 3384 | 7076 | 34%* | 2405 | 7076 |
| **Egypt** | Ali[29] | 2018 | Beni-Suef | Random cluster sampling (areas), random sampling (households) | Young women and girls | 12-25 | 55% | 1846 | 3353 |  |  |  |
|  | Mohammed [30] | 2018 | Rural area in Minia | Multi-stage systematic random sampling | Rural area inhabitants | 18+ | 76.6% | 320 | 418 |  |  |  |
|  | Zayed[32] | 2012 | Cairo & Giza | Random | Women and girls | 5-30 | 63.9% | 156 | 244 |  |  |  |
|  | Salama[31] | 2021 | Six governorates representing Egypt | Multi-stage cluster sampling | Children | 9 months – 16 years |  |  |  | 29.7% | 508 | 1,710 |
| **Iraq** | Abdulah[28] | 2019 | Iraqi Kurdistan region( Duhok, Erbil, and Sulaiymaniy) | Two stage random sampling | Women | All age groups | 46.8% | 2361 | 5048 |  |  |  |
| **Saudi Arabia** | Milaat[33] | 2018 | Hali semi-urban region | Random cluster sampling (location), multi-stage random sampling (household) | Girls | ≤18 years |  |  |  | 80.3% | 175 | 218 |
| **AFR** | | | | | | | | | | | | |
| **Tanzania** | Galukande[21] | 2015 | Arusha | Multi-stage sampling; Systematic random sampling | Women | 18+ Mean 34 years | 69.2% | 467 | 675 |  |  |  |
| **South Africa** | Scorgie[1] | 2010 | KwaZulu-Natal | Multi-stage cluster | Women | 18-60 | 3% | 26 | 867 |  |  |  |
| **Nigeria** | Ifeanyichukwu[16] | 2015 | Okada Community, Edo State | Cluster Sampling | Women of reproductive age | 15-49 | 28.7% | 90 | 325 |  |  |  |
|  | Jimoh [20] | 2018 | Tsibiri village, Giwa Local Government area of Kaduna state, Nigeria. | All possible participants | Girls and women answering for their children | 10-49 |  |  |  | 27.7% | 61 | 220 |
|  | Alo[17] | 2011 | Southwest Nigeria | Stratified random sampling | Mothers with at least one daughter circumcised | 15-49 | 75% | 315 | 420 | 71%* | 298 | 420 |
|  | Adeyinka [18] | 2009 | Igbo-Ora town, Oyo state, Southwestern Nigeria | Multi-stage sampling | Adult women | 18+ | More than 78.7% | 118 | 155 |  |  |  |
|  | Johnson[19] | 2012 | 3 villages that make up the Ayadehe clan in Itu local govt. area in Akwa Ibom state, southern Nigeria | Multi-stage (location) and random (participants) | Women | <20 – 40+ | 92.7% | 202 | 218 |  |  |  |
| **Ethiopia** | Oljira[7] | 2016 | Harar | Two stage systematic random sampling | Women and girls | 0-12 (daughters)  15+ (mothers) | 79.5% | 669 | 842 | 19% | 160 | 842 |
|  | Gajaa[6] | 2016 | Hababo Guduru District | Two stage systematic random sampling | Women of reproductive age and daughters | Women aged 15-49, Girls aged 0-14 | 98.2% | 599 | 610 | 48%* | 293 | 610 |
|  | Gebrekirstos[8] | 2014 | Axum town, North Ethiopia | Multi-stage random sampling | Children under 5 and their mothers | <5  15+ (mothers) | 0.7% | 5 | 746 | 0% | 0 | 752 |
|  | Yirga[5] | 2012 | Kersa district, East Hararge, Oromia region | Systematic random sampling | Women of reproductive age | 15-49 | 92.3% | 792 | 858 | 88.1%* | 288 | 327 |
|  | Andualem [9] | 2016 | East Gojjam Zone, Western Amhara | Systematic random sampling | Mothers and infants | Mothers 15-49, daughters 0-2 | 96% | 689 | 718 | 49% | 403 | 805 |
|  | Bogale[10] | 2014 | Bale zone | Stratified random sampling | Women of reproductive age and daughters | 15-49 (mothers) | 78.5% | 486 | 634 | NA | 150† | NA |
|  | Mitike[4] | 2009 | Aysha, Kebribeyah and Hartishek refugee camps, Somali Regional State in Eastern Ethiopia | Systematic random sampling | Refugees from Somalia | 12+ | 42.4% | 112 | 288 |  |  |  |
|  | Sood [11] | 2022 | Afar and Southern Nations, Nationalities and Peoples (SNNP), Addis Ababa City Administration. | Multi-stage sampling; random stratified sample of clusters | Households with adolescent girl and a primary caregiver in the household | 10-19 (girls), and older women | 65% women and girls combined |  |  |  |  |  |
|  | Abebe[12] | 2020 | Afar and Amhara | Multi-stage sampling; Systematic random sampling | Women with daughters under 15 years old | 15-49 (women), 0-14 (girls) | 98% | 398 | 405 | 75.9%‡ | 796 | 1048 |
|  | Melese[13] | 2020 | Degadamot district, Amhara regional state, Northwest Ethiopia | Multi-stage sampling; Systematic random sampling | Women with daughters under 5 years old | 0-5 Girls |  |  |  | 70.80% | 230 | 325 |
|  | Gudeta[14] | 2022 | Keffa Zone, Southwest Ethiopia | Multi-stage cluster sampling | Mothers with daughters younger than 15 years | Mothers 15-49, daughters under 15 years | 21.2% | 159 | 750 | 1.6% | 12 | 750 |
| **Burkina Faso** | Greis[2] | 2020 | 10 villages and one sector of Nouna town | Two-stage stratified (Villages)  Random (Participants) | Young women and girls | 12-20 | 43.2% | 301 | 696 |  |  |  |
|  | Komboigo[3] | 2019 | Ouagadougou | Multi-Stage cluster | Women living with a partner | 16-65 | 53.4 | 307 | 575 |  |  |  |
| **Kenya** | Mudege [15] | 2012 | Korogocho and Viwandani , 2 informal settlements in Nairobi, Kenya | Population based | Young women and girls in informal settlements | 12-24 | 61.3% | 323 | 527 |  |  |  |
| **Sierra Leone** | Bjälkander [22] | 2012 | Bombali and Port Loko Districts in Northern Sierra Leone | Simple random sampling (streets), systematic random sampling (households),  purposive (sampling participants) | Young women and girls | 10-20 | 61.1% | 189 | 310 |  |  |  |
| **Mali** | Diabate[23] | 2019 | Kayes, Koulikoro, Sikasso, Segou, Mopti, and Bamako | Unclear (Original survey is unavailable) | Girls living in localities | 0-8 |  |  |  | 71.4% | 1413 | 1979 |

* Women reported that at least 1 daughter had FGM/C in the household.. † Youngest daughter had FGM/C. ‡ Due to inconsistent data reported in the study, this number was calculated by the authors of this review.

Abbreviations: AFR: African Region EMR: Eastern Mediterranean Region, FGM/C: Female Genital Mutilation/Cutting

**S7 Table.** Types of FGM/C in Sub-Regional Representative Studies

| **Region** | **Author** | **Year** | **Country** | **Prevalence Women (%)** | **Total FGM/C Women** | **Prevalence Girls (%)** | **Total FGM/C Girls** | **Type 1 (%)** | **Type 2 (%)** | **Type 3 (%)** | **Type 4 (%)** | **Other Type** | **Don't Know/Missing Type** |
| --- | --- | --- | --- | --- | --- | --- | --- | --- | --- | --- | --- | --- | --- |
| AFR | Andualem [9] | 2016 | Ethiopia | 96% | 689 | 49% | 403‡ | 48.9‡ | 51.1%‡ |  |  |  |  |
|  | Bogale[10] | 2014 | Ethiopia | 78.5% | 486 | NA | 150† | 2.5%*, 4.7%† | 78.6%*, 87.3%† | 7.8%*, 8%** |  | 11.1%* |  |
|  | Gudeta[14] | 2022 | Ethiopia | 21.2% | 159 | 1.6% | 12 |  |  |  | 61%*, 16.7% ‡ | Type I and II reported together: 83%‡, 83.3% ‡ |  |
|  | Johnson[19] | 2012 | Nigeria | 92.7% | 202 |  |  | 7.9%* | 71.2%* |  |  |  | 20.9%* |
|  | Greis[2] | 2020 | Burkina Faso | 43.2% | 301 |  |  | 20.8%* | 69.6%* | 2%* |  |  |  |
|  | Gele[24] | 2013 | Somalia | 97% | 104 |  |  |  |  | 81.3% |  | Type I and II reported together: 15.9%* |  |
| EMR | UNICEF Somalia [25] | 2014 | Somalia  (Somaliland DHS) | 99.1% | 5812 | 27.7% | 1587 | 2.7%*,  3.3%‡ | 8.6%*,  9.6%‡ | 84.9%*,  11.6%‡ |  |  | 3%*,3.2%‡ |
|  | UNICEF Somalia [26] | 2014 | Somalia  (Northeast Zone DHS) | 98% | 5382 | 30.6% | 1779 | 1.4%*, 0.9%‡ | 5.7%*, 4.7%‡ | 86.7%*, 22.5%‡ |  |  | 4.2%*, 2.5%‡ |

* % of Women

† % of youngest daughter

‡ % of girls

Somaliland and Northeast Zone MICS calculate the preavelnce of type out of the total number of participants, and report type 2 as “flesh removed” and type 3 as “sewn closed”.

Abbreviations: FGM/C: Female Genital Mutilation/Cutting

**S8 Table.** Characteristics of FGM/C Procedure in Sub-Regional Representative Studies

| **Country** | **Author** | **Year** | **Age at FGM/C** | **Performer of FGM/C** | **Location of Procedure** |
| --- | --- | --- | --- | --- | --- |
| **AFR** | | | | | |
| **Burkina Faso** | Greis[2] | 2020 | 0–4 years (54.0%), 5–9 years (39.5%), 10–14 years (6.2%), 15+ years (0.3%) | Traditional (97.4%), Medical (1.1%), Other (1.6%) |  |
| **Ethiopia** | Oljira[7] | 2016 |  | Relatives, neighbors, and health personnel (5.6%), Traditional (94.4%) |  |
|  | Gajaa[6] | 2016 | Daughters: 1-4 (2%), 5-8 (20.7%), 9-12 (23.1%), 12-15 (1.6%) | Traditional (92.8%) |  |
|  | Mitike[4] | 2009 | Mean age: the first daughter (older) 7.5 (± 1.7), last (younger) daughter 6.5 (± 1.4) years. Optimum age: 5–8 years old | Traditional (99.1%), Medical (0.9%) |  |
|  | Yirga[5] | 2012 |  | Traditional (94.1%) (76.1% by local healers and 18% by elderly people) |  |
|  | Andualem[9] | 2016 |  | Traditional (100%) | Home (96%), Circumciser's home (4%) |
|  | Abebe[12] | 2020 |  | Traditional cutters (72.8%), Traditional birth attendant (8.9%), other (0.9%) |  |
|  | Gudeta[31] | 2022 | Mother's age at circumcision: I don’t know (45.5%); 5–9 (30.8); 10–14 (18.2%); ≥15 (5.7). Daughters: <10 years 83.3%; ≥10 years: 16.7% | Mothers: Traditional (98.7%), health professional (1.3%) Daughters: Traditional (100%) |  |
|  | Bogale[10] | 2014 | Mean 7.89 (SD ± 4.56). |  |  |
| **Kenya** | Mudege[15] | 2012 | Mean: 10, Median: 13 |  |  |
| **Nigeria** | Ifeanyichukwu[16] | 2015 | Mean 4.12 ± 1.17, 0-5 years (83.4%) | Traditional (67.7%), Medical (43.3%) | Home (40%)  Traditional home (33.3%)  Health facility (26.7%) |
|  | Johnson[19] | 2012 | < 6 (5.4%), 6-12 (69.8%), 13-18 (23.3%), 18+ (1.5%) | Traditional (53.5%), Medical (0.5%), Native Doctor (unclear if medically trained) (46%) | Home (99.5%) |
| **Sierra Leone** | Bjälkander[22] | 2012 | 0 – 1 (1.6%), 2 – 4 (7.4%), 5 – 9 (21.3%), 10 – 14 (21.9%), 15+ (2.9%), Don’t Know (5.8%) | Traditional (85.7%), Medical (13.2%), Nurse & Sowei (traditional) together (0.5%), don't know (0.5%) |  |
| **Tanzania** | Galukande[21] | 2015 | ≤ 11 years 107 (15.8%) 12–15years 169 (25%) ≥ 16 years 143 (21.2%) |  |  |
| **Mali** | Diabate[23] | 2019 | <1 year (22.9%), 1 (~5%), 2 (~5%), 3 (~ 5%). 6+ (0%). |  |  |
| **EMR** | | | | | |
| **Egypt** | Mohammed[30] | 2018 | Range 4–17, (mean ± SD) 11.5 ± 2.3 | Traditional (91.3%), Medical (8.7%) |  |
|  | Salama[31] | 2021 |  | Medical professionals (76%), Midwife or Daya (20.1% ), don’t’ know (3.9%) | Private clinic (40.6%)  Home (59.4%) |
|  | Zayed[32] | 2012 | Mean 10.846 (SD ± 1.98) years and ranged between 8-15 | Traditional (33.3%), Medical (64.1%), Other (2.6%) | Home (56.5%), Clinic (43.5%)  Hospital (5%) |
| **Saudi Arabia** | Milaat[33] | 2018 | 7 or less (59.4%), 8-10 (0.6%), 11-14 (2.9%), 15-17 (1.7%), 18 (35.4%) | Traditional (2.8%), Medical (97.1%) |  |

Abbreviations: AFR: African Region EMR: Eastern Mediterranean Region, FGM/C: Female Genital Mutilation/Cutting

25.	UNICEF Somalia SMoPaND. Somaliland Multiple Indicator Cluster Survey 2011, Final Report. . Nairobi, Kenya: UNICEF, Somalia and Somaliland Ministry of Planning and National Development, Somaliland., 2014.

26.	UNICEF Somalia MoPaIC. Northeast Zone Multiple Indicator Cluster Survey 2011, Final Report. Nairobi, Kenya: UNICEF, Somalia and Ministry of Planning and International Cooperation., 2014.
