## Supplementary material for "The global prevalence of female genital mutilation/cutting: A systematic review and meta-analysis of national, regional, facility and school-based studies": S6 Results

**S6 Results:** School, Community or Facility based studies excluding studies on migrant populations

**S9 Table.** Characteristics of school, community or facility-based studies excluding studies on migrant populations

|  | **Author** | **Year** | **Data**  **Collection** | **Sampling method and Study Design** | **Sub-Region** | **Population Description** | **Ethnicity** | **Age** | **Exam or Patient report** | **Data Collection Site** | **Types** | **Risk of Bias** |
| --- | --- | --- | --- | --- | --- | --- | --- | --- | --- | --- | --- | --- |

| **AFR** | | | | | | | | | | | | |
| --- | --- | --- | --- | --- | --- | --- | --- | --- | --- | --- | --- | --- |
| **Burkina Faso** | Ndiaye[1] | 2010 | 2007 | Purposive, Cross-Sectional | Fada Ngourma | Women who gave birth | NA | 14-44 | Exam | Hospital /Clinic  (Multiple) | 1,2,3 | Low |
|  | Ouédraogo [2] | 2017 | NA | Convenience, Cross Sectional | Boromo and Bittou | Hawkers (street vendors) | NA | 13-24 | Patient Report | Traffic Station | NA | Mod. |
| **Gambia** | Kaplan[3] | 2013 | 2010-2011 | Purposive, Cross Sectional | Western Health Region | Women examined for antenatal care or delivery in hospitals and health centers | Mandinka (n=175), Wolof (n=120), Fula (n=116), Sarahole (n=30), Djola (n=76), Serer (n=13), Other (n=19) | 12-45 | Exam | Hospital/clinic | 1,2,3. | Mod. |
| **Tanzania** | Suleiman[4] | 2021 | 2004-2014 | All women in the hospital facility, Retrospective cross-sectional | Kilimanjaro region | Women who delivered singletons |  | 15-49 | Exam | Hospital/clinic | 1,2,3,4. | Low. |
| **Ethiopia** | Gudu[5] | 2017 | 2015 | Purposive, Cross-Sectional | Jijiga | Nulliparous women who gave birth | Somali (98.6%) and Oroma (1.4%) | Mean 21.8 | Patient report | Hospital s/Clinics | 2 and 3 | Mod. |
|  | Gebremariam[6] | 2016 | 2014 | Muti-stage stratified random sampling, Cross-Sectional | Jigjiga district, Somali regional state, eastern Ethiopia | High school and college students | Somali (65%), Amhara (18.7%), Oromo (29%), Other (13.4%) | 15-24 | Patient report | School | 1,3, 1 and 3 together | Low |
|  | Tamire[7] | 2013 | 2011 | Multi-staged cluster sampling, Cross-Sectional | Hadiya zone, Southern Ethiopia | High school students | Hadiya (89.0%), Gurage (4%), Kambata (3.6%), Amhara (2.3%), Silte (0.9%), other (0.3%) | 13-25 | Patient report | School | NA | Low |
|  | Abathun[8] | 2018 | 2015 | Purposive (location), stratified random sampling (participants), Cross-Sectional | Harari and Somali Regions of Ethiopia | High school students | Somali, Harari, Others | 16-22 | Patient Report | School | 1 and 3 | Low |
|  | Shay[9] | 2010 | 2008 | Random sampling, Cross-Sectional | Addis Ababa | School girls | Amhara (31%), Ooromo (23.6%), Tigray (6.4%), Guraghe (31.2%), Others (7.9%) | <5-20+ | Patient Report | School  (Multiple) | NA | Mod. |
| **Ghana** | Nonterah [10] | 2019 | 2003 to 2013 | Retrospective audit of records, Cohort | Kassena-Nankana district- North Eastern Ghana | Attending Clinic | NA | <20-35+ | Both | Hospital/ Clinic | NA | Mod. |
|  | Sakeah[11] | 2018 | NA | Random and convenience, Cross-Sectional | Bawku municipality and Pusiga District *** | Women of reproductive age | Kusassi (1.33%) Maprussi (21.33%) Busanga (60.96%) Moshie (8.31%) Hausa (1.33%) Other (6.74%) | 15-49 | Patient Report | Household | NA | Low |
| **Mali** | Dicko-Traore[12] | 2014 | 2011 | Purposive, Cross Sectional | Bamako | Girls attending clinic | NA | 0-15 | Exam | Hospital/ Clinic | NA | Mod. |
| **Nigeria**  **Nigeria** | Ezeoke[13] | 2021 |  | Multi-stage sampling of schools and purposive selection of students, cross sectional | North Central Nigeria | Secondary school students |  | 13-19 | Patient Report | School | NA | Mod. |
|  | Chinawa [14] | 2020 |  | Multi-stage sampling of schools; stratified selection of schools; random selection of students, cross sectional | Enugu metropolis | Secondary school students | Igbo (n=412, 99.5%), Hausa (n=1, 0.2%), Yoruba (n=1, 0.2%) | 13-21 years | Patient Report | School | NA | Low. |
|  | Ezenyeaku[15] | 2011 | 2009-2010 | Purposive, Cross Sectional | Enugu and Awka (Southeast geopolitical zone of the Nigeria) | Attending clinic | NA | 14-50 | Patient report | Hospital/ Clinic | NA | Mod. |
|  | Lawani[16] | 2014 | 2012 | Purposive, Prospective | Abakaliki, Ebonyi, southeast Nigeria | Women seeking maternity services | NA | 15-49 | Exam | Hospital/ Clinic  (Multiple) | 1,2,3 | Mod. |
|  | Makinde[17] | 2012 | 2007 | Purposive, Descriptive prospective | South-West of Nigeria | Girls visiting emergency and gynecological wards | Hausa (4.1%), Ibo (4.1%), Yoruba (85.1%), Any Other (6.7%) | 0-14 | Exam | Hospital/ Clinic | 1,2,3,4 | Mod. |
|  | Dattijo[18] | 2010 | 2007 | Systematic random sampling, Cross-Sectional | Jos, north-central Nigeria | Attending antenatal care | Hausa (19.6%), Igbo (15.4%), Yoruba (13.8%), Berom (8.8%), Mughavul (5.8%), Ngas (5.4%) Edo (5%), Tarok (3.8%), Idoma (3.5%), Tiv (3.5%), other (15.4%). | <20-35+ | Patient Report | Hospital/ Clinic | Clitoridectomy, Excision, Infibulations | Mod. |
|  | Ogah[19] | 2019 | 2016 | Consecutive, Cross-Sectional | Ilorin, Kwara state | Attending health clinic | NA | 15-60 | Both | Hospital/ Clinic | NA | Mod. |
|  | Ashimi[20] | 2014 | 2014 | Systematic random sampling, Cross-Sectional | Jigawa state, Northwest Nigeria | Attending antenatal clinic | Hausa (70.6%), Fulani (26.0%), Yoruba (1.9%), Igbo (1.5%) | 15-40 | Patient report | Hospital/ Clinic  (Multiple) | 1,2, Hymen removal, Gishiri, Angurya | Low |
|  | Ashimi[21] | 2015 | 2014 | Systematic random sampling, Cross-Sectional | Three clinics, Birnin Kudu, Jigawa state | Infants presented to 3 clinics | Hausa (73%), Fulani (27%) | 1-21+ days | Both | Hospital/ Clinic  (multiple) | 1,2,3,4 | Low |
|  | Adeniran [22] | 2015 | 2014 | Multistage and purposive, Cross-Sectional | 18 schools in Ilorin, North Central Nigeria, all those eligible were invited | Secondary school teachers | NA | 20-65 | Patient report | School | NA | Mod. |
|  | Ibrahim[23] | 2013 | 2012 | All possible participants were included, Cross-Sectional | Bayelsa state, Niger-Delta of Nigeria | Nurses and doctors | Not FGM/C only: Ijaw (48.3%), Igbo (24.6%), Hausa (1.7%), Others (18.6%) no response (6.8%) | < 25 - 55 + | Patient report | Hospital/ Clinic  (Multiple) | NA | Mod. |
|  | Dike[24] | 2012 | NA | Purposive, Cross-Sectional | Afikpo, Ebonyi State, Southeastern Nigeria | Female students of Nursing and Midwifery | NA | 15-45 | Patient Report | School | NA | Low |
|  | Garba[25] | 2012 | 2011 | Purposive, Cross Sectional | Kano, Northern Nigeria | Infants | Hausa/Fulani ethnic group (58%), other ethnic group (42.3 %) | < 5 – 16 + days | Mother's report | Hospital/ Clinic | 1,2,3,4 | Low |
|  | Iliyasu[26] | 2012 | NA | Multistage sampling, Cross Sectional | Bayero University, Kano, Kano State, Northern Nigeria | University students | Hausa (44.3%), Fulani (17.5%), Yoruba (18.7%), Igbo (4.7%), Others (14.8%) | 17-40 | Patient Report | School | NA | Low |
|  | Jimoh[27] | 2018 | 2011 | Unclear, Cross-Sectional | Tsibiri village, a rural community in Giwa Local Government area of Kaduna state, north western region of Nigeria. | Women and girls who were married | NA | 15-49,10-14 | Patient Report | Community | NA | Low |
|  | Agbede[28] | 2019 | 2018 | Convenience, Cross Sectional | 4 wards in Ede South LGA, Osun State | Parents of daughters with FGM/C | Yoruba (94.7%), Igbo (3.7%), Hausa (1.6%) | 15- 40+ | Patient Report | Community | NA | Mod. |
| **Sierra Leone** | Bjälkander[29] | 2012 | 2006 | Random sampling, Cross-Sectional | Bo Town, Bo District, Southern region and Makeni Town, Bombali District, Northern region | Attending clinic | Temne (30.5%), Limba (26%), Mende (20.8%), Fulah (4.5%), Loko (7.2%), Susu (2.7%), Kono (3.2%), Korankoh (3.2%), Madingo (1.9%) | 11-45 | Patient Report | Hospital/ Clinic  (Multiple) | NA | Low |
| **Uganda** | Ivanova[30] | 2019 | 2018 | Convenience, Cross-Sectional | Nakivale refugee settlement | Refugees from DR Congo, Burundi, Rwanda, Ethiopia, Somalia, Uganda, South Sudan, Tanzania, Eritrea, Kenya | NA | 13-19 | Patient Report | Refugee Camp | NA | Mod. |
| **EMR** | | | | | | | | | | | | |

| **Djibouti** | Minsart[31] | 2015 | 2012-2014 | All possible participants, Cohort | Djibouti City | Women who gave birth | NA | <25-35+ | Patient report and exam | Hospital/ Clinic | 1,2,3 | Mod. |
| --- | --- | --- | --- | --- | --- | --- | --- | --- | --- | --- | --- | --- |
| **Somalia** | Adigüzel[32] | 2018 | 2017 | Purposive, Cross-Sectional | Mogadishu, Beladwayne, Kismaayo, Jawhar, Ceelbuur, Baardhere, Benadir, lower Shabelle, middle Shabelle, Hiiraan, Galgaduud, Mudug, Bakool, Gedo, lower Jubba | Presented to the obstetrics and gynecology outpatient clinics, married with at least one daughter | NA | Mean 28.76SD ±8.77 | Both | Hospital/ Clinic  (Multiple) | 1,2,3,4 | Low |
| **Sudan** | Birge[33] | 2017 | 2014-2015 | Convenience, Cross-Sectional | Darfur | Patients/working at hospital | NA | 17 - 65 | Both | Hospital/ Clinic | 1,2,3 | Mod. |
|  | Ali[34] | 2012 | 2012 | Random, Cross-Sectional | Eight schools in Kassala, Eastern Sudan | Students | Hadandawa (15.0%), Biniamir (40.4%), other tribes ( 44.5%) | 9-16 | Patient Report | School  (Multiple) | NA | Mod. |
|  | Sharfi[35] | 2013 | 2007-2012 | Purposive, Cross-Sectional | Khartoum | University students and out-clinic patients | NA | 20-62 | Both | Hospital/ Clinic and School | 3 | Mod. |
|  | Akbas[36] | 2019 | 2016 | All possible participants, Cross-Sectional | Nyala | University students | NA | Mean 19.5 SD ±1.95 | Patient Report | School | NA | Low |
|  | Mahgoub[37] | 2019 | 2018-2019 | Multi-stage sampling of schools, Quasi-experimental | Karary Locality, Khartoum State, Sudan | Students | Jaalia tribe (31.2%) | 14-17 | Patient report | School | NA | Mod. |
|  | Birge[38] | 2021 |  | Cross sectional, purposive | Nyala, Darfur | Attending clinic |  | 18+ | NA | Clinic | 1,2,3 | Mod. |
| **Iraq** | Yasin[39] | 2013 | 2007-2009 | Convenience, Cross-Sectional | Erbil, Kurdistan region | Attending clinic | NA | 15-49 | Both | Hospital/Clinic  (Multiple) | 1,2 | Mod. |
|  | Saleem[40] | 2013 | 2011 | Purposive, Cross-Sectional | Kurdistan region | Attending clinic | NA | 0.5-20 | Both | Hospital/Clinics  (Multiple) | 1,2,3,4 | Mod |
| **Egypt** | Hassanin[41] | 2012 | 2011 | Purposive, Cross-Sectional | Upper Egypt | Mothers lived in urban areas, completed secondary school, heard about the ban, with children in school | NA | 8-14 | Patient Report | Hospital/Clinic  (Multiple) | NA | High |
|  | Mitwaly[42] | 2017 | 2014-2015 | Purposive, Cross-Sectional | Luxor city ,Upper Egypt | Attending clinic | NA | 15-49 | Exam | Hospital/Clinic | 1,2 | Mod. |
|  | Arafa[43] | 2018 | 2016-2017 | Multistage random sampling, Cross-Sectional | Beni-Suef | University students | NA | Mean/SD 20.89 ± 1.68 | Patient Report | School | NA | Mod. |
|  | Mostafa[44] | 2017 | 2016 | All possible participants, Cross-Sectional | Beni-Suef | Overweight and obese premenopausal women | NA | 20-49 | Patient Report | Hospital/Clinic | NA | Mod. |
|  | Abdel-Aleem[45] | 2016 | 2011-2014 | Purposive, Cross-Sectional | Assiut and Sohag | Recently married | NA | 17-31 | Both | Hospital/Clinic | 1,2,3 | Low |
|  | Ahmed[46] | 2017 | 2015-2016 | Purposive, Cross-Sectional | Suez Canal | Students attending clinic | NA | 14-19 | Patient Report | Hospital/Clinic  (Multiple) | NA | Mod. |
|  | Rasheed[47] | 2011 | 2008-2010 | All possible participants, Cross-Sectional | Sohag and Qena | Attending clinic | NA | 5-25 | Patient Report | Hospital/Clinic | NA | High |
|  | Abolfotouh[48] | 2015 | 2012-2013 | Convenience, Cross-Sectional | Upper (Southern) and lower (Northern) Egypt | Medical students from 19 Egyptian universities, each in a different governorate | NA | 18+ | Patient Report | Online | NA | Low |
|  | Elbendary[49] | 2021 | 2018 | Random selection, Cross-sectional | Fayoum | Attending Clinic | NA | 18-45 | Exam | Clinic | 1,2,3,4 | Low |
|  | Galal[50] | 2022 | NA | Online, Cross-Sectional | Upper and lower Egypt | Medical Students | NA | Mean 21 years | Patient report | Online | NA | Mod. |
| **Iran** | Dehghankhalili[51] | 2015 | 2010-2013 | Purposive, Cross-Sectional | Minab, Dehbaz, Bandar-e-Lenge, Qeshm, Bandar-e-Khamir, and Bastak in Hormozgan, Southern Iran | Attending Clinic | NA | 14-38 | Exam | Clinic | 1,2,3,4, clitoris nicking | Mod. |

**SEAR**

| **Malaysia** | Rashid [52] | 2019 | NA | Snowball, Cross-Sectional | Kedah and Penang, Northern region of Peninsular Malaysia | Villagers | NA | 18+ | Patient report | Village | 4 | Mod. |
| --- | --- | --- | --- | --- | --- | --- | --- | --- | --- | --- | --- | --- |
|  | Rashid[53] | 2009 | 2008-2009 | Convenience, cross sectional | Five villages in north Malaysia. | All women from villages | NA | 1 month-91 years | Patient report | Villages | NA | Low |
|  | Khalid [54] | 2017 | 2012 | Sequential convenience, Cross-Sectional | Selangor | Attending Clinic | Muslims alone: Malay (88.3%) Indonesian (5%), Others (6.7%) / Non- Muslims alone: Chinese (57.1%), Indians (32.7%), Others (10.2%) | 18+ | Patient report | Hospital/Clinic  (Multiple) | NA | Mod. |

*****Types of FGM/C mentioned were: Clitoral tip excision, Complete clitoridectomy, Clitoridectomy/labia minora Excision, Clitoridectomy/labia minora/Inner majora excision.

Abbreviations: AFR: African Region EMR: Eastern Mediterranean Region SEAR: South East Asian Region

**S10 Table.** Proportion of FGM/C in Women and Girls in School, Community or Facility based excluding studies on migrant populations.

|  | **Author** | | **Year** | **Sampling method** | **Sub- Region** | | **Population description** | **Age** | **FGM/C (%)** | **Total FGM** | **Sample Size** |
| --- | --- | --- | --- | --- | --- | --- | --- | --- | --- | --- | --- |
| **Nigeria** | Ashimi[20] | | 2014 | Systematic random sampling | Jigawa state, Northwest Nigeria | | Attending antenatal clinic | 15-40 | 30.9% | 100 | 323 |
|  | Ashimi [21] | | 2015 | Systematic random sampling | Three clinics, Birnin Kudu, Jigawa state | | Infants presented to 3 clinics | 1-21+ days | 47.8% | 215 | 450 |
|  | Ezenyeaku [15] | | 2011 | Purposive | Enugu and Awka, Southeast Nigeria | | Attending clinic | 14-50 | 42.1% | 144 | 342 |
|  | Makinde[17] | | 2012 | Purposive | South-West of Nigeria | | Visiting emergency and gynecological wards | 0-14 | 41.9% | 237 | 565 |
|  | Ogah[19] | | 2019 | Consecutive | Ilorin, Kwara state | | Attending health clinic | 15-60 | 49% | 98 | 200 |
|  | Adeniran [22] | | 2015 | Multistage and purposive | 18 schools in Ilorin, North Central Nigeria | | Secondary school teachers | 20-65 | 42.2%* | 109 | 258 |
|  | Ibrahim[23] | | 2013 | All possible participants were included | Bayelsa state, Niger-Delta of Nigeria | | Nurses and doctors | <25 - 55 + (not only FGM/C sample) | 27.1% | 19 | 70 |
|  | Dike[24] | | 2012 | Purposive | Afikpo, Ebonyi State, Southeastern Nigeria | | Nursing and midwifery students | 15-45 | 54.3% | 146 | 269 |
|  | Iliyasu[26] | | 2012 | Multistage random sampling | Bayero University, Kano, Kano State, Northern Nigeria | | University Students | 17-40 | 12.1% | 43 | 359 |
|  | Lawani[16] | | 2014 | Purposive | Abakaliki, Ebonyi, southeast Nigeria | | Women seeking maternity services at 2 clinics | 15-49 | 66.3% | 342 | 516 |
|  | Garba[25] | | 2012 | Purposive | Kano, Northern Nigeria | | Infants attending clinic | < 5 –16+ days | 13% | 26 | 200 |
|  | Agbede[28] | | 2019 | Convenience | 4 wards Ede South LGA, Osun State | | Parents of daughters with FGM/C | 15-40+ | 38.8% | 146 | 376 |
|  | Dattijo[18] | | 2010 | Systematic random sampling | Jos, north-central Nigeria | | Attending antenatal care | < 20 –35+ | 31.3% | 81 | 260 |
|  | Ezeoke[13] | | 2021 | Multi-stage sampling of schools and purposive selection of students | North Central Nigeria | | Secondary school students | 13-19 | 35%** | 699 | 2000 |
|  | Chinawa[14] | | 2020 | Multi-stage sampling of schools; stratified selection of schools; random selection of students | Enugu metropolis | | Secondary school students | 13-21 years | 9.4% | 39 | 414 |
|  | Jimoh[27] | | 2018 | Purposive | Tsibiri village, a rural community in Giwa Local Government area of Kaduna state, northwestern region of Nigeria. | | Women of reproductive age | 15-49,10-14 (married) | 0.4% | 1 | 220 |
| **Mali** | Dicko-Traore  [12] | | 2014 | Purposive | Bamako | | Girls aged who were hospitalized | 0-15 | 73% | 224 | 305 |
| **Sierra Leone** | | Bjälkander [29] | 2012 | Random sampling | Bo Town, Bo District, in the southern region and in Makeni Town, Bombali District, in the northern region of Sierra Leone | | Attending antenatal care clinic (Facility based) | 11-45 | 100% | 258 | 258 |
| **Ethiopia** | Tamire[7] | | 2013 | Multi-staged cluster sampling | Hadiya zone, Southern Ethiopia | | High school students | 13 - 25 | 82.2% | 641 | 780 |
|  | Abathun[8] | | 2018 | Purposive (location), stratified (participants) | Somali and Harari Regions | | Sampled Girls and Boys who are attended primary and secondary schools from the selected schools | 16-22 | 32.1% | 79 | 246 |
|  | Gebremariam[6] | | 2016 | Muti-stage stratified random sampling | Jigjiga district, Somali regional state, eastern Ethiopia | | High school and college female students | 15-24 | 82.6% | 538 | 662 |
|  | Gudu[5] | | 2017 | Purposive | Jijiga | | Nulliparous women who gave birth | Mean 21.8 | 91.7% | 264 | 288 |
|  | Shay[9] | | 2010 | Random sampling | Addis Ababa | | School girls | < 5 – 20+ | 26% | 106 | 407 |
| **Ghana** | Nonterah[10] | | 2019 | Retrospective audit of records | Kassena-Nankana district- North Eastern Ghana | | Women who had delivered children at a hospital. | <20-35+ | 17.7% | 1647 | 9306 |
|  | Sakeah[11] | | 2018 | Random and convenience | Bawku municipality and Pusiga District | | Women of reproductive age | 15-49 | 61.3% | 509 | 830 |
| **Burkina Faso** | Ndiaye[1] | | 2010 | Purposive | Fada Ngourma | | Women who gave birth in 4 maternity wards | 14-44 | 59% | 210 | 354 |
|  | Ouédraogo[2] | | 2017 | Convenience | Boromo and Bittou | | Hawkers/Street vendors | 13-24 | 60.6% | 160 | 264 |
| **Uganda** | Ivanova[30] | | 2019 | Convenience | Nakivale refugee settlement | | Refugees | 13-19 | 10.4% | 27 | 260 |
| **Gambia** | Kaplan[3] | | 2013 | Purposive | Western Health Region | | Women examined for antenatal care or delivery in hospitals and health centers | 12-45 | 75.6% | 431 | 570 |
| **Tanzania** | Suleiman[4] | | 2021 | All possible participants, hospital records | Kilimanjaro region | | Women who delivered single births | 15-49 | 15.4% | 4675 | 30,286 |
| **SEAR** | | | | | | | | | | | |
| **Malaysia** | Rashid[52] | | 2019 | Snowball | Kedah and Penang, in the Northern region of Peninsular Malaysia | | Villagers | 18+ | 99.3% | 601 | 605 |
|  | Rashid[53] | | 2009 | Convenience, cross sectional | Five villages in north Malaysia. | | All women from villages | 1 month-91 years | 94.8% | 597 | 630 |
|  | Khalid[54] | | 2017 | Sequential convenience | Selangor | | Attending clinic | 18+ | 70.6% | 353 | 500 |
| **EMR** | | | | | | | | | | | |
| **Djibouti** | Minsart [31] | | 2014 | All possible participants | | Djibouti City | Women who gave birth | <25-35+ | 95.5% | 614 | 643 |
| **Somalia** | Adigüzel[32] | | 2018 | Purposive | | Mogadishu, Beladwayne, Kismaayo, Jawhar, Ceelbuur, and Baardhere, and regions such as Benadir, lower Shabelle, middle Shabelle, Hiiraan, Galgaduud, Mudug, Bakool, Gedo, and lower Jubba | Presented to the obstetrics and gynecology outpatient clinics,married with at least one daughter (Facility based) | Mean 28.76±8.77 | 99.7% | 355 | 356 |
| **Sudan** | Mahgoub[37] | | 2019 | Multi-stage sampling of schools | | Karary Locality, Khartoum State, Sudan | Students | 14-17 | 30.3% | 47 | 154 |
|  | Birge[38] | | 2021 | Cross sectional, Purposive | | Nyala, Darfur | Attending clinic | 18+ | 87.2 | 3767 | 4320 |
|  | Birge[33] | | 2017 | Convenience | | Darfur | Patients/working at hospital | 17 - 65 | 87.9% | 210 | 239 |
|  | Akbas[36] | | 2019 | All possible participants | | Nyala | University students | Mean/SD 19.5±1.95 | 80.1% | 330 | 412 |
|  | Sharfi[35] | | 2013 | Purposive | | Khartoum | University students and out-clinic patients | 20-62 | 73.4% | 1468 | 2000 |
|  | Ali[34] | | 2012 | Random | | Eight schools in Kassala, Eastern Sudan | Students | 9-16 | 83.3% | 810 | 972 |
| **Iran** | Dehghankhalili[51] | | 2015 | Purposive | | Minab, Dehbaz, Bandar-e-Lenge, Qeshm, Bandar-e-Khamir, and Bastak (rural areas) in Hormozgan, Southern Iran | Attending healthcare clinic | 14-38 | 68.5% | 535 | 780 |
| **Egypt** | Rasheed[47] | | 2011 | All possible participants | | Sohag and Qena, Egypt | Attending clinic | 5-25 | 89.2% | 3711 | 4158 |
|  | Mitwaly[42] | | 2017 | Purposive | | Luxor city ,Upper Egypt | Attending clinic | 15-49 | 89.1% | 1047 | 1175 |
|  | Arafa[43] | | 2018 | Multistage random sampling | | Beni-Suef | University students | Mean 20.89 (SD ± 1.68) | 47.3% | 815 | 1723 |
|  | Ahmed[46] | | 2017 | Purposive | | Suez Canal | Students attending clinic | 14-19 | 66.2% | 135 | 204 |
|  | Mostafa[44] | | 2017 | All possible participants | | Beni-Suef | Overweight and obese premenopausal women attending nutrition clinic | 20-49 | 59.3% | 89 | 150 |
|  | Abdel-Aleem[45] | | 2016 | Purposive | | Assiut and Sohag | Recently married attending 2 clinics | 17-31 | 87.4% | 376 | 430 |
|  | Hassanin[41] | | 2012 | Purposive | | Upper Egypt | Mothers in urban areas, completed secondary school, heard about the ban, with children in school | 8-14 | 71.6% | 358 | 500 |
|  | Abolfotouh[48] | | 2015 | Convenience | | NA | Medical students | 18+ | 14.7% | 47 | 320 |
|  | Elbendary[49] | | 2021 | Random selection | | Fayoum | Attending clinic | 18-45 | 62% | 62 | 100 |
|  | Galal[50] | | 2022 | All possible participants | | Upper and lower Egypt | Medical Students | Mean 21 years | 19.4% | 142 | 733 |
| **Iraq** | Yasin[39] | | 2013 | Convenience | | Erbil, Kurdistan region, Iraq | Attending clinic | 15-49 | 58.6%*** | 1164 | 1987 |
|  | Saleem[40] | | 2013 | Purposive | | Kurdistan region | Attending clinics | 0.5-20 | 23% | 348 | 1508 |

- Makinde[17] and Lawani[16] had a prospective design. All studies were cross-sectional, except Minsart[31] was a cohort study.

*Out of the female school teachers

**Without excluding those who were unsure if they had been mutilated.

*** Prevalence according to clinical examination.

Abbreviations: AFR: African Region EMR: Eastern Mediterranean Region SEAR: South East Asian Region FGM/C: Female Genital Mutilation/Cutting. FGM/C:

**S11 Table.** Types of FGM/C in School, Community or Facility based studies excluding studies on migrant populations.

| **Country** | **Author** | **Year** | **FGM/C Sample** | **Sample Size** | | **1** | **2** | **3** | **4** | **Other Type / Did Not Know (%)** |
| --- | --- | --- | --- | --- | --- | --- | --- | --- | --- | --- |
| **AFR** | | | | | | | | | | |
| **Nigeria** | Ashimi[20] | 2014 | 100 | 323 |  | |  |  |  | Gishiri cut (56.0%), Angurya (49.0%), type I and type II together (2.0%), one participant: removal of hymen. Angurya and Gishiri are types of nicks. |
|  | Ashimi[21] | 2015 | 215 | 450 | 11.2% | | 2.3% | 2.3% | 43.3% | Do not know (40.9%) |
|  | Dattijo[18] | 2010 | 81 | 260 |  | |  |  |  | Clitoridectomy (34.2%) Excision (3.25%), Dont know (63.9%) |
|  | Makinde[17] | 2012 | 237 | 565 | 35.44% | | 58.22% | 6.33% | 0 |  |
|  | Ibrahim[23] | 2013 | 19 | 70 |  | |  |  |  | Did not know if they were circumcised or not (4.3%) |
|  | Lawani[16] | 2014 | 342 | 516 | 28.1% | | 59.6% | 12.3% |  |  |
|  | Garba[25] | 2012 | 26 | 200 | 96.2% | | 3.8% |  |  |  |
|  | Iliyasu[26] | 2012 | 43 | 359 |  | |  |  |  | Flesh removed (37.2%), nicked (9.3%), cut and sewn (4.7%), do not know (20.9%) |
| **Tanzania** | Suleiman[4] | 2021 | 4675 | 30,286 | 61% | | 37% |  |  | Type III and IV reported together 2% |
| **Gambia** | Kaplan[3] | 2013 | 431 | 570 | 75.6% | | 24.4% | 0.004% |  |  |
| **Burkina Faso** | Ndiaye[1] | 2010 | 210 | 354 | 28% | | 28% | 3% |  |  |
| **Ethiopia** | Gebremariam[6] | 2016 | 538 | 662 | 49.3% | |  | 45.4% |  | Both type 1 and 3 (5.3%) |
|  | Gudu[5] | 2017 | 264 | 288 |  | | 7.6% | 92.4% |  |  |
| **SEAR** | | | | | | | | | | |
| **Malaysia** | Rashid[52] | 2019 | 601 | 605 | |  |  |  | 100% |  |
| **EMR** | | | | | | | | | | |
| **Egypt** | Mitwaly[42] | 2017 | 1047 | 1175 | | 58.5% | 41.5% |  |  |  |
|  | Abdel-Aleem[45] | 2016 | 376 | 430 | | 12.8% |  |  |  | Both type 2 and 3 (87.2%) |
|  | Elbendary[49] | 2021 | 62 | 100 | | 14% | 86% | 0 | 0 |  |
| **Iraq** | Yasin[39] | 2013 | 1164 (verified by clinical examination) | 1987 | | 99.6% | 0.4% |  |  |  |
|  | Saleem[40] | 2013 | 348 (with 239 examined cases) | 1508 | | 76.2% | 13.4% | 0% | 10.5% |  |
| **Iran** | Dehghankhalili[51] | 2015 | 535 | 780 | | 27.9% | 5.9% | 6% |  | Clitoral nicking (28.7%) |
| **Somalia** | Adigüzel[32] | 2018 | 355 | 356 | | 82.8% | 10.4% | 2.3% | 4.5% |  |
| **Djibouti** | Minsart [31] | 2014 | 614 | 643 | | 1.1% | 57.4% | 37% |  |  |
|  | Birge[38] | 2021 | 3767 | 4320 | | 8.5% | 35% | 50.4% |  |  |
|  | Birge[33] | 2017 | 210 | 239 | | 23.3% | 51% | 25.7% |  |  |
|  | Sharfi[35] | 2013 | 1468 | 2000 | |  |  | 62.9% |  |  |

Abbreviations: AFR: African Region EMR: Eastern Mediterranean Region SEAR: South East Asian Region FGM/C: Female Genital Mutilation/Cutting

**S12 Table.** Characteristics of FGM/C Procedure in School, Community or Facility based studies excluding studies on migrant populations.

|  | **Author** | **Year** | **Age at FGM/C** | **Performer of FGM** | **Location of Procedure** |
| --- | --- | --- | --- | --- | --- |
| **AFR** | | | | | |
| **Ethiopia** | Gudu[5] | 2017 | Most at 7 years (40.3%) (Range 2- 9yrs). |  |  |
|  | Gebremariam[6] | 2016 | ≤ 6 (20.4%), 7-10 (49.8%), 11-14 ( 29.8%) | Traditional circumcisers (72.9%), traditional birth attendant (25.5%), health professional (1.6%) | Home (63.6%) |
|  | Tamire[7] | 2013 | Mean 11(SD±2.3) years. | Traditional (69.1%), Medically Trained (29%), Don't Know (1.9%) | Home (82.5%) |
|  | Abathun[8] | 2018 | 6 to 14 (45.8%), don't know (23.8%) | Traditional (73%), Medically Trained (3.6%) |  |
|  | Shay[9] | 2010 | <1 year (20.8%), 1-5 years (50.9%), 6-10 (18.9%), >10 (9.4%) | Traditional (77.4%), Medically trained (22.6%) |  |
| **Nigeria** | Ezeoke[13] | 2021 | mean age 3.85±3.24 (range 1 to 8) years |  |  |
|  | Chinawa[14] | 2020 | <5 years (51.3%), 5 to 15 years (15.4%), ≥16 years (33.3%) | Nurse/midwife (48.7%), Doctors (15.4%), Traditional expert (15.4%) Mother (12.8%), Grand mother (7.7%) |  |
|  | Makinde[17] | 2012 | < 1 month (41.7%), 1 month-1 year (51.5%), 1-10 years (3.4%), more than 10 years (3.4%) | Traditional (64.6%), Medically trained (35.4%) |  |
|  | Dattijo[18] | 2010 | <1 year (44.6%), 1-5 years (41.9%) >5 years (13.5%) | Traditional (56.5%), Medically trained (8.2%), don't know (35.3%) |  |
|  | Ashimi[20] | 2014 |  | Traditional (100%) |  |
|  | Ashimi[21] | 2015 | Days: 1–7 (93.5%), 8–14 (4.4%), 15–21 (0.4%), 22–28 (0.9%), 29–35 (0%), 36–42 (0%), 43 ≥ (0.9%) | Traditional (100%) |  |
|  | Adeniran[22] | 2015 | Mean 4.76 (SD± 4.86) | Traditional (67.9%), Medically Trained (32.1%) | Home (63.9%), Clinic (36.1%) |
|  | Garba [25] | 2012 |  | Traditional (84.6%), Medically Trained (15.4%) |  |
|  | Iliyasu[26] | 2012 | 1 – 4 years (48.8%), 5 – 10 years (23.3%) infancy (4.7%), don't know (18.6%) | Traditional (74.4%), Medically Trained (11.6%), don't know (13.9%). | Home (55.8%), Clinic (4.7%), Circumciser's house (32.6%) |
|  | Agbede[28] | 2019 | Youngest child circumcised at age: 0–11 months (20.7%), 1–4 years (10.6%), 5–8 years (4.8%), 9–12 years (2.7%) |  |  |
| **Sierra Leone** | Bjälkander[29] | 2012 | 0-1 (0.7%), 2-4 (7.1%), 5-9 (18.2%), 10-14 (41.6%), 15+ (25.3%), don't know (7.1%) |  |  |
| **DR Congo** | Ivanova[30] | 2019 | Mean age: 7.3 (95% CI: 5.4, 9.1) |  |  |
| **EMR** | | | | | |
| **Iraq** | Yasin[39] | 2013 | <4 (16.6%), 4–7 (60.2%), 8–11 (21.2%), 12–15 (1.9%), ≥16 (0.1%) | Traditional (99.1%), Medically trained (0.9%) |  |
|  | Saleem[40] | 2013 | 0–2 (20.9%), 3–4 (32.6%), 5–6 (25%), 7 and older (20.9%) | Traditional (84.6%), Medically (15.4%) | City (25.9%), District (25.6%), Sub-District (32.9%), Village (15.7%) |
| **Egypt** | Hassanin[41] | 2012 | ≤8 (25.7%), 8–9 (32.9%), 9–10 (19.3%), ≥11 (22.1%) | Medically (54.5%), No answer (54.1%) |  |
|  | Elbendary[49] | 2021 | Median age 13.2 ±2.2 | Gynecological doctor (32.5%); Surgeon (11.5%); Midwife (56%); Barber (0%) | Hospital (22.5%), Home (60%), private (17.5%) |
|  | Mitwaly[42] | 2017 | 6 years (most frequent) followed by 5 years | Traditional (4.4%), Medically (95.7%) |  |
|  | Ahmed[46] | 2017 | Mean/SD 7.8 ± 1.1 | Traditional (32.6%) Medically (67.4%) |  |
|  | Rasheed[47] | 2011 | Mean/SD 8.2± 0.9 | Graph is unclear. General Practitioners by far the most common. | Home (74.3%), Clinic (25.7%) |
| **Somalia** | Adigüzel[32] | 2018 | 1-14 years | Traditional (74.4%), Medically Trained (24.2%), don't know (1.4%). |  |
| **Sudan** | Sharfi[35] | 2013 | <6 (96.9%) | Traditional (94.5%) |  |
|  | Akbas[36] | 2019 | 5-12 years (93%) mean age 7.97 ± 2.49 |  |  |
| **Iran** | Dehghankhalili[51] | 2015 | Mean 5.6 (SD± 3.6) (ranging from 2 to 38 years) | Traditional (100%) |  |
| **SEAR** | | | | | |
| **Malaysia** | Rashid[52] | 2019 | Median—6 years Range—birth to 30 years | Traditional (60.7%), Medically (39.3%) |  |
|  | Rashid [53] | 2009 | < 12 months old (88.6%), 13 - 24 months (6%), 24 months + (6%) | Traditional (67.8%), Medical (28.4%), don't know (3.8%) | Home (67.8%), Clinic (28.4%), unsure (3.8%) |

Abbreviations: AFR: African Region EMR: Eastern Mediterranean Region SEAR: South East Asian Region FGM/C: Female Genital Mutilation/Cutting
