## Supplementary material for "The global prevalence of female genital mutilation/cutting: A systematic review and meta-analysis of national, regional, facility and school-based studies": S7 Results

**S7 Results:** Studies on Migrant Populations.

**S13 Table.** Characteristics of Studies on Migrant Populations.

| **Host** | **Author** | **Year** | **Data Collection** | **Sampling Method, Study Design** | **Country of Origin** | **Sub-Region** | **Population Description** | **Age** | **Exam/ Patient report** | **Data Collection Site** | **Total FGM** | **Risk of Bias** |
| --- | --- | --- | --- | --- | --- | --- | --- | --- | --- | --- | --- | --- |
| EUR | | | | | | | | | | | | |
| **Belgium, Denmark, UK, France** | Leye[1] | 2018 | NA | Purposive, Case series | Unclear | NA | Involved in court cases | 15-49 | NA | NA | Belgium- 21, Denmark- 2, France- 30, UK- 179 | Mod. |
| **Finland** | Koukkula[2] | 2016 | 2010-2012 | Stratified random sampling, Cross Sectional | Somalia (43%), Kurdistan (57%) | Helsinki, Espoo, Vantaa, Turku, Tampere, Vaasa | Migrants | 18-64 | Patient Report | Home, Clinic, Other | 186 | Mod. |
| **Germany** | Loucas[3] | 2017 | 2015-2016 | Purposive, Cross Sectional | Syria (33.6%), Egypt (16.5%), Kosovo (6.9%), Albania (6.7%), Somalia (4.6%), Eritrea (4.8%), Serbia (3.7%), Afghanistan (23.2%) | Mainz, Ingelheim | Migrants | 0.5-18 | Patient  Report | Refugee Centers | 23 | High |
|  | Zinka[4] | 2018 | 2017 | Convenience, Retrospective evaluation of the FGM/C reports | Nigeria (n = 109), Somalia (n = 27), Sierra Leone (n=6), Eritrea (n=6), Ethiopia (n=4), Tanzania (n=1) | Munich | Asylum seekers | 3 weeks – 45 years | Exam | Community / aid organizations | Women: 45 | Mod. |
|  |  |  |  |  |  |  |  |  |  |  | Girls: 7 |  |
|  | Koschollek[5] | 2020 | 2015-2016 | Convenience, Cross Sectional | Sub-Saharan Africa | Munich, Rhine-Ruhr region, Cologne, Berlin, Frankfurt am Main, Hanover | Migrants | 18-25 (21.3%), 26-35 (39.8%), 36-45 (26.9%),>45 (12.1%) | Patient Report | Public Places | 281 | Mod. |
|  | Hänselmann[6] | 2011 | 2008-2009 | Purposive, Case series | Nigeria, Burkina Faso, Eritrea (43.5%), Ghana, Sudan, Somalia (26.9%) | Nationwide with a focus on the Rhine-Neckar Area | Migrants | 19-59 | Patient report | Hospital/ Clinic  (multiple) | 24 participated (out of 37) | High. |
| **Italy** | Castagna[7] | 2018 | 2007- 2016 | Entire Database, Case series | Nigeria (n=64), Democratic Republic of Congo (n=24),, Ivory Coast (n=23), Cameroon (n=13), Somalia (n=4), Guinea (n=2), Ethiopia (n=2), Gambia (n=1), Eritrea (n=1), Gabon (n=1), Mali (n=1) | Turin | Migrants | 14-48 | Exam | Rape center | 18 | Mod. |
| **Malta** | Padovese[8] | 2013 | 2010-2011 | Hospital records, Cross Sectional | Somalia (70.1%), Eritrea (10.7%), Sudan (4%), Ethiopia (3.7%), Mali (3.1%), Ivory Coast (2.1%), Nigeria (2.1%), Chad (0.5%), Burkina Faso (0.5%), Niger (0,5%), Others (2.7%) | NA | Asylum Seekers | 0-46 (not only FGM) | Both | Hospital/ Clinic | 163 | Mod. |
| **Netherlands** | Korfker[9] | 2012 | 2009 | All midwifery clinics, Cross sectional | Burkina Faso, Ivory Coast, Djibouti, Egypt, Eritrea, Ethiopia, Gambia, Guinea Bissau, Liberia, Mali, Mauretania, Sierra Leone, Somalia, Sudan, Chad | NA | Migrants | NA | Exam | Hospital/ Clinic  (All midwifery practices in the country) | 470 | Mod. |
|  | Kawous[10] | 2020 | 2018 | Cross sectional, from records |  | NA | Women who gave birth in a primary care midwifery practice | Reproductive age | Exam | Hospitals/Clinics | 523 | Low |
| **Norway** | Taraldsen[11] | 2021 | 2004-2015 | All possible cases from hospital records, Case series | Somalia (74%), Eritrea (10%), Ethiopia (5%) | NA | All women who had been examined for an FGM/C related issue at an outpatient clinic | Mean 26 years | Exam | Hospital Database | 891 | Low |
|  | Mbanya[12] | 2018 | 2014 | Respondent driven sampling, Cross Sectional | Somalia | Oslo | Migrants | 16-25 | Patient Report | Community based | 82 | Mod. |
| **Portugal** | Division of Sexual, Reproductive, Child and Youth Health; Division of Health Statistics and Monitoring; Shared Services Ministry of Health (SPMS)[13] | 2018 | 2014-2017 | Population based, electronic database | The Republic of Guinea-Bissau; Senegal; Nigeria; Gambia; Ivory Coast; Eritrea; Somalia; Benin; Egypt; Ghana; Sierra Leone | Lisbon and Tagus Valley Health Region | Women who attended hospital with FGM | 14-64 | Exam | The Electronic Health Record (RSE) | 237 | Low |
| **Greece** | Vrachnis[14] | 2012 | 2009 | Purposive, Case series | Ethiopia, Eritrea, Egypt, Somalia | Athens | Migrants | 19-31 | Both | Hospital/ Clinic | 7 | Low |
| **Switzerland** | Frick[15] | 2021 | 2010-2016 | Retrospective cohort study | Somalia (34%) Eritrea (28%) Burkina Faso (6%) Ethiopia (5%) Guinea (5%) Sudan (5%) Other (16%) | Geneva | Women attending specialised FGM clinic | NA | Exam | Hospital/ clinic | 360 | Low |
|  | Cottler-Casanova[16] | 2021 | 2016-2018 | Cross-sectional | Participants were from 30 FGM/C practicing countries | Geneva, Lausanne, Bern, Zurich | Women and girls admitted to Swiss university hospital | 0-49 years | Not Clear | Hospital/ clinic | 207 | Mod. |
| **Sweden** | Wahlberg[17] | 2017 | 2015 | Purposive, cCRT | Somalia | Gothenburg and Malmö | Migrants | 18+ | Patient Report | Somali organization | 187 | Mod. |
|  | Wahlberg[18] | 2019 | 2015 | Purposive and Snowball, Cross-Sectional | Somalia | Malmo, Gothenburg, Stockholm, Uppsala | Migrants | 18+ | Patient Report | Somali organisation, cafe´s, Swedish courses, mosques. | 270 | Mod. |
| **Turkey** | Kartal[19] | 2021 | 2019 | Case series, cross sectional | Somalia | NA | University students | Mean age 20.30 ± 1.01 | Patient Report | University | 117 | Mod. |
| **England** | Hodes [20] | 2016 | 2006-2014 | Purposive, Case series | 67% from Somalia, % unavailable for Kenya, Ethiopia, Gambia, Zambia, Malaysia | London | Migrants | Children | Exam | Hospital/ Clinic | 27 | Mod. |
|  | Ali[21] | 2020 | 2014-2019 | Prospective case series, Hospital records | Multiple countries (40% Somalia). 96% African, 4% Asian | London | Patients attending clinic | Mean 13 years (2-16) | Exam | Hospital/ Clinic | 55 | Mod. |
|  | Fawcett[22] | 2018 | 2015-2016 | Retrospective database analysis, case series |  | Staffordshire | Patients attending emergency department | 23-40 years old | Exam | Hospital/ Clinic | 34 | Mod. |
|  | Creighton[23] | 2016 | 2014 - 2015 | Purposive, Case series | Somalia, Saudi Arabia, Gambia, UK, unknown | Inner London | Migrant children/ children of migrants | 0-18 | Exam | Hospital/ Clinic | 18 | Mod. |
| **Scotland** | Ford[24] | 2018 | 2010-2013 | Entire database (Successive Cross Sectional) | Nigeria, Sudan, The Gambia, Democratic Republic of Congo, Egypt, Iraq, Sierra Leone, Somalia and Tanzania, Malaysia, India, Brunei, United Arab Emirates, Saudi Arabia and England | Lothian | Migrants accessing maternity services | NA | Exam | Hospitals /Clinics | 107 | Mod. |
| **UK + Ireland** | Hodes[25] | 2020 | 2015-2017 | Purposive based case series | Sudan, Somalia, Gambia, and Eritrea, Europe, the Middle East, South-East Asia |  | Children | 0-16, mean 3 | Exam | Hospital/clinic | 103 | Low |
| AMR | | | | | | | | | | | | |
| **USA** | Sudhinaras-et[26] | 2019 | 2017-2013 | Population Database, Descriptive | South Asia (73.9%), Southeast Asia (7.8%), Africa (7.4%), Latin America and the Caribbean (4.4%), and Europe/Central Asia (6.4%). | California | Refugees | 0-65+ | Patient Report | Household | 162 | Mod. |
|  | Akinsulure-Smith[27] | 2016 | 1996-2014 | Entire database, Descriptive | Guinea, Cameroon, Sierra Leone, Zaire/Democratic Republic of Congo, Cote d’Ivoire, Burkina Faso, Congo, Liberia, Mauritania | New York City | Survivors of Torture | 34.3  Mean  9.7  (SD) | Patient Report | Hospital/ Clinic | 133 | Mod. |
|  | Chu[28] | 2015 | 2014 | Convenience, Case series | Sierra Leone (67.6%), Guinea (20.6%), Mali (17.6%), Gambia (16.2%) | New York City | Migrants | 18 +, 35.2  (Mean) 13.4  (SD) | Patient Report | Location of choice. | 46 | Mod. |
|  | Akinsulure-Smith[29] | 2014 | NA | Purposive, Cross-Sectional | Sierre Leone (48%) and Liberia (52%) | New York City | Migrants | 20-57 | Patient Report | Community site | 7 | Mod. |
|  | Geynisman-Tan[30] | 2019 | NA | Convenience, Cross Sectional | Unclear | New York, Boston, Chicago, Minneapolis, San Francisco | All but one were migrants | 24-40 | Patient Report | Online | 30 | Mod. |
|  | Johnson-Agbakwu[31] | 2022 | 2017 | Community-based purposive snowball sample, Cross-sectional | Somalia | Phoenix and Tucson, Arizona | Somali migrants and refugees | 15+ years | Patient Report | Community | 687 | Mod. |
|  | Wikholm[32] | 2020 | 1996-2020 | Convenience from records | countries with high FGM/C prevalence | New York | Asylum seekers | 18+ years | Both | Hospital/Clinic | 100 | High |
|  | Michlig[33] | 2021 | 2017 | Purposive snowball sampling, cross sectional | Somaliland | Phoenix and Tucson, Arizona | Bantu Somali | 15+ years | Patient report | Community, health needs assessment survey | 680 | Mod. |
|  | Ukoha[34] | 2015 | NA | Convenience, Cross Sectional | Nigeria | DFW, Texas | Migrants,  Igbo (100%) | 19-55 | Patient Report | Online | Mothers: 64 | Mod. |
|  |  |  |  |  |  |  |  |  |  |  | Girls: 21 |  |
| WPR | | | | | | | | | | | | |
| **Australia** | Zurynski[35] | 2017 | 2012-2017 | Purposive, Case series | Kenya, Sudan, Australia, Eritrea, Ethiopia, Sierra Leone, Somalia, East Africa | NA | Refugees, Migrants | 0-18 | Exam | Online | 59 | Mod. |
|  | Varol[36] | 2016 | 2006-2012 | Entire Hospital Record, Case control | Tanzania, Burundi, Rwanda, Uganda, Sudan, Ethiopia, Eritrea, Djibouti, Somalia, and Kenya, Sierra Leone, Liberia, Guinea, Nigeria and the Democratic Republic of Congo, Egypt, Iran, Iraq, Saudi Arabia and Yemen, Pakistan, Sri Lanka, Indonesia and Singapore. | Unnamed Region | Pregnant Migrants | 15-40+ | Exam | Hospital/ Clinic | 196 | Mod. |
|  | Gibson-Helm[37] | 2014 | 2002–2011 | Entire Database, Case series | North Africa, Middle and East Africa, West Africa | NA | Migrants/ refugees at pregnancy clinic | NA | Exam | Hospital/ Clinic | 78 | High |
|  | Davis[38] | 2019 | 2011-2015 | Entire Database of Retrospective cohort case series. | Somalia, Sudan, Sierra Leone, Ethiopia, Egypt, Indonesia, Other | Sydney | Women who gave birth | <20 to over 35 | Exam | Hospital/ Clinic | 142 | Mod. |
|  | Shukralla[39] | 2020 | 2014 | Retrospective purposive study | Born in Africa, Malaysia, New Zealand | Western Australia | Women who gave birth | 15-39 years old | Exam | Hospital/ Clinic | 53 | Mod. |
| EMR | | | | | | | | | | | | |
| **Saudi Arabia** | Rouzi [40] | 2020 | 2016-2017 | All women attending clinic, Cross-Sectional | Saudi 49.7% Naturalised 13.1% Non Saudi 37.1% | Jeddah | Migrants and nationals | Mean 33·4 ± 9·95 years | Exam | Hospital/ Clinic | 175 | Mod. |
|  | Rouzi[41] | 2017 | 2014 - 2015 | Hospital records, longitudinal | Sudan | Jeddah | Migrants | 23-49 years old | Exam | Hospital/ Clinic | 107 | Mod. |
|  | Rouzi[42] | 2017 | 2014 - 2015 | Consecutive convenience, Cross Sectional | Sudan | Jeddah | Migrants | 39.76 (mean) | Patient Report | Hospital/ Clinic | 179 | Mod. |
|  | Malak[43] | 2020 | 2019 | Random, Cross Sectional | Saudi (89.7%) Non-Saudi (10.3%) | NA | Locals & Migrants | 18+ | Patient Report | Electronic | 50 | Mod. |
| **UAE** | Al Awar[44] | 2020 | 2016-2017 | Purposive, cross sectional | African country, Arab country, Asian country, European country North/South America, Australia, NZ, UAE | Al Ain and Abu Dhabi | Locals & Migrants | 18-50+ years old | Patient Report | Three University Campuses | 344 (mothers) 114 (girls) | Mod. |

* Tissue removed and sewn closed, tissue removed and some stitching, some tissue removed, pricking. †Flesh removed, Genital area just nicked, Genital area sewn closed

Abbreviations: EMR: Eastern Mediterranean Region. SEAR: South East Asian Region. EUR: European Region. WPR: Western Pacific Region AMR: American Region

**S14 Table.** Prevalence of FGM/C in Migrant Populations.

| **Country** | **Author** | **Year** | **Study Design and Sampling Method** | **Country of Origin** | **Subregion** | **Population description** | **FGM/C (%)** | **Total FGM** | **Sample Size** |
| --- | --- | --- | --- | --- | --- | --- | --- | --- | --- |
| **EMR** | | | | | | | | | |
| **Saudi Arabia** | Malak[43] | 2020 | Random, Cross Sectional | Saudi and Non Saudi Nationals | NA | Migrants and nationals | 9.4% | 50 | 530 |
|  | Rouzi[40] | 2020 | All possible women, Cross-Sectional | Sudan | Jeddah | Migrants and nationals | 18.2% | 175 | 963 |
|  | Rouzi [42] | 2017 | Consecutive convenience, Cross Sectional | Sudan | Jeddah | Migrants | 67.3% | 179 | 266 |
| **UAE** | Al Awar[44] | 2020 | Purposive, cross sectional | African country, Arab country, Asian country, European country North/South America, Australia, NZ, UAE | Al Ain and Abu Dhabi | Locals & Migrants | 41.1% (mothers) | 344 (mothers) | 831 (mothers) |
|  |  |  |  |  |  |  | 26.4% (girls) | 114 (girls) | 333 (girls) |
| **EUR** | | | | | | | | | |
| **Portugal** | Division of Sexual, Reproductive, Child and Youth Health; Division of Health Statistics and Monitoring; Shared Services Ministry of Health (SPMS) [13] | 2018 | Population based, electronic database | The Republic of Guinea-Bissau; Senegal; Nigeria; Gambia; Ivory Coast; Eritrea; Somalia; Benin; Egypt; Ghana; Sierra Leone | Lisbon and Tagus Valley Health Region | Attended hospital with FGM |  | 237 | NA |
| **Netherlands** | Kawous[10] | 2020 | Cross sectional, from records |  |  | Migrants | 0.54% | 523 | 96932 |
|  | Korfker[9] | 2012 | All midwifery clinics, Cross sectional | Burkina Faso, Ivory Coast, Djibouti, Egypt, Eritrea, Ethiopia, Gambia, Guinea Bissau, Liberia, Mali, Mauretania, Sierra Leone, Somalia, Sudan, Chad | NA | Migrants | 0.32% | 470 | 145,492 |
| **Malta** | Padovese[8] | 2013 | Hospital records, Cross Sectional | Somalia, Eritrea, Sudan, Ethiopia, Mali, Ivory Coast, Nigeria, Chad, Burkina Faso, Niger, Others | NA | Asylum Seekers | 42.5% | 163 | 384 |
| **Norway** | Mbanya[12] | 2018 | Respondent driven sampling, Cross Sectional | Somalia | Oslo | Migrants | 51.6% | 82 | 159 |
| **Germany** | Koschollek[5] | 2020 | Convenience, Cross Sectional | Sub-Saharan Africa | Munich, the Rhine-Ruhr region, Cologne, Berlin, Frankfurt am Main, and the region of Hanover | Migrants | 26.9% | 122 | 288 |
|  | Loucas[3] | 2017 | Purposive, Cross-Sectional | Syria, Egypt, Kosovo, Albania, Somalia, Eritrea, Serbia, Afghanistan | Mainz, Ingelheim | Refugees | 11% | 23 | 209 |
|  | Zinka[4] | 2018 | Convenience, retrospective reports | Nigeria, Somalia, Eritrea, Sierra Leone, Ethiopia, Tanzania | Munich | Asylum seekers | Women 67% | 45 | 67 |
|  |  |  |  |  |  |  | Girls 8.14% | 7 | 86 |
| **Sweden** | Wahlberg[18] | 2019 | Purposive and Snowball, Cross-Sectional | Somalia | Gothenburg and Malmö | Migrants | 85% | 270 | 318 |
|  | Wahlberg[17] | 2017 | Purposive, cCRT | Somalia | Gothenburg and Malmö | Migrants | 97.9%% | 187 | 191 |
|  | Cottler-Casanova[16] | 2021 | Cross-sectional, purposive | Participants were from 30 FGM/C practicing countries | Geneva, Lausanne, Bern, Zurich | Migrants | 2.3% | 207 | 8927 |
| **Scotland** | Ford[24] | 2018 | Entire database | Lothian, originally from: Nigeria, Sudan, The Gambia, Democratic Republic of Congo, Egypt, Iraq, Sierra Leone, Somalia and Tanzania, Malaysia, India, Brunei, United Arab Emirates, Saudi Arabia, England and unknown | Lothian | Migrants attending maternity services | 0.24% | 107 | 44,460 |
| **Finland** | Koukkula[2] | 2016 | Convenience, Cross Sectional | Somalia and Kurdistan | Helsinki, Espoo, Vantaa, Turku, Tampere, Vaasa | Migrants | 47.81% | 186 | 389 |
| **AMR** | | | | | | | | | |
| **USA** | Sudhinaraset[26] | 2019 | Population database, descriptive | Africa (145/162 patients), Europe/Central Asia, Latin America and the Caribbean, South Asia | California | Refugees | 2.1% | 162 | 8751 |
|  | Ukoha[34] | 2015 | Convenience, Cross Sectional | Nigeria | DFW metropolitan area in Texas | Migrants – mothers  and daughters | Mothers- 46% | 64 | 139 |
|  |  |  |  |  |  |  | Daughters- 33.3% | 21 | 67 |
|  | Akinsulure-Smith[27] | 2016 | Entire database, Descriptive | Guinea, Cameroon, Sierra Leone, Zaire/Democratic Republic of Congo, Cote d’Ivoire, Burkina Faso, Congo, Liberia, Mauritania | New York City | Survivors of Torture | 25.9% | 133 | 514 |
|  | Johnson-Agbakwu[31] | 2022 | Community-based purposive snowball sample, Cross-sectional | Somalia | Phoenix and Tucson, Arizona | Somali migrants and refugees | 79% | 687 | 848 |
|  | Wikholm[32] | 2020 | Convenience from records | countries with high FGM/C prevalence | New York | Asylum seekers | 84% | 100 | 119 |
|  | Michlig[33] | 2021 | Purposive snowball sampling, cross sectional | Somaliland | Phoenix and Tucson, Arizona | Refugees/ Migrants | 85.3% | 680 | 797 |
|  | Akinsulure-Smith[29] | 2014 | Purposive, Cross-Sectional | Sierra Leone and Liberia | New York City | Migrants | 30% | 7 | 23 |
| **WPR** | | | | | | | | | |
| **Australia** | Davis[38] | 2019 | Entire Database of Retrospective cohort | Somalia, Sudan, Sierra Leone, Ethiopia, Egypt, Indonesia, Other | Sydney | Refugees/ Migrants | 1.64% | 142 | 8622 |
|  | Shukralla[39] | 2020 | Retrospective purposive study | Born in Africa, Malaysia, New Zealand | Western Australia | Migrants | 0.71% | 53 | 8480 |
| **Case series** | | | | | | | | | |
| **Saudi Arabia** | Rouzi [41] | 2017 | Hospital records, longitudinal case series | Sudan | Jeddah | Migrants |  | 107 |  |
| **Belgium** | Leye[1] | 2018 | Purposive, Case series | NA | NA | Involved in court cases | 100% | 21 | 21 |
| **Denmark** | Leye[1] | 2018 | Purposive, Case series | NA | NA | Involved in court cases | 100% | 2 | 2 |
| **Croatia** | Leye[1] | 2018 | Purposive, Case series | NA | NA | Involved in court cases | 100% | 179 | 179 |
| **France** | Leye[1] | 2018 | Purposive, Case series | NA | NA | Involved in court cases | 100% | 30 | 30 |
| **Greece** | Vrachnis[14] | 2012 | Purposive, Case series | Ethiopia, Eritrea, Egypt, Somalia | Athens | Migrants | 100% | 7 participated, 11 identified | 7 |
| **Turkey** | Kartal[19] | 2021 | Purposive, Case series | Somalia |  | Migrants |  | 117 |  |
| **Norway** | Taraldsen[11] | 2021 | All possible cases from hospital records, Case series | Somalia (74%), Eritrea (10%), Ethiopia (5%) | NA | Migrants | 97% | 891 | 913 |
| **Italy** | Castagna[7] | 2018 | Entire Database, Case series | Nigeria, Democratic Republic of Congo, Ivory Coast, Cameroon, Somalia, Guinea, Ethiopia, Gambia, Eritrea, Gabon, Mali | Turin | Migrants | 12.5% | 18 | 143 |
| **Switzerland** | Frick[15] | 2021 | Retrospective, case series | Somalia (34%) Eritrea (28%) Burkina Faso (6%) Ethiopia (5%) Guinea (5%) Sudan (5%) Other (16%) | Geneva | Migrants | 100% | 360 | 360 |
| **Germany** | Hänselmann[6] | 2011 | Purposive, Case series | Nigeria, Burkina Faso, Eritrea (43.5%), Ghana, Sudan, Somalia (26.9%) | Germany, Focus on the Rhine-Neckar region | Migrants |  | 24 participated, 37 identified | 37 |
| **England** | Hodes [20] | 2016 | Purposive, Case series | Kenya, Ethiopia, Gambia, Zambia, Malaysia | London | Migrants | 57.45% | 27 | 47 |
|  | Fawcett[22] | 2018 | Retrospective database analysis, case series |  | Straffordshire | Refugees/ Migrants |  | 34 |  |
|  | Creighton[23] | 2016 | Purposive, Case series | Somalia, Saudi Arabia, Gambia, UK, unknown | Inner London | Migrant children/ children of migrants | 100% | 18 | 38 suspected |
| **UK + Ireland** | Hodes[20] | 2020 | Purposive, case series | Sudan, Somalia, Gambia, and Eritrea, Europe, the Middle East, South-East Asia |  | Migrants |  | 103 | 103 |
| **US** | Chu[28] | 2015 | Convenience, Case series | Sierra Leone, Guinea, Mali, and Gambia | New York City | Migrants | 70% | 48 | 68 |
|  | Geynisman-Tan[30] | 2019 | Convenience, Case series | Unclear | New York, Boston, Chicago, Minneapolis, and San Francisco, USA | All but one were migrants | 100% | 30 | 30 |
| **Australia** | Zurynski[35] | 2017 | Purposive, Case series | Kenya, Sudan, Australia, Eritrea, Ethiopia, Sierra Leone, Somalia, East Africa | NA | Refugees and Migrants | 100% | 59 | 59 |
|  | Varol[36] | 2016 | Entire Hospital Record, Case control | Tanzania, Burundi, Rwanda, Uganda, Sudan, Ethiopia, Eritrea, Djibouti, Somalia, and Kenya, Sierra Leone, Liberia, Guinea, Nigeria and the Democratic Republic of Congo, Egypt, Iran, Iraq, Saudi Arabia and Yemen, Pakistan, Sri Lanka, Indonesia and Singapore. | NA | Pregnant migrants | 100% | 196 | 196 |
|  | Gibson-Helm[37] | 2014 | Entire Database, Case series | Africa | NA | Migrants/ refugees at pregnancy clinic | 100% | 78 | 78 |

Abbreviations: EMR: Eastern Mediterranean Region. SEAR: South East Asian Region. EUR: European Region. WPR: Western Pacific Region AMR: American Region FGM/C: Female Genital Mutilation/Cutting

**S15 Table.** Types of FGM/C in Migrant Populations.

| **Author** | **Year** | **Country of Origin** | **Host Country** | **Total FGM** | **Sample Size** | **Type 1 (%)** | **Type 2 (%)** | **Type 3 (%)** | **Type 4 (%)** | **Don't Know/Missing Type (%)** | **Other (%)** |
| --- | --- | --- | --- | --- | --- | --- | --- | --- | --- | --- | --- |
| **EMR** | | | | | | | | | | | |
| Rouzi[40] | 2020 | Sudan | Saudi Arabia | 175 | 963 |  |  | 6.3% | 26.3% | 46.3% | I and II together (21.1%) |
| Rouzi[41] | 2017 | Sudan | Saudi Arabia | 107 |  | 39% | 25% | 36% |  |  |  |
| Al Awar[44] | 2020 | African country, Arab country, Asian country, European country North/South America, Australia, NZ, UAE | UAE | 344 - Mothers | 831- mothers | Mothers -62.8% | Mothers -16.6% | Mothers -5% |  |  |  |
|  |  |  |  | Daughters- 114 | Daughters-333 | Daughters-81.6% | Daughters-18.4% |  |  |  |  |
| **WPR** | | | | | | | | | | | |
| Zurynski[35] | 2017 | Kenya, Sudan, Australia, Eritrea, Ethiopia, Sierra Leone, Somalia, East Africa | Australia | 59 | 59 | 16.9% | 8.5% | 8.5% | 10.2% |  |  |
| Varol[36] | 2016 | Tanzania, Burundi, Rwanda, Uganda, Sudan, Ethiopia, Eritrea, Djibouti, Somalia, and Kenya, Sierra Leone, Liberia, Guinea, Nigeria and the Democratic Republic of Congo, Egypt, Iran, Iraq, Saudi Arabia and Yemen, Pakistan, Sri Lanka, Indonesia and Singapore. | Australia | 196 | 196 | 33.2% | 33.2% |  |  |  |  |
| Davis[38] | 2019 | Somalia, Sudan, Sierra Leone, Ethiopia, Egypt, Indonesia, Other | Australia | 142 | 8622 | 21.2% | 24.1% | 41.1% |  | 13.6% |  |
| Shukralla[39] | 2020 | Born in Africa, Malaysia, New Zealand | Australia | 53 | 7494 | 30% | 32% | 4% |  |  |  |
| **AMR** | | | | | | | | | | | |
| Geynisman-Tan[30] | 2019 | Unclear | USA | 30 | 30 | 40% | 23.3% | 23.3% |  | 13% |  |
| Chu[28] | 2015 | Sierra Leone, Guinea, Mali, and Gambia | USA | 48 | 68 | 40% |  |  | 8.9% |  | Type 1 or 2 (91.9%) |
| Ukoha[34] | 2015 | Nigeria | USA | Mothers -64 | 139 |  | 84.4% | 48.4% |  | 50.8% |  |
|  |  |  |  | Daughters -21 | 67 |  | 70% | 52% |  | 50% |  |
| Johnson-Agbakwu [31] | 2022 | Somalia | USA | 687 | 848 | 32% | 20% | 35% |  |  |  |
| Wikholm [32] | 2020 | Asylum seekers from countries with high FGM/C prevalence | USA | 100 | 119 | 4.6% | 84.6% | 9.2% |  |  |  |
| Michlig [33] | 2021 | Somaliland | USA | 680 | 797 | 36.9% | 23% | 40.1% |  |  |  |
| **EUR** | | | | | | | | | | | |
| Korfker[9] | 2012 | Burkina Faso, Ivory Coast, Djibouti, Egypt, Eritrea, Ethiopia, Gambia, Guinea Bissau, Liberia, Mali, Mauretania, Sierra Leone, Somalia, Sudan, Chad | Netherlands | 470 | 145,492 |  |  | 40% |  | 10% | 50% |
| Kawous[10] | 2020 |  | Netherlands | 523 | 96932 |  |  | 32% |  |  |  |
| Vrachnis[14] | 2012 | Ethiopia, Eritrea, Egypt, Somalia | Greece | 7 | 7 | 1 case | 4 cases | 2 cases |  |  |  |
| Zinka[4] | 2018 | Nigeria, Somalia, Eritea, Sierra Leone, Ethiopia, Tanzania | Germany | Women- 45 | 67 | 18 cases | 30 cases | 4 cases |  |  |  |
|  |  |  |  | Girls - 7 | 86 |  |  |  |  |  |  |
| Hänselmann[6] | 2011 | Nigeria, Burkina Faso, Eritrea (43.5%), Ghana, Sudan, Somalia (26.9%) | Germany | 24 | NA (223 clinics) | 12.6% | 58.3% | 25% | 4.1% |  |  |
| Padovese[8] | 2013 | Somalia, Eritrea, Sudan, Ethiopia, Mali, Ivory Coast, Nigeria, Chad, Burkina Faso, Niger, Others | Malta | 163 | 384 | 15.3% | 4.9% | 51.5% | 28.2% |  |  |
| Castagna[7] | 2018 | Nigeria, Democratic Republic of Congo, Ivory Coast, Cameroon, Somalia, Guinea, Ethiopia, Gambia, Eritrea, Gabon, Mali | Italy | 18 | 143 | 44.44% | 38.9% | 11.1% |  |  |  |
| Hodes[20] | 2016 | Kenya, Ethiopia, Gambia, Zambia, Malaysia | England | 27 | 47 | 7.4% | 29.6% | 0% | 40.7% |  |  |
| Creighton[23] | 2016 | Somalia, Saudi Arabia, Gambia, UK, unknown | England | 18 | 38 | 5.55% | 30.86% | 22.2% | 61.1% |  |  |
| Ali[21] | 2020 | Multiple countries (40% Somalia) | England | 55 | 148 | 27% | 25% | 5% | 24% |  |  |
| Hodes[20] | 2020 | Sudan, Somalia, Gambia, and Eritrea, Europe, the Middle East, South-East Asia | UK + Ireland | 103 |  |  |  | 21% | 21% |  | Type I or II (58%) |
| Wahlberg[18] | 2019 | Somalia | Sweden | 270 | 318 | 7% | 12% | 25%*  41%† |  | 1% |  |
| Wahlberg[17] | 2017 | Somalia | Sweden | 187 | 191 | 5% | 11% | 32%* 51%† |  |  |  |
| Kartal [19] | 2021 | Somalia | Turkey | 117 |  | 43.2% | 49.9% |  |  |  |  |
| Division of Sexual, Reproductive, Child and Youth Health; Division of Health Statistics and Monitoring; Shared Services Ministry of Health (SPMS) [13] | 2018 |  | Portugal | 237 |  | 3.4% | 54.9% | 41.3% |  |  |  |
| Taraldsen[11] | 2021 |  | Norway | 891 | 913 | 7.5% | 7.7% | 83.3% | 1.3% |  |  |
| Frick[15] | 2021 | Somalia (34%) Eritrea (28%) Burkina Faso (6%) Ethiopia (5%) Guinea (5%) Sudan (5%) Other (16%) | Switzerland | 360 |  |  | 55.6% |  |  |  |  |
| Cottler-Casanova[16] | 2021 |  | Switzerland | 207 | 8927 | 12.6% | 27.5% | 44.9% | 1.4% | 13.5% |  |

*Flesh removed and some stitching † Flesh removed and sown closed.

Abbreviations: EMR: Eastern Mediterranean Region. SEAR: South East Asian Region. EUR: European Region. WPR: Western Pacific Region AMR: American Region FGM/C: Female Genital Mutilation/Cutting

**S16 Table.** Characteristics of FGM/C Procedure for Migrant Populations.

|  | Country | Author | Year | Age at FGM/C | Performer of FGM | Location of Procedure |
| --- | --- | --- | --- | --- | --- | --- |
| **EUR** | England | Hodes[20] | 2016 | under 1 (14.8%), 1-3 (11.1%), 4-6 (18.5%), 7-9 (29.6%), 10-12 (7.4%), 13 (3.7%), unknown (18.5%) | Traditional (41%), Medically (35%), not described (37%) |  |
|  |  | Ali[21] | 2020 |  |  |  |
|  |  | Creighton[23] | 2016 | mean age 6.8 years (range 7 months– 10 years). | Traditional (38%), Medically (62%) | Clinic (38%). Other: One girl - in her bedroom in London at the age of 10. She was cut along with her sister and two cousins. Type 2 FGM/C was confirmed on examination. Another child - while on a family holiday, consistent with type 4 FGM. One child’s parents took her to Malaysia at the age of 7 months and a prick was made to the clitoris by a practitioner at the local hospital with parental consent. The parents were unaware that this traditional practice constituted FGM. |
|  | Greece | Vrachnis[14] | 2012 | Between 3-8 (range), mean 4.9 | Traditional – one patient | Outdoors “in the bush” – one patient |
|  | Germany | Hänselmann[6] | 2011 | Infancy (28.6%), 11-15 years (14.3%) |  |  |
|  | UK + Ireland | Hodes[20] | 2020 |  | 45% health professionals, 36% traditional, 12% relative |  |
|  | Turkey | Kartal[19] |  | Before the age of 7 | 64% medical professional, 46% traditional |  |
|  | Norway | Taraldsen[11] |  | Median 7 years |  |  |
|  | Portugal | Division of Sexual, Reproductive, Child and Youth Health; Division of Health Statistics and Monitoring; Shared Services Ministry of Health (SPMS) [13] |  | 6.6 years mean, 0-37 range |  |  |
| **EMR** | **Saudi Arabia** | Malak[43] | 2020 | One day to a month (52.0%), two months to a year (18.0%), two to five years (6.0%), and six to 15 years (10.0%), don't remember (14%) |  |  |
|  |  | Rouzi[40] | 2020 | Within one week after birth (57.7%) | Traditional (20%), Medically (58.8%), Other (1.2%), Don't know (20%) | Home (58.9%), Clinic (23.4%), midwife's house (4.6%), other (13.2%) |
| **AMR** | USA | Wikholm[32] | 2020 | Average 9 years |  |  |
|  |  | Geynisman-Tan[30] | 2019 | Between ages 1 week and 16 years, median, 6 years |  |  |
|  |  | Chu[28] | 2015 | Under 1 year old (14.6%), 1 to 5 years old (17.4%), 6 to 10 years old (37%), 11 to 15 years old (17.4%), 15 years old or older (4.3%) | Traditional (90.6%), Medically (9.5%) | Country where the procedure occurred: In home country (95.5%) In other West African country (4.5%) |
|  |  | Ukoha[34] | 2015 |  | Mothers: Traditional (73.5%), Medically Trained (17.2%), 9.4 (don't know/other). Daughters: Traditional (60.9%), Medically Trained (39.1%), |  |
| **WPR** | **Australia** | Zurynski[35] | 2017 |  |  | Where FGM/C occurred based on country of birth: Sudan: Sudan (3) Malaysia (2) Eritrea (1) Kenya: Kenya (4) Sudan (3) Somalia (2) Eritrea: Eritrea (2) Somalia (1) Australia: Australia (2) Indonesia (1) Sierra Leone: Sierra Leone (2) Uganda: Somalia (1) Somalia: Somalia (1) Egypt : Egypt (1) |

Abbreviations: EMR: Eastern Mediterranean Region. SEAR: South East Asian Region. EUR: European Region. WPR: Western Pacific Region AMR: American Region FGM/C: Female Genital Mutilation/Cutting
