## Supplementary figures and images for "The global prevalence of female genital mutilation/cutting: A systematic review and meta-analysis of national, regional, facility and school-based studies"

### S1 Figure

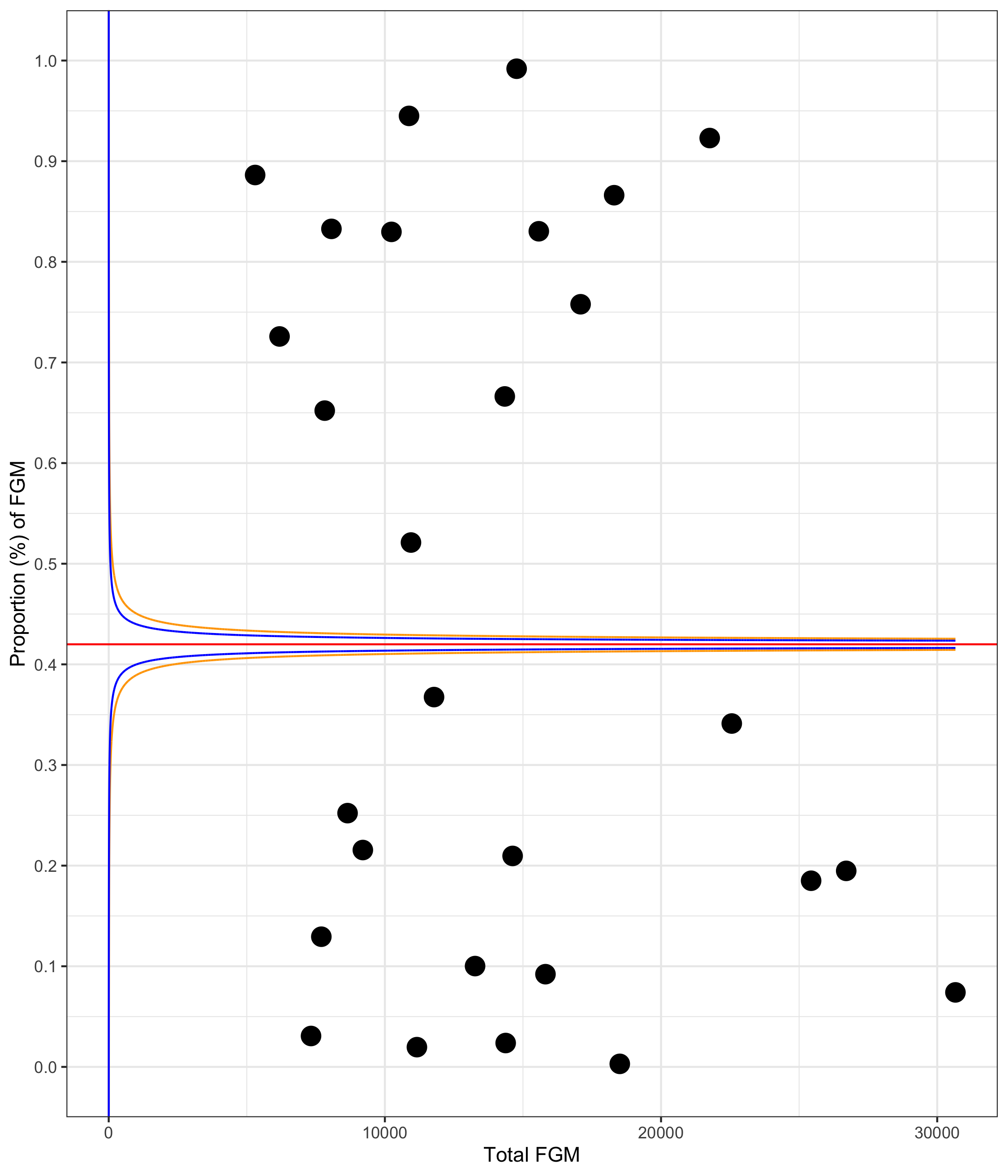

### S1 Figure

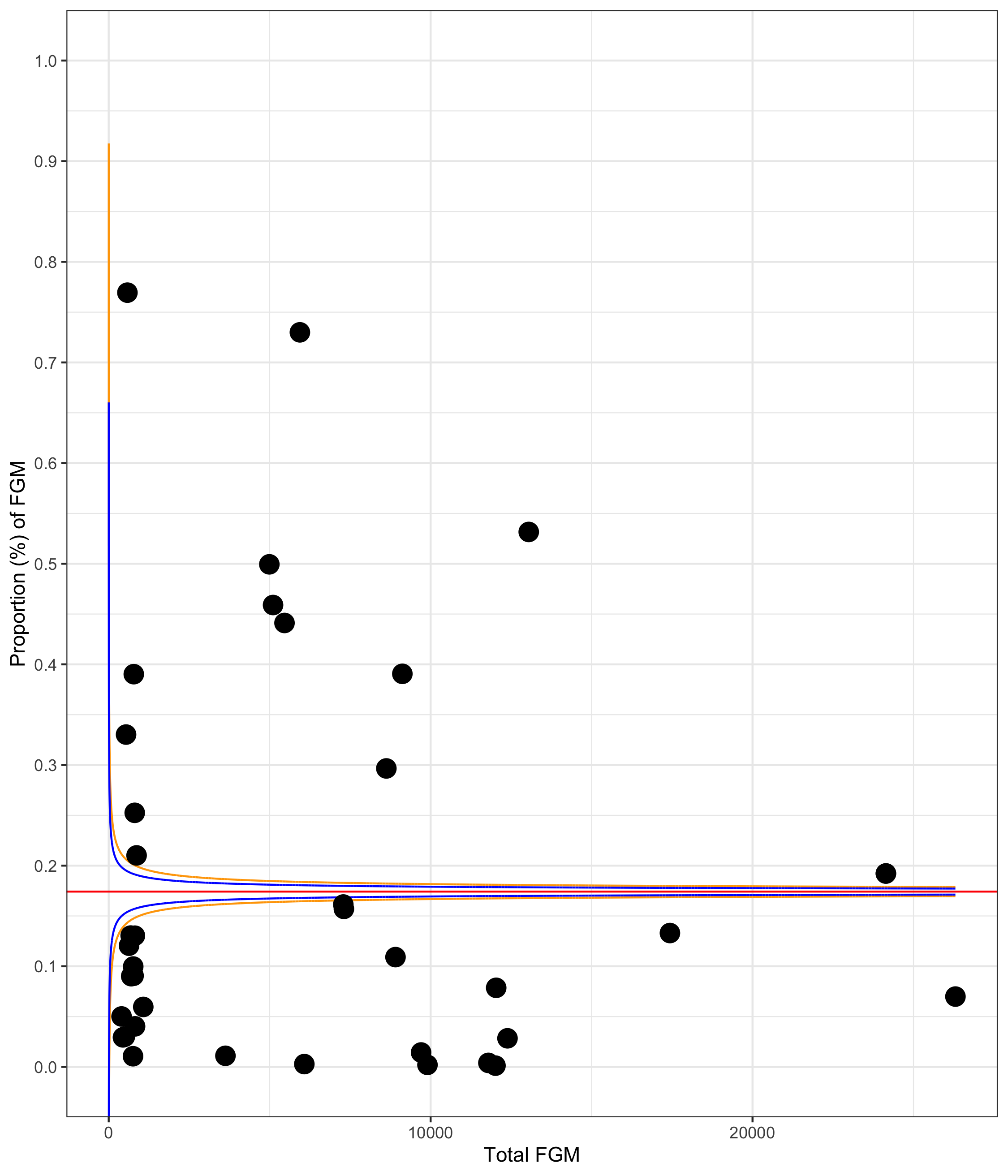
